# Exploring the genetic overlap between 12 psychiatric disorders

**DOI:** 10.1101/2022.04.12.22273763

**Authors:** Cato Romero, Josefin Werme, Philip R. Jansen, Joel Gelernter, Murray B. Stein, Daniel Levey, Renato Polimanti, VA Million Veteran Program, Christiaan de Leeuw, Danielle Posthuma, Mats Nagel, Sophie van der Sluis

## Abstract

The widespread comorbidity among psychiatric disorders (PDs) demonstrated in epidemiological studies^1–5^ is mirrored by non-zero, positive genetic correlations from large scale genetic studies^6–10^. We employed several strategies to uncover pleiotropic SNPs, genes and biological pathways^7,8^ underlying this genetic covariance. First, we conducted cross-trait meta-analysis on 12 PDs to identify pleiotropic SNPs. However, the majority of meta-analytic signal was driven by only one or a few PDs, hampering interpretation and joint biological characterization of the meta-analytic signal. Next, we performed pairwise comparisons of PDs on the SNP, gene, genomic region, gene-set, tissue-type, and cell-type level. Substantial overlap was observed, but mainly among pairs of PDs, and mainly at less stringent *p*-value thresholds. Only heritability enrichment for “conserved genomic regions” and “nucleotide diversity” was significant for multiple (9 out of 12) PDs. Overall, identification of shared biological mechanisms remains challenging due to variation in power and genetic architecture between PDs.

---

We obtained GWAS summary statistics for 12 PDs (see **Table 1** for an overview and acronyms, **Supplementary Note 1** for sources; **Supplementary Fig. 1** for a flowchart of this study; **Supplementary Fig. 2**). GWAS sample sizes varied between PDs (median: 116,961, range: 9,725 (OCD) - 386,533 (INS); **Supplementary Table 1**). After harmonization and filtering to all SNPs shared between summary statistics (see **Methods, Supplementary Note 2**), SNP-based heritability 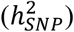 and genome-wide genetic correlations (*r*_*g*_’s) were computed using LD Score regression (LDSC)^9,11^. The median 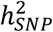 was 12.4% (range: 4.4% - 37.7%), substantially lower than family-based heritability estimates^12^ (**Supplementary Table 2**). Overall, the power of the 12 PD GWASs, as well as the strength of the genetic signal uncovered by them, varied considerably, which may affect subsequent detection of genetic overlap. Estimated *r*_*g*_’s were moderate (median = 0.24, range: -0.13 - 0.83) and not always significant (**Supplementary Table 3**). Substantial *r*_*g*_’s were observed both between pairs of PDs with low 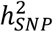 (e.g., for ANX and DEP, 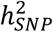 is 4.4% and 10.3%; *r*_*g*_ = 0.83) as well as relatively high 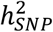 (e.g., for BIP and SCZ, 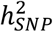 is 19.6% and 24.4%; *r*_*g*_ = 0.67) (**Fig. 1A-1B**; **Supplementary Fig. 3-4**). Genomic structural equation modelling (gSEM)^13^, used to scrutinize the genetic covariance structure between traits, confirmed good fit (CFI = 0.93, SRMR = 0.074) of a previously reported genetic four-factor model of PDs^8^ although not all factor loadings were significantly different from zero (**Supplementary Fig. 5**). Additionally, using the Complex Trait Genetics Virtual Lab platform^14^ (University of Queensland & QIMR Berghofer Medical Research Institute), we observed broadly similar *r*_*g*_ patterns of the 12 PDs with 1,376 external traits. Together, these findings indicate genetic similarity among PDs (**Supplementary Fig. 6-12**), suggesting the possible existence of SNPs, genes or biological pathways affecting general PD-liability.

**Table 1.** Overview of all PDs included in this study. Overview of all 12 PDs, abbreviations as used throughout the manuscript, the sample size (N) on which summary statistics are based, associated Pubmed ID or URL (for preprint) and the number of SNPs included in the original summary statistics, before we applied filtering (see **Methods)**.

| PD | Abbreviation | N | Pubmed ID/ URL | Original Nsnps |
| --- | --- | --- | --- | --- |
| Attention deficit/hyperactivity disorder | ADHD | 53,293 | 30478444 | 8,047,420 |
| Alcohol use disorder | ALC | 202,004 | 30940813 | 6,895,250 |
| Anorexia nervosa | ANO | 72,517 | 31308545 | 8,219,102 |
| Anxiety | ANX | 275,822 | 31748690 & 31906708 | 7,014,585 |
| Autism spectrum disorder | ASD | 46,350 | 30804558 | 9,112,386 |
| Bipolar disorder | BIP | 51,710 | 31043756 | 13,413,244 |
| Depression | DEP | 173,005 | 29700475 | 13,554,550 |
| Insomnia | INS | 386,533 | 30804565 | 10,862,567 |
| Obsessive-compulsive disorder | OCD | 9,725 | 28761083 | 8,409,516 |
| Post-traumatic stress disorder | PTSD | 214,138 | 33510476 | 8,839,303 |
| Schizophrenia | SCZ | 161,405 | https://doi.org/10.1101/2020.09.12.20192922 | 7,585,077 |
| Tourette syndrome | TS | 14,307 | 30818990 | 8,265,318 |

**Figure 1.**
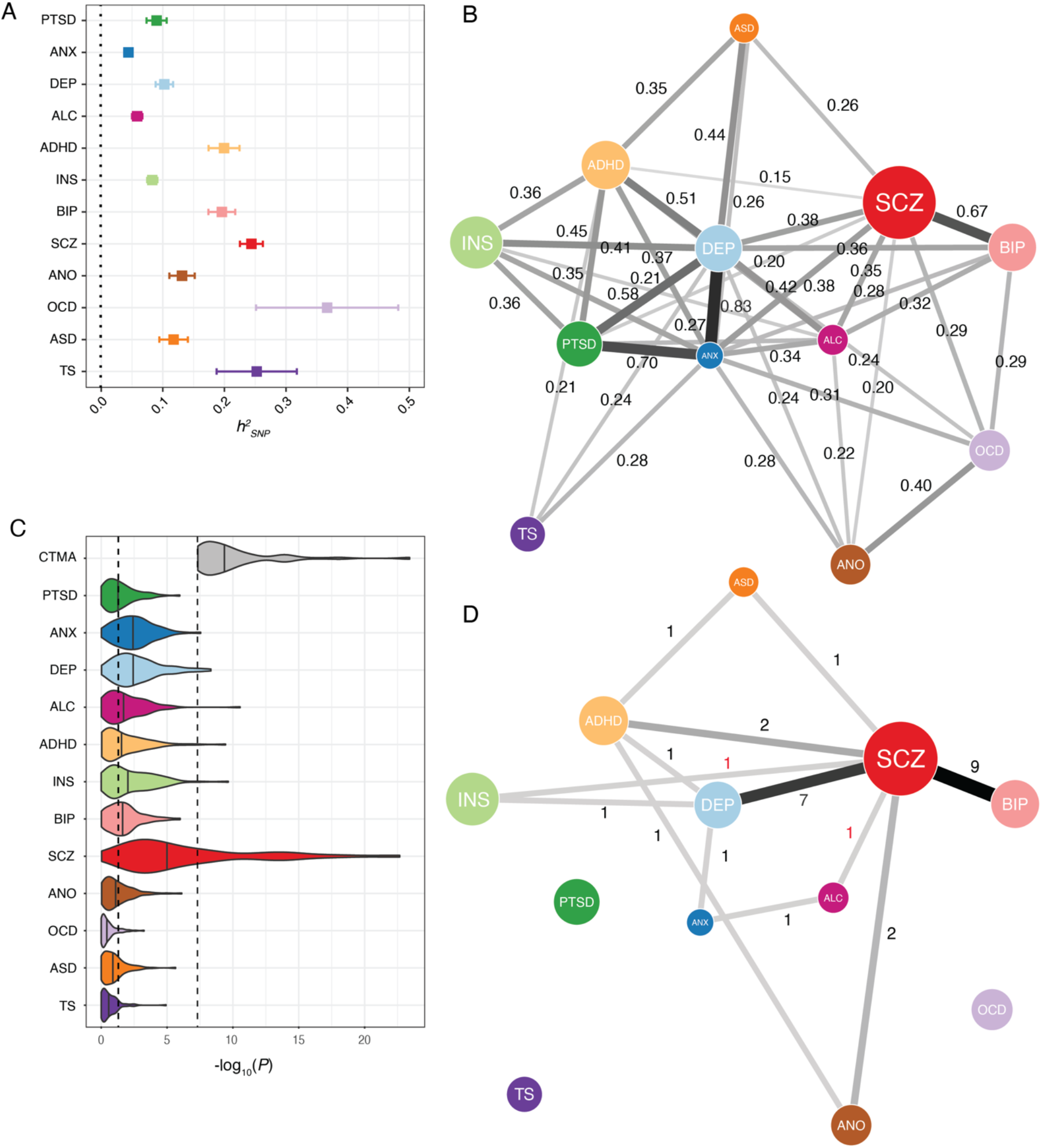
SNP-based heritability, global and local genetic correlations and top SNP p-values of the 12 PDs. A) Error-bar plot of the SNP-based heritability estimates for the 12 PDs, computed using LDSC. Magnitude as well as precision of the estimates varied substantially across PDs. Order of PDs is based on clustering of the global genetic correlations among the 12 PDs (**Supplementary Fig. 14**); B) Network visualization of the genome-wide (i.e., global) genetic correlations between the PDs, computed using bivariate LDSC. Connections represent significant genetic correlations, with thicker connections denoting stronger genetic correlations. The size of the nodes is weighed by the sample size and SNP-based heritability of the given PD 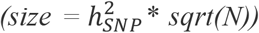. Many genetic correlations were moderate to high and significant after Bonferroni correction; C) P-values of the 12 individual PDs for the 92 lead SNPs identified in the cross-trait meta-analysis. Vertical lines within the violin plots represent the median p-value for specific PDs. X-axis displays - log10 transformed p-values. The left and right dashed lines denote p = 0.05 and p = 5×10^−8^, respectively. SCZ showed disproportionately strong signal, whereas PTSD, ALC, ADHD, BIP, ANO, OCD, ASD, and TS showed median p-values close to or above the p = 0.05 threshold; D) Network visualization of the number of significant local genetic correlations between the PDs, computed using bivariate LAVA. Connections represent how many significant local r_g_’s were identified between pairs of PDs, with thicker connections denoting higher numbers. Numbers in black font indicate positive local r_g_’s, whereas numbers in red font indicate negative local r_g_’s. The orientation and the size of the nodes were set to mirror that of the network visualization of global r_g_’s.

Cross-trait meta-analysis of genetically related traits has previously been used to increase the power to detect pleiotropic SNPs and genes as well as shared biological annotations among different types of cancers, PDs, and immunological and inflammatory diseases^7,8,15–19^. Accordingly, we conducted a sample-size weighted cross-trait GWAS meta-analysis using mvGWAMA^20^ (**Methods**), aiming first to identify pleiotropic SNPs that contribute to general genetic PD-liability. The cross-trait meta-analysis identified 4,674 genome-wide significant (GWS; *p* < 5×10^−8^) SNPs, of which approximately 50% (*n* = 2,413) were not identified for any of the 12 PDs individually. Using FUMA^21^, a publicly available platform for annotation of GWAS results, we identified 92 LD-independent lead SNPs for the cross-trait meta-analysis that mapped to 75 independent genomic risk loci (**Methods**; **Supplementary Table 4**; **Supplementary Fig. 13**). Of these 75 loci, 28 did not overlap with loci reported in previous cross-PD meta-analyses^7,8^ (which reported 146 and 184 loci, respectively) possibly due to differences in the (number of) traits or datasets included, analysis methods used, or genomic locus definition (see **Supplementary Table 5** for a comparison of summary statistics). To identify associated genes based on our meta-analytic SNP-results, we used (1) genome-wide gene-based association study (GWGAS) in MAGMA^22^, and (2) positional mapping, expression quantitative trait locus (eQTL) mapping, and chromatin interaction mapping in FUMA (**Methods**). The SNPs mapped to a total of 345 genes, including 96 (49 through GWGAS; *α*_*BON*_ = 0.05 / nGenes = 0.05 / 17,287 = 2.68×10^−6^) that were not identified for the individual PDs **(Supplementary Table 6-7)**.

Genetic signal observed in the cross-trait meta-analysis may be driven by signal shared among all included PDs, or by strong signal in just one or a few of the included PDs^23^. To investigate how well each of the 12 PDs was represented by our meta-analytic results, we computed genetic correlations between the meta-analysis and the underlying 12 PDs and inspected the *p*-values of individual PDs for the 92 meta-analytic lead SNPs. The *r*_*g*_’s between the meta-analysis and the 12 PDs were all significant, but varied substantially, from *r*_*g*_ = 0.27 for TS to *r*_*g*_ = 0.84 for DEP (overall median = 0.55), partly as a function of their individual sample sizes (**Supplementary Table 3**; **Supplementary Fig. 15)**. The *p*-values of the 12 individual PDs for the 92 meta-analytic lead SNPs are shown in **Fig. 1C**. SCZ clearly showed the strongest association for the meta-analysis lead SNPs (median = 1.69 × 10^−5^), while 8 PDs had median *p*-values close to or above the *p* = 0.05 threshold (**Supplementary Fig. 16**). Additionally, 62% of mapped genes and 55% of GWGAS genes identified in the meta-analysis overlapped with genes identified for SCZ, while ANO showed the next highest percentage at only 11% for both mapped and GWGAS genes (**Supplementary Table 6-7**). These findings suggest that a considerable part of our cross-trait meta-analysis signal was primarily driven by SCZ — the PD with the strongest genetic signal — hindering interpretation of the cross-trait meta-analytic signal as representing “general liability to PDs”. We repeated the meta-analysis without SCZ and found that the number of risk loci, lead SNPs, and mapped genes identified in the meta-analysis dropped by 40-60% (**Supplementary Table 8**). Additionally, only 29 out of the 49 GWGAS genes previously identified in the cross-trait meta-analysis but not in individual PDs, remained significant. This confirmed our hypothesis that the cross-trait meta-analytic signal was strongly determined by SCZ and not necessarily representative of genetic variance common to multiple PDs.

Closer examination of the signal of the 92 cross-trait meta-analytic lead SNPs showed that for the majority of lead SNPs, only 3 or fewer of the 12 PDs contributed to the GWS signal, even at less-stringent p-value thresholds, and the contributing PDs were not the same for every lead SNP (e.g., for p < .0001 and p <.001, 5% and 15% of the 92 lead SNPs were associated to >3 of the 12 PDs, respectively; **Supplementary Table 9, Supplementary Fig. 16**). While detection of pleiotropic SNPs through joint evaluation of multivariate (sub-significant) signal is considered a strength of meta-analysis^23^, joined functional annotation of SNPs with such heterogeneous association patterns may not make sense from a biological perspective.

Given the apparent interpretational challenges of our cross-trait meta-analytic results, we subsequently conducted pairwise comparisons between individual PD summary statistics on the locus, SNP, gene, gene-set, and gene-property level to assess overlap of PDs^24–27^. Genetic correlations estimated on the locus-level, rather than genome-wide, can detect genomic regions that harbor genetic factors associated to multiple PDs. To this end, we used LAVA^28^ to estimate multivariate genetic correlations in those regions that showed univariate signal for multiple PDs (**Methods**). Out of 2,495 defined genomic regions, 1,057 (42.4%) showed univariate genetic signal (*p* < 1×10^−4^) for more than one PD, and 97 (3.9%) for more than half of the PDs (**Supplementary Table 10**). However, when testing the local genetic correlations within these regions, only 29 regions reached significance after Bonferroni correction and no region showed significant correlation for more than two traits (*α*_*BON*_ = .05/(number of bivariate test conducted) = 0.05 / 2,532 = 1.97×10^−5^; **Fig. 1D; Supplementary Table 11**; **Supplementary Fig. 17**). The observation that no more than two PDs are correlated within any given region is further supported by direct comparisons of SNP- and gene-based results between the 12 individual PDs. Of the 13,749 SNPs and 507 GWGAS genes that were GWS for any of the 12 PDs, only 215 SNPs (*p* < 5×10^−8^; **Supplementary Table 12**) and 39 GWGAS genes were GWS (*p* < 2.89×10^−6^) for more than 1 PD, of which only 2 genes (*SORCS3* & *AC074091*.*13*) were GWS for more than 2 PDs (**Supplementary Table 6**). The majority of this overlap, as well as the majority of the significant local *r*_*g*_’s, was observed between two genetically highly correlated PDs, both with considerable 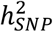: SCZ and BIP (genome-wide *r*_*g*_ = 0.67), sharing 117 SNPs, 28 genes, and 9 positive local *r*_*g*_’s. In contrast, the highest genetically correlated PDs, ANX and DEP (genome-wide *r*_*g*_ = 0.83), shared no overlapping GWS SNPs or genes, and only 1 positive local *r*_*g*_, probably due to their low overall genetic signal. Out of all 29 significant local *r*_*g*_’s, 15 regions contained GWGAS genes significant for either PD in the PD pair, suggesting that the local genetic correlation is driven by co-variance in these genes (**Supplementary Table 13;** see **Supplementary Note 3** for detailed examples).

Although only few overlapping genes and regions were observed between PDs other than SCZ and BIP, different genes can potentially converge in the same biological pathways or be enriched in similar tissues or cell types. To examine this, we conducted MAGMA gene-set and gene property analysis. After correcting for 10,126 tests and 12 PDs (i.e., 9,508 gene-sets^29^, and gene expression levels of 53 tissues types^30^ and 565 cell types^31^; *α*_*BON*_ = .05 / 10,126 / 12 = 4.10×10^−7^) no test surpassed the significance threshold. Taking a less stringent approach, correcting separately for gene-sets, tissues or cell types, we observed convergence among sets of PDs for 10 brain tissues and among pairs of PDs for 6 cell types, but not for gene-sets (**Fig. 2**; **Supplementary Table 14-16**). In addition, even more lenient enrichment tests of gene function through consultation of the SynGO database^32^, and investigation of gene paralog members confirmed little convergence across PDs’ mapped genes (**Supplementary Note 4**). To assess whether certain functional genomic categories show enrichment for heritability among multiple PDs, we performed stratified heritability analysis using LDSC^33^. After correction for the 61 tested functional categories across all PDs (*α*_*BON*_ = .05/ (61 × 12) = 6.83×10^−5^), significant enrichment of genetic signal was observed for both general and specific conserved genomic areas as well as for nucleotide diversity in all PDs except PTSD, OCD, and TS (**Supplementary Table 17**; **Supplementary Fig. 18**).

**Figure 2.**
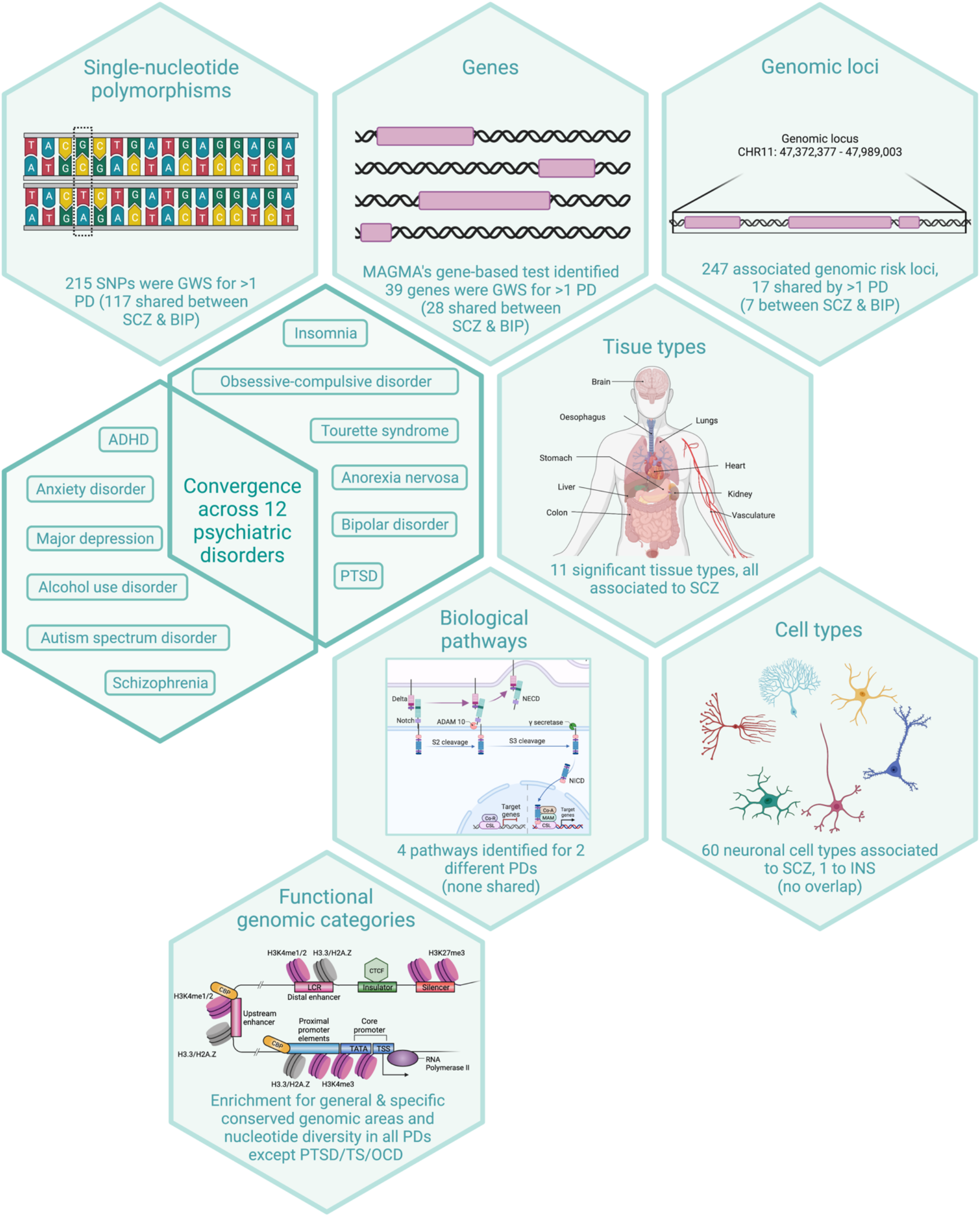
Schematic overview of all annotation analyses conducted on 12 PDs. This figure summarizes all the ways we annotated the genetic association results for the 12 individual PDs, and the extent to which we identified annotations shared between PDs. First, we examined overlap in SNPs, mapped genes, and genomic loci. Subsequently, we conducted gene-set and gene-property analyses, examining, for each of the 12 PDs, whether genetic signal was enriched in 9,508 gene sets, 565 cell types, and 53 tissue types. Partitioned LDSC was used to test for enrichment of 61 functional genomic categories. Note: all pictures used in this figure are illustrations and do not reflect the actual pathways identified in the analysis. The figure of functional genomic categories was adapted from Ong & Corces (2011)^34^. Created with BioRender.com (see **URLs**).

As GWS SNPs, genes, and biological annotations showed little overlap between PDs, we hypothesized that genetic overlap might be enriched at sub-significant *p*-value levels. To explore this, we conducted Fisher’s exact tests to examine overlap in the top associated SNPs and genes across all pairwise combinations of PDs for different *p*-value thresholds (**Methods**; **Supplementary Table 18-19**; **Supplementary Fig. 19-20**). Significant enrichment of overlapping, independent SNPs was only observed between ANO-SCZ and DEP-ADHD for SNPs detected at different *p*-value thresholds (from *p* = 0.1 to *p* = 1×10^−4^; *α*_*BON_SNP*_ = 0.05 / (number of PDs pairs × number of *p*-value thresholds)) = 0.05 / (66 × 12) = 6.31×10^−5^). Significant enrichment of overlapping genes, however, was observed for PDs across all thresholds, but especially *p* = 0.05 to *p* = 0.001, reaching a peak at *p =* 0.05, where 53 out of 66 possible PDs pairs showed significant gene enrichments (*α*_*BON_GENE*_ = 0.05 / (number of PDs pairs × number of *p*-value thresholds) = 0.05 / (66 × 8) = 9.47×10^−5^; **Fig. 3**).

**Figure 3.**
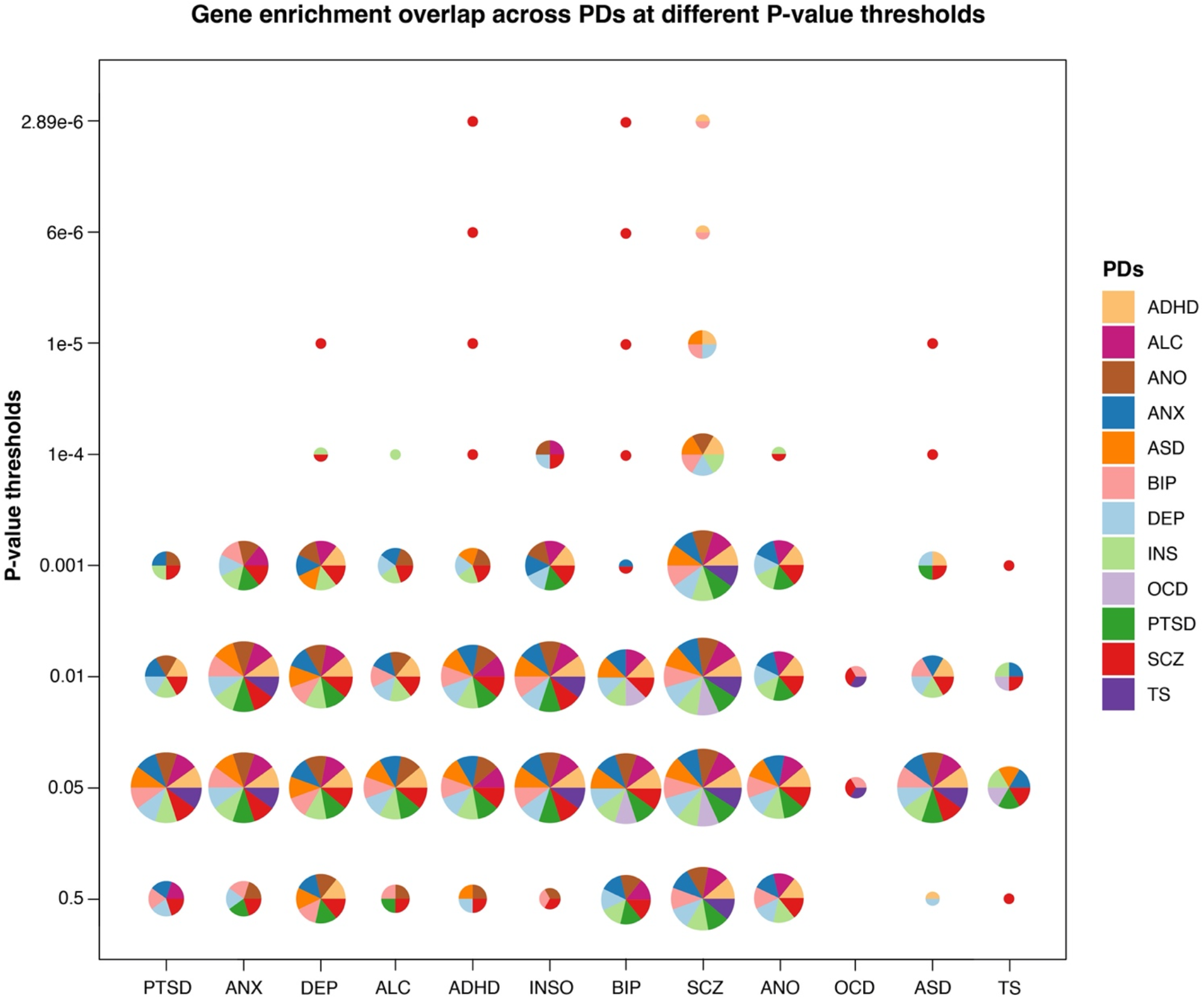
Results of Fisher’s exact tests for gene enrichment overlap across 12 PDs. Fisher’s exact tests were conducted to test for overlap in associated genes between the 12 PDs for different p-value thresholds (y-axis). Colors in pie-charts indicate which PDs showed significant enrichment with the PD on the x-axis, while circle size is scaled by the number of observed trait enrichments. Overall, genetically correlated PDs only show significant gene overlap at high (i.e., non-significant) p-value thresholds. Across PDs, the highest number of significant gene enrichment overlap is observed at p = 0.05, while for p = 1×10^−5^, all observed gene enrichments are between SCZ and other PDs.

In conclusion, using publicly available summary statistics of 12 PDs, we demonstrated the possibilities and interpretational challenges of cross-trait meta-analysis, and extended upon previous cross-PD research by simultaneously investigating overlap of genomic loci, SNPs, genes, gene-sets, tissue-types and cell-types among a large set of PDs. Like previous studies^24^, we mainly observed overlap among *pairs* of PDs, suggesting that genetic overlap could be largely pair-specific. Although no loci or SNPs, and only 2 genes were associated with more than two PDs, we did observe extensive overlap among multiple PDs for 3 functional genomic categories and widespread sub-significant gene-overlap enrichment for most of the included PDs. While PDs consistently show moderate-to-high genetic correlations, identification of shared SNPs, genes or biological mechanisms remains challenging. This challenge is aggravated by differences between PDs in 1) genetic architecture (i.e., the accessibility and clarity of the underlying genetic signal), 2) diagnostic accuracy or reliable phenotypic operationalization, and 3) sample sizes and the ease with which additional participants can be recruited. Additionally, we should temper expectations about identification of shared biological mechanisms when sizable genetic correlations are observed for traits whose overall genetic signal is low to begin with. Finally, even if shared genetic mechanisms are identified, they will generally only partly explain the observed phenotypic comorbidity between PDs, the remainder being due to shared environmental factors. The current study highlights multiple pressing challenges that affect the potential success of cross-trait genetic research. Resolving such challenges is necessary to advance our understanding of biological mechanisms underlying comorbid disorders.

## Methods

### Information on summary statistics

Access to GWAS summary statistics was obtained for 12 psychiatric disorders (PDs) (see **Supplementary Table 1** for an overview of the PDs and publication sources, and **Supplementary Note 1** for information on the individual studies). For details on the analysis protocols, we refer to the original publications.

### Harmonization of summary statistics

Prior to analysis, we harmonized the summary statistics files as consistent file structure facilitates efficient analysis in specialized software like genomic SEM^8^, LDSC^4,13^, and mvGWAMA^9^. First, we added a column representing the unique SNP ID (defined as ‘CHR:BP:A1_A2’, where CHR represents the chromosome number, BP the basepair position of the variant and A1 and A2 are the alleles in alphabetical order), and used this variable as SNP identifier in various downstream analyses. Second, for summary statistics in which information on the per-SNP sample size was missing, we consistently imputed an *N* of 90% of the total reported sample size. Finally, we imputed minor allele frequencies (MAF) for those summary statistics in which such information was unavailable, using the UK Biobank as reference data.

### Filtering of summary statistics

Insertions and deletions (indels) were excluded from analyses since reference data may not contain indels, and multiple summary statistics did not contain indels. To assure fair and interpretable comparisons across the 12 PDs, we filtered all summary statistics to include only those SNPs that were available for all 12 PDs (see **Supplementary Note 2**). After filtering, 3,857,404 SNPs remained, representing a substantial drop in the number of SNPs for most PDs (**Supplementary Table 1**). For additional information on the rationale behind this filtering step, and a detailed comparison of (the results of) filtered and unfiltered summary statistics, see **Supplementary Note 2**.

### Estimating SNP-based heritability and genome-wide genetic correlation

We used Linkage Disequilibrium Score regression (LDSC)^11^ to estimate the SNP-based heritability 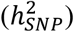 of each of the 12 PDs, representing the proportion of phenotypic variance that is explained by all common genetic variants included in the analysis. Heritability estimates on the liability scale were obtained by including estimates of population and study prevalence of each disorder. Precomputed LD scores were used that were calculated using European samples in the 1000 Genomes data (see **URLs**). Additionally, stratified LDSC^23^ was used to examine whether any of the 61 functional genomic categories was enriched for SNPs that contribute to the *h*^*2*^_*SNP*_ of the 12 PDs. Enrichment was defined as the proportion of *h*^*2*^_*SNP*_ in a given category divided by the proportion of SNPs in that category.

Bivariate LDSC^9^ was used to compute genetic correlations (*r*_*g*_’s) between the 12 PDs (cross-trait) as well as between unfiltered and filtered versions of the PD summary statistics (within-disorder). Bivariate LDSC, like single-trait LDSC, only requires summary statistics, and produces estimates that are unbiased by sample overlap.

### Genomic structural equation modelling and confirmatory factor analysis

Genomic structural equation modelling (SEM^12^) utilizes a multivariable extension of LDSC and combines the genetic signal from GWASs on multiple traits with their genetic correlation to estimate a genetic covariance matrix. The diagonal of the genetic covariance matrix contains the SNP-based heritability for each trait, while the off-diagonal terms reflect genetic covariances. In a second matrix, the information regarding estimation variability is stored (i.e., squared standard errors of genetic variances (diagonal) and covariance (off-diagonal)). Based on these two matrices, genomic SEM estimates the parameters of any user-specified genomic factor model without requiring individual-level data, or producing inaccurate results due to sample overlap.

Using confirmatory factor analysis (CFA), we fitted the four-factor model as reported by Grotzinger et al.^8^ as 11 out of our 12 PDs overlapped with this study. Based on the results from an exploratory factor analysis, the additional trait, INS, was hypothesized to load on the neurodevelopmental factor.

### Genetic correlations to 1,376 external traits using Complex Trait Genetics Virtual Lab

Through the Complex Trait Genetics Virtual Lab (CTG-VL)^13^ website (see **URLs**), we computed genetic correlations between the 12 PDs and a total of 1,618 external traits. Genetic correlation in CTG-VL is based on LDSC regression (see section above). Through manual filtering we assessed each trait and filtered out a total of 242 traits that either were duplicates or unrelated traits, such as ‘Job codes’ and ‘Leisure/social activities’. The remaining 1,376 traits were manually assigned to 7 main categories: *cognition, drug, medical, medicine, personal, physical*, and *psychologically*. The genetic correlations of individual traits within each category were then plotted across all PDs and Euclidean clustering was applied to the individual traits to visualize (dis)concordance in *r*_*g*_-patterns between PDs.

### Cross-trait meta-analysis using mvGWAMA

To scrutinize the shared variance across all 12 PDs, we conducted a cross-trait meta-analysis (CTMA) using mvGWAMA^14^ (see **URLs**), a freely available python script that can be used to conduct GWAS meta-analysis when there is sample overlap. Rather than requiring details on sample overlap and phenotypic correlations across samples, mvGWAMA uses the cross-trait LD score regression intercept to estimate sample overlap. For details we refer to the original publication^14^.

### Genomic risk loci and functional annotation of GWAS results

FUMA^15^ (see **URLs**) is an online platform that annotates SNPs to their biological functionality and maps implicated genes. Biological functionality is characterized for lead SNPs and SNPs in LD using potential regulatory functions (RegulomeDB score), deleteriousness score (CADD score), effects on gene functions (using ANNOVAR), and mRNA expression levels (using eQTL and chromatin interaction data).

Prior to annotation, FUMA first defines *independent significant SNPs* which have a genome-wide significant *p*-value (5×10^−8^) and are independent at *r*^*2*^<0.6. Based on LD information from UK Biobank genotypes, a subset of these *independent significant SNPs* is labeled as *lead SNPs* (independent from each other at *r*^*2*^<0.1). If LD blocks of *lead SNPs* are less than 250 kb apart, then they are merged. We annotated all SNPs in LD with the most significant SNP to get insight into the possible biological reasons for observing a statistical association, i.e., annotation was performed for all SNPs that were in LD (*r*^*2*^>0.6) with one of the *independent significant SNPs*, had a *p*-value lower than 1 × 10^−5^, and a minor allele frequency (MAF) > 0.0001.

### Mapping SNPs to genes

SNPs were mapped to genes using four different approaches. Using FUMA, SNPs were mapped to genes based on 1) their physical position in the genome, 2) eQTL associations, and 3) indirect genome interactions based on chromatin mapping (for details see Watanabe et al., 2017^15^). In addition, we conducted a genome-wide gene-based association study (GWGAS) through MAGMA^16^ (see **URLs**). MAGMA yields gene-based *p*-values through evaluation of the joint association effect of all SNPs within a gene, while accounting for LD between SNPs. The SNP locations were defined in reference to the human genome Build 37 (GRCh37/hg19), and only protein-coding genes (17,287) were included in the analysis. These gene-mapping analyses were conducted on the summary statistics of all 12 PDs and on the cross-trait meta-analysis summary statistics.

### Local genetic correlation analysis

LAVA (Local Analysis of [co]Variant Association; see **URLs**)^17^ is a tool developed to identify genomic regions that drive the genetic correlation between traits. Using LAVA, we first tested for univariate local genetic signal within each PD (i.e., the local 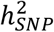). Regions that showed univariate signal for >1 PD were selected to test for local genetic correlation. To this end we used a threshold of *p* < 1×10^−4^, so as to filter out clearly non-associated loci (or loci clearly devoid of any univariate heritability) while retaining a large proportion of ‘sub-GWS’ loci that nonetheless may contribute to the overall 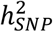, in order to get a more global overview of the shared genetic signal. This resulted in a total of 2,532 conducted bivariate tests as several tests will be conducted within the same region when multiple PDs show univariate signal in the same region. LAVA accounts for correlated SNPs due to LD by converting marginal SNP effects into their joint effects, based on an external LD reference. The 1000 Genomes (v3) data was used as the LD reference panel and regions were constructed by partitioning the genome into 2,495 semi-independent blocks of ∼1Mb. LAVA uses the intercept from bivariate LDSC to account for potential sample overlap. *P*-values of the local genetic correlations were corrected for the total number of bivariate tests conducted (threshold: *p* = 0.05 / no. of bivariate tests = 0.05 / 2,532 = 1.97×10^−5^).

### Gene-set and gene-property analysis

Through MAGMA’s gene-set and gene-property analysis, we tested 9,508 gene sets (canonical pathways and biological processes sets from MSigDB^18^; C2: CP & C5: BP), 53 tissue types (GTEx v8)^19^, and 565 cell types from Dropviz level2^20^. We applied multiple testing correction by combining the gene sets, tissue types and cell types (0.05 / (9,508 + 53 + 565)), and additionally also indicate which results survive additional correction for the number of PDs (0.05 / (9,508 + 53 + 565) / 12).

### SynGO and paralog annotation follow-up of mapped genes across all PDs

We conducted follow-up gene-set enrichment tests and annotations on all the mapped genes identified by FUMA (see above) for each PD separately, and compared annotations across PDs. To explore potential overlap, we did not correct for multiple testing when testing for significant gene-set enrichment. SynGO^22^ (see **URLs**) features 3,242 expert-curated synaptic annotations against 1,233 human genes that are grouped into 293 nested synaptic processes and cellular compartments. A user-defined gene-list can be inputted to SynGO to determine whether any of the genes in the gene-list has synaptic annotations and whether any synaptic processes or cellular compartments are significantly enriched. The enrichment test uses a Fisher’s exact test to determine whether the input gene-list shows higher than expected enrichment in any synaptic gene-set compared to a background reference of GTEx v7 brain expressed genes. Figures can be downloaded to illustrate enriched gene-sets and visualize potential overlap between multiple gene-list inputs.

The Gene Cards database^35^ (see **URLs**) provides 21,663 paralog annotations in a total of 3,217 paralog families for all human genes. All mapped genes were annotated to their respective paralog family and paralog families with genes from more than two different PDs were retained. Known gene and paralog family functions were extracted from the Gene Cards database to interpret functional similarities between genes.

### Testing overlap of shared SNPs and genes across disorders

Fisher’s exact tests were conducted to test for overlap in SNP- and gene-based signal across the 12 PDs for a range of *p*-value thresholds. For SNP-level analyses, we first applied LD-based pruning to all summary statistics, retaining only independent SNPs (a subset of 10,000 UKB participants was used as a reference set; *r*^*2*^ = 0.8). Fisher’s exact tests on the gene-level included all genes. For more specific information on how this test was conducted we refer to the supplementary information of (Nagel et al., 2018)^36^.

## Data Availability

All data produced in the present study are available upon reasonable request to the authors.

## URLs

Precomputed LD scores: https://github.com/bulik/ldsc

FUMA GWAS platform: http://fuma.ctglab.nl/

MAGMA gene-based and gene property analysis: https://ctg.cncr.nl/software/magma

LAVA local genetic correlations: https://ctg.cncr.nl/software/lava

mvGWAMA and effective sample size calculation: https://github.com/Kyoko-wtnb/mvGWAMA

Functional categories for stratified heritability: https://alkesgroup.broadinstitute.org/LDSCORE/

Single-cell data used in cell-type analysis: http://dropviz.org/

Synaptic Gene Ontologies platform: https://www.syngoportal.org

Complex Trait Genetics Virtual Lab platform: https://view.genoma.io/

GeneCards database: https://www.genecards.org

BioRender: www.BioRender.com

## Acknowledgements

This work was partially supported by NWO Gravitation program BRAINSCAPES: A Roadmap from Neurogenetics to Neurobiology (NWO: 024.004.012). The analyses were carried out on the Genetic Cluster Computer, which is financed by the Netherlands Scientific Organization (NWO: 480-05-003), by the VU University, Amsterdam, the Netherlands, and by the Dutch Brain Foundation, and is hosted by the Dutch National Computing and Networking Services SurfSARA. This research has been conducted using the UK Biobank resource under application number 16406. We thank the numerous participants, researchers, and staff from many studies who collected and contributed to the data. In particular, we!d like to express our gratitude to all MVP and UKB participants that have been so generous to share their data for analysis.

Figure 2 was created with BioRender.com. This figure was, in part, adapted from the “Central Dogma”, “Expression of ACE2 Receptor in Human Host Tissues” and “Notch Signaling Pathway” templates, by BioRender.com (2020). Retrieved from https://app.biorender.com/biorender-templates.

## Author Contributions

S.vd.S conceived the study. C.R. and M.N. performed the main analyses. J.W. and C.d.L were consulted on LAVA analyses and results. J.G., M.B.S, D.L. and R.P. conducted the original study on PTSD and provided early access to the PTSD summary statistics. C.R., M.N. and S.vd.S. wrote the paper. All authors discussed the results and commented on the paper.

