## Supplementary figures for "Exploring the genetic overlap between 12 psychiatric disorders"

Romero, C. et al.

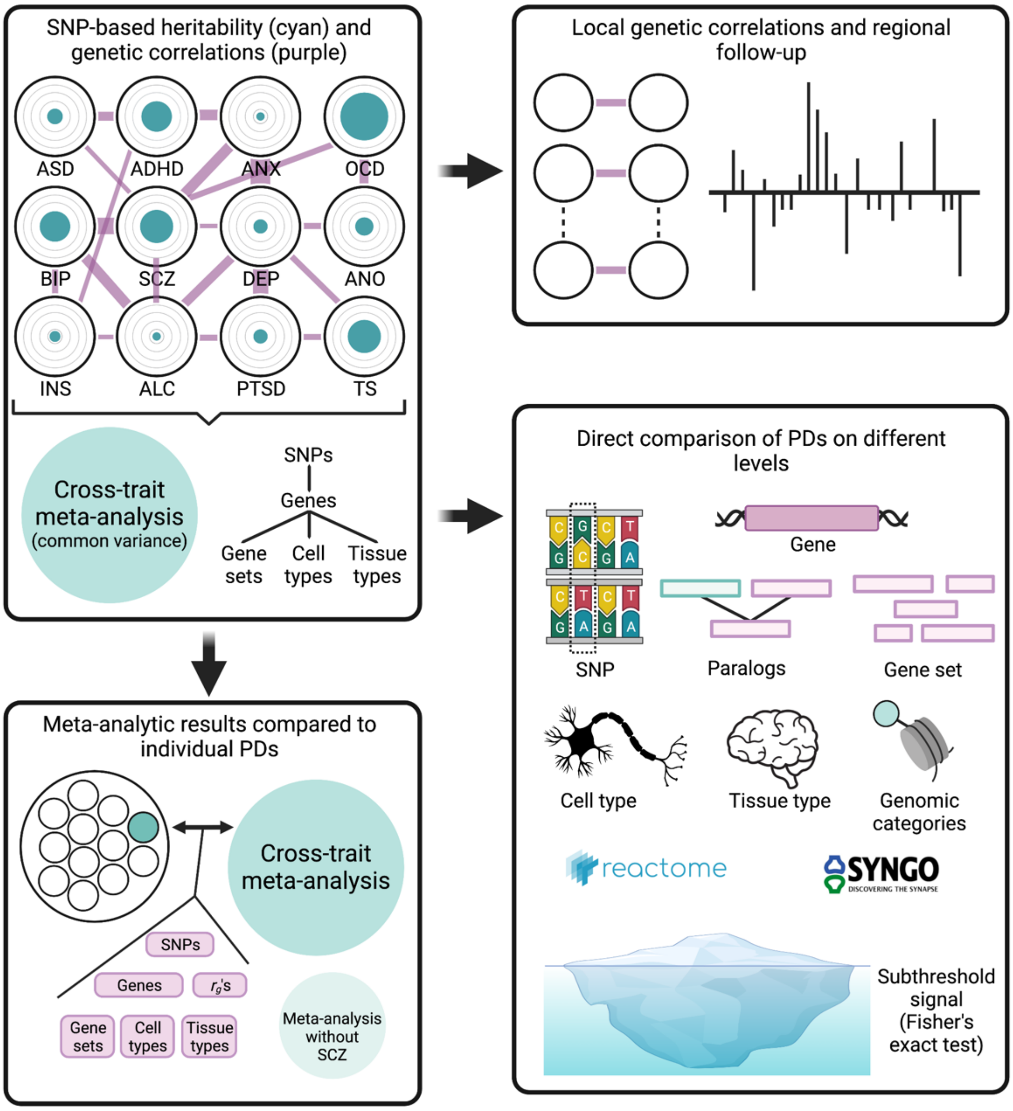

**Supplementary Fig. 1. Flow chart of analyses performed in the current study**

Schematic representation of analyses preformed in the current study. GWAS results from 12 PDs were obtained from several sources. SNP-based heritability and genetic correlations were estimated before PDs were combined in a cross-trait meta-analysis. SNP and gene results from the meta-analysis were then compared to results of individual PDs. A second meta-analysis was conducted without inclusion of SCZ to assess the effect of excluding the most statistically powered phenotype. Comparisons between individual PDs were then made on the SNP-, gene-, and gene follow-up level in addition to comparing subthreshold and local genetic correlation signal.

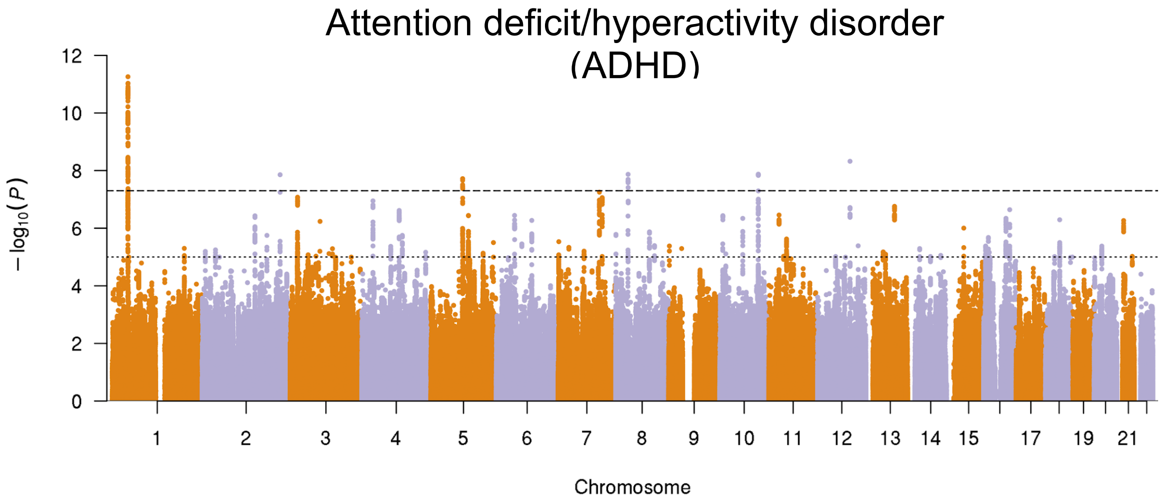
Attention deficit/hyperactivity disorder (ADHD)
−log10(P)
Chromosome

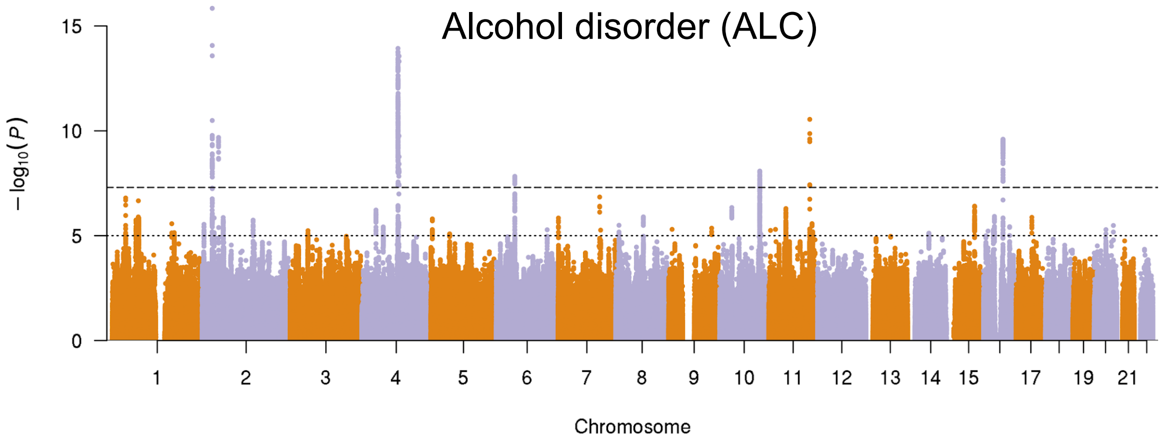
Alcohol disorder (ALC)
−log10(P)
Chromosome

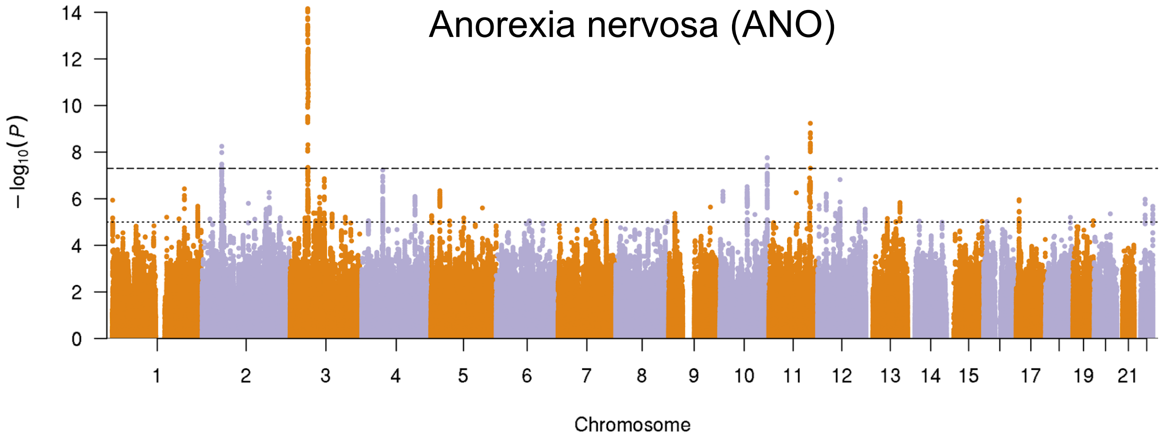
Anorexia nervosa (ANO)
−log10(P)
Chromosome

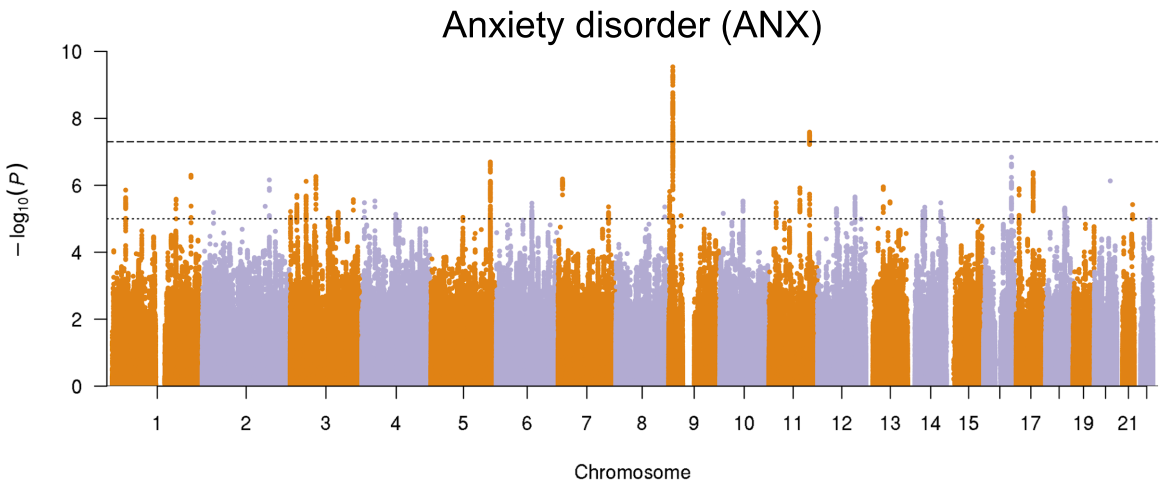
Anxiety disorder (ANX)
$-\log_{10}(P)$
0 2 4 6 8 10
1 2 3 4 5 6 7 8 9 10 11 12 13 14 15 17 19 21
Chromosome

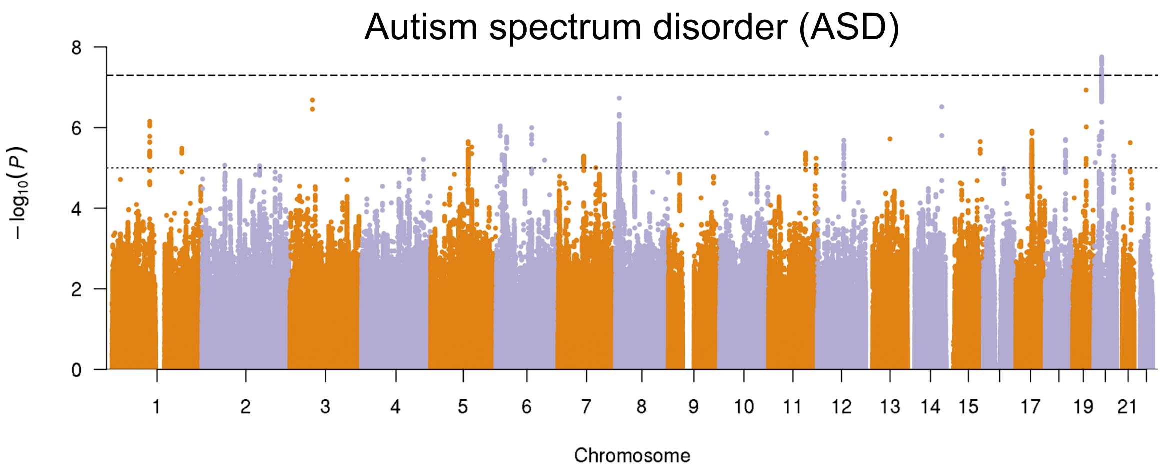
Autism spectrum disorder (ASD)
$-\log_{10}(P)$
0 2 4 6 8
1 2 3 4 5 6 7 8 9 10 11 12 13 14 15 17 19 21
Chromosome

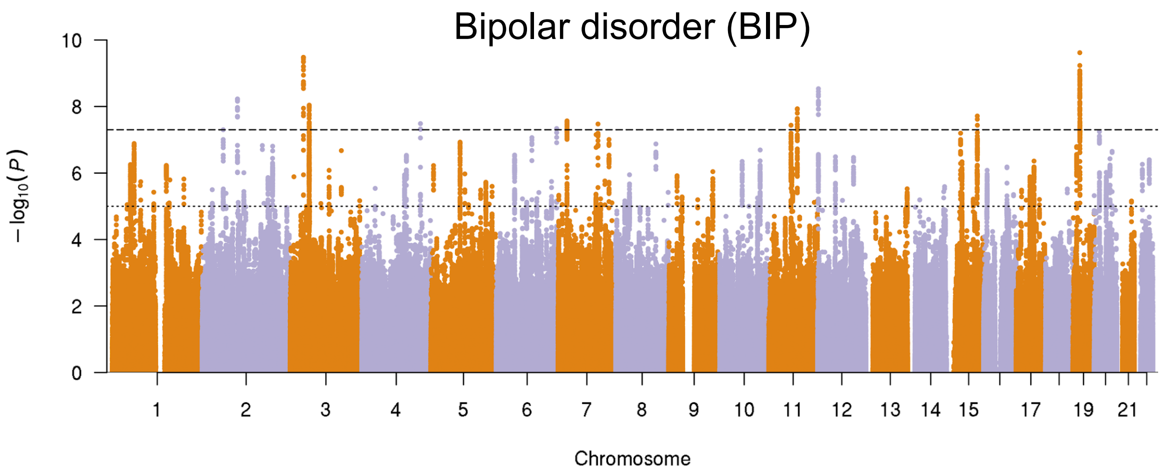
Bipolar disorder (BIP)
$-\log_{10}(P)$
0 2 4 6 8 10
1 2 3 4 5 6 7 8 9 10 11 12 13 14 15 17 19 21
Chromosome

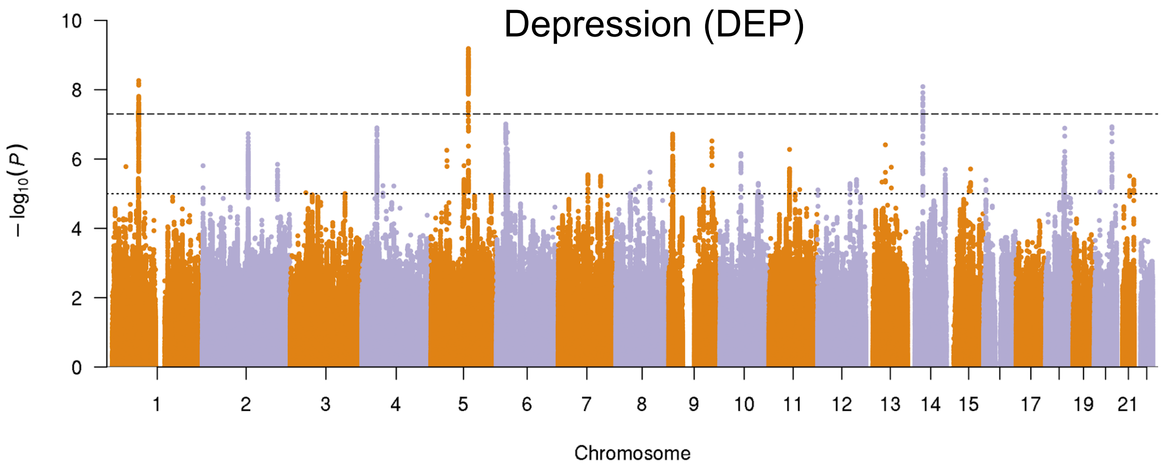
Depression (DEP)
10
8
6
4
2
0
− log10(P)
17
19
21
Chromosome

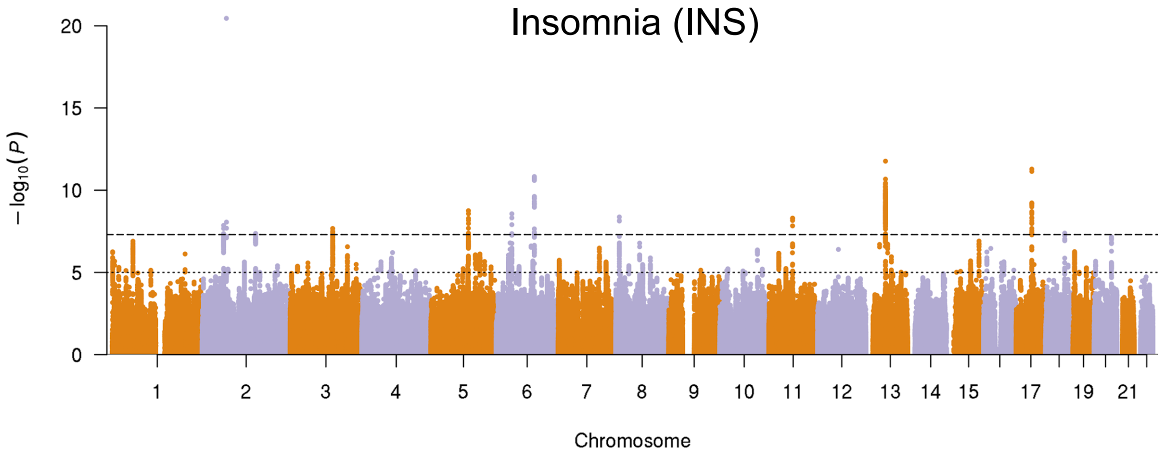
Insomnia (INS)
20
15
10
5
0
− log10(P)
17
19
21
Chromosome

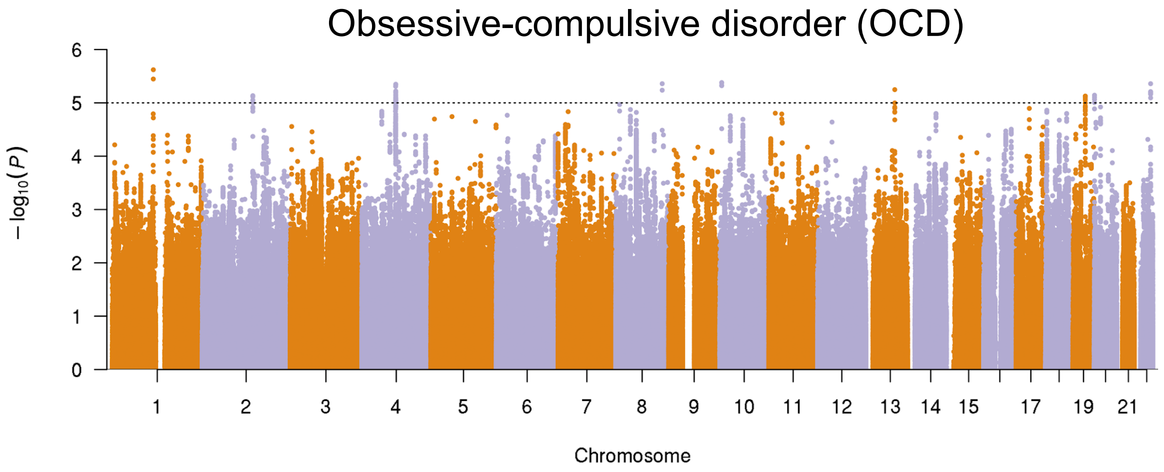
Obsessive-compulsive disorder (OCD)
6
5
4
3
2
1
0
− log10(P)
17
19
21
Chromosome

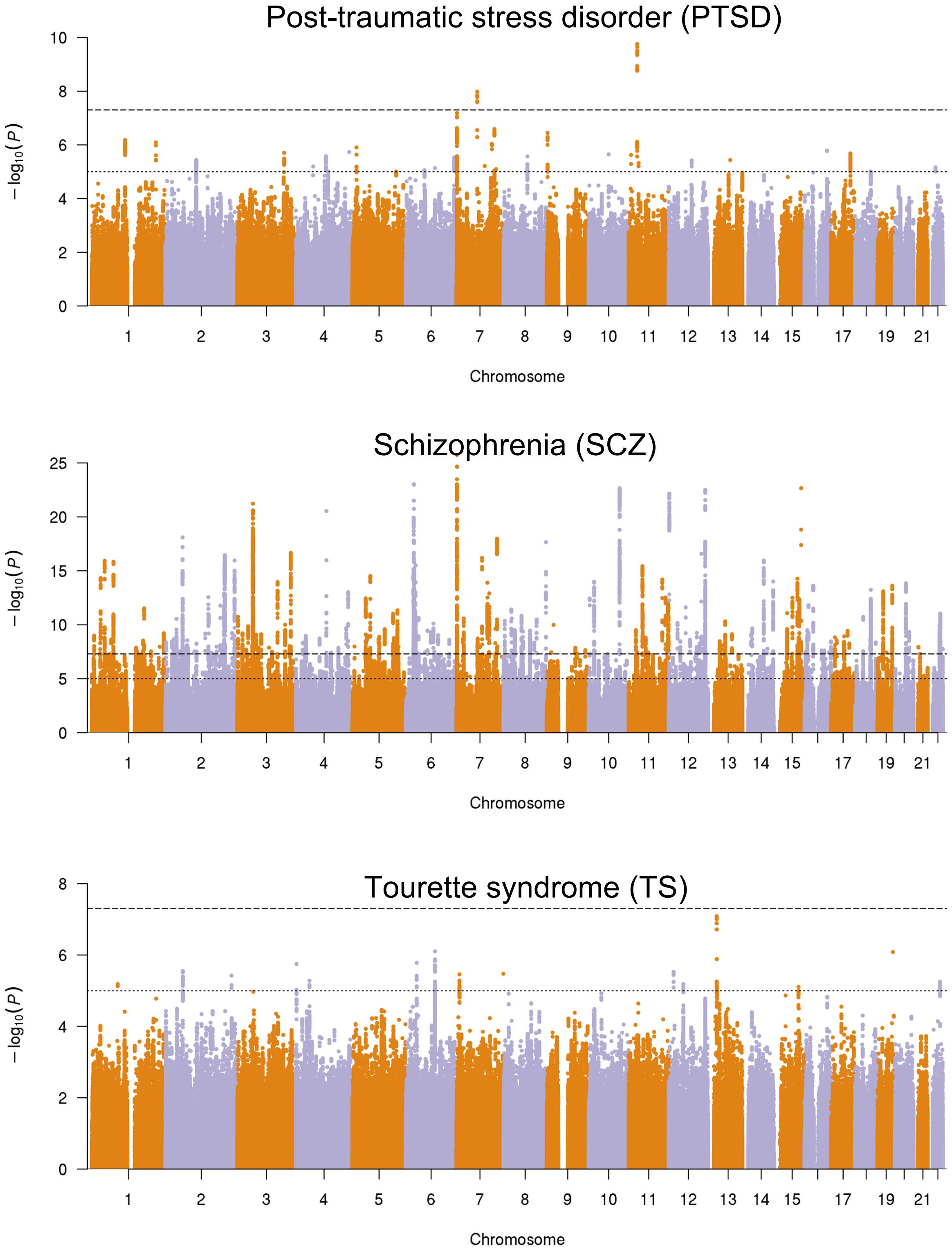

**Supplementary Fig. 2. Manhattan plots of SNP-based associations for 12 PDs**

SNP association results from all the PDs included in the study (in alphabetic order). Genomic position is shown on the x-axis and –log10 transformed SNP *p*-values are plotted on the y-axis. Dashed lines indicate GWS ($p < 5x10^{-8}$) and dotted lines indicate the 'suggestive' significance threshold ($p < 5x10^{-5}$). Only SNPs shared across all summary statistics were included.

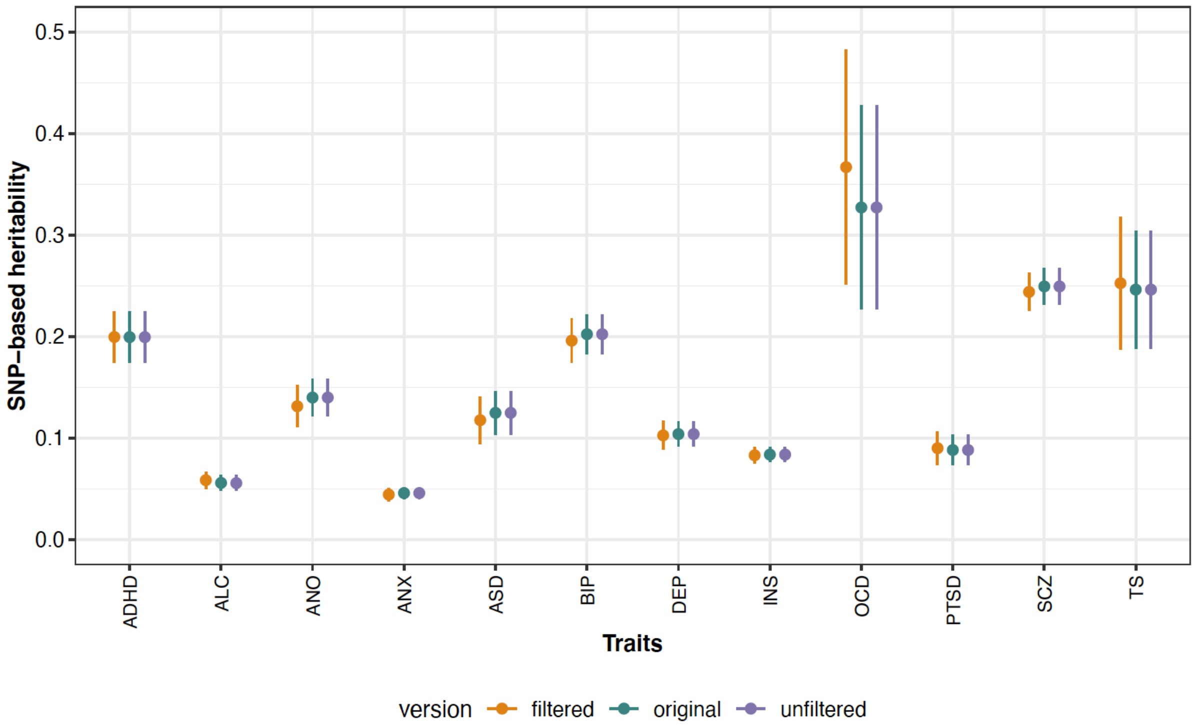

**Supplementary Fig. 3. SNP-based heritability estimates for the original, unfiltered, and filtered versions of the summary statistics of all 12 PDs**

SNP-based heritability estimated using LDSC for all PDs show little to no sign of variation between original, unfiltered, and filtered versions. Y-axis displays the proportion of total phenotypic variation that is attributable to common genetic variants (lines representing error-bar of estimate).

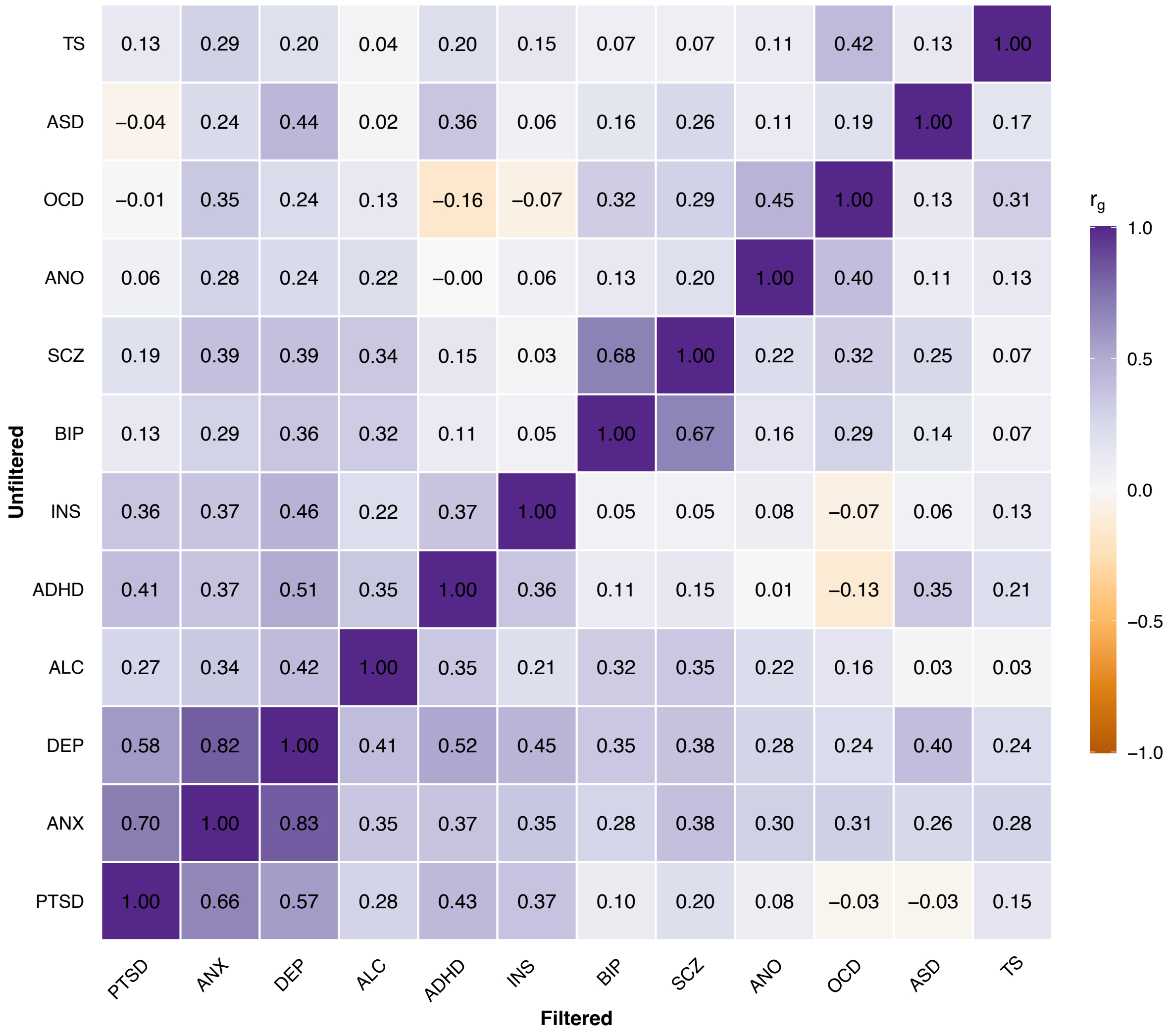

**Supplementary Fig. 4. Heatmap of cross-trait $r_g$'s within and between the unfiltered and the filtered summary statistics of the 12 PDs**

Genetic correlations were computed using bivariate LDSC. The bottom triangle features $r_g$'s between the PDs using the filtered summary statistics, whereas the top triangle features $r_g$'s between the PDs using the unfiltered summary statistics. Overall, the PDs show low to moderate genetic correlation, and patterns are very similar when using the unfiltered versus the filtered summary statistics.

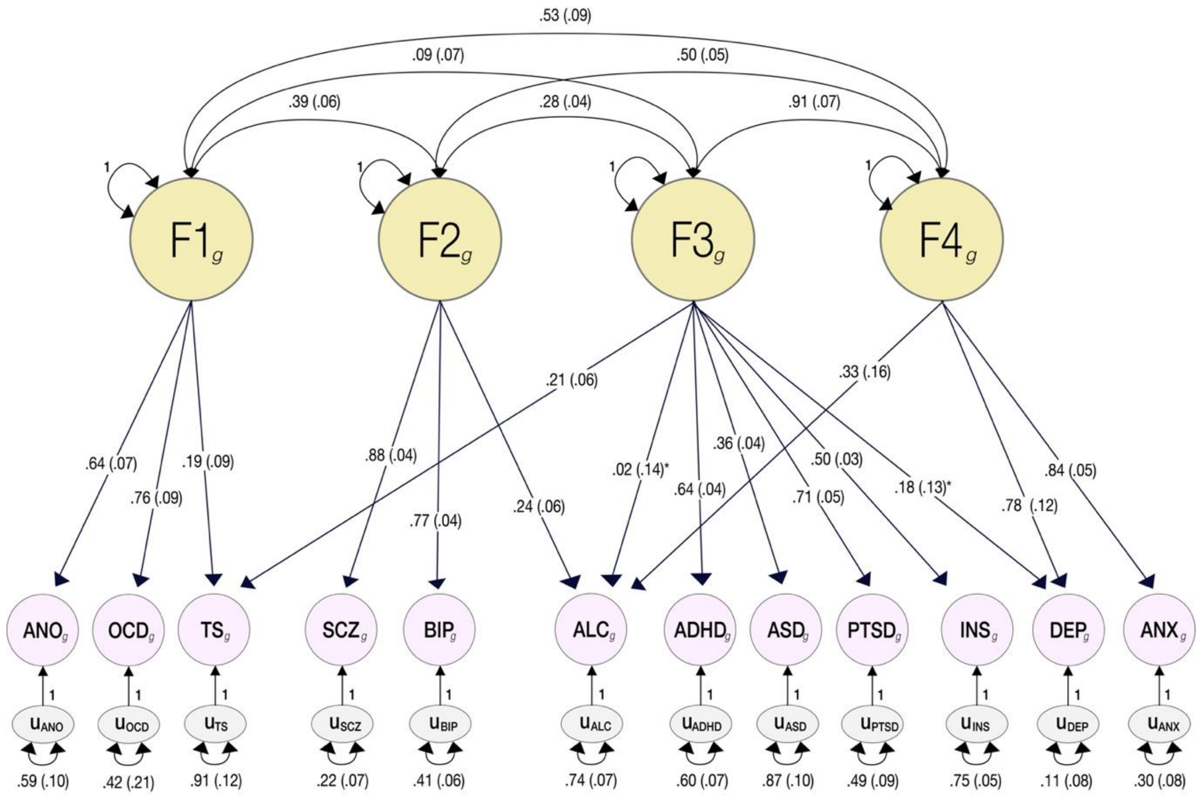

**Supplementary Fig. 5. CFA plot (gSEM)**

The model was adopted from Grotzinger et al.[1] as 11 out of 12 traits overlapped with this study. Based on the global genetic correlations, the additional trait, INS, was hypothesized to load onto the neurodevelopmental and internalizing factor. A CFA model identical to Grotzinger et al. with the additional loadings of INS on the neurodevelopmental factor had good fit indices (AIC = 316, CFI = 0.93, SRMR = 0.074). Displayed estimates have been standardized. Estimates denoted with * are not significant at P = 0.05.

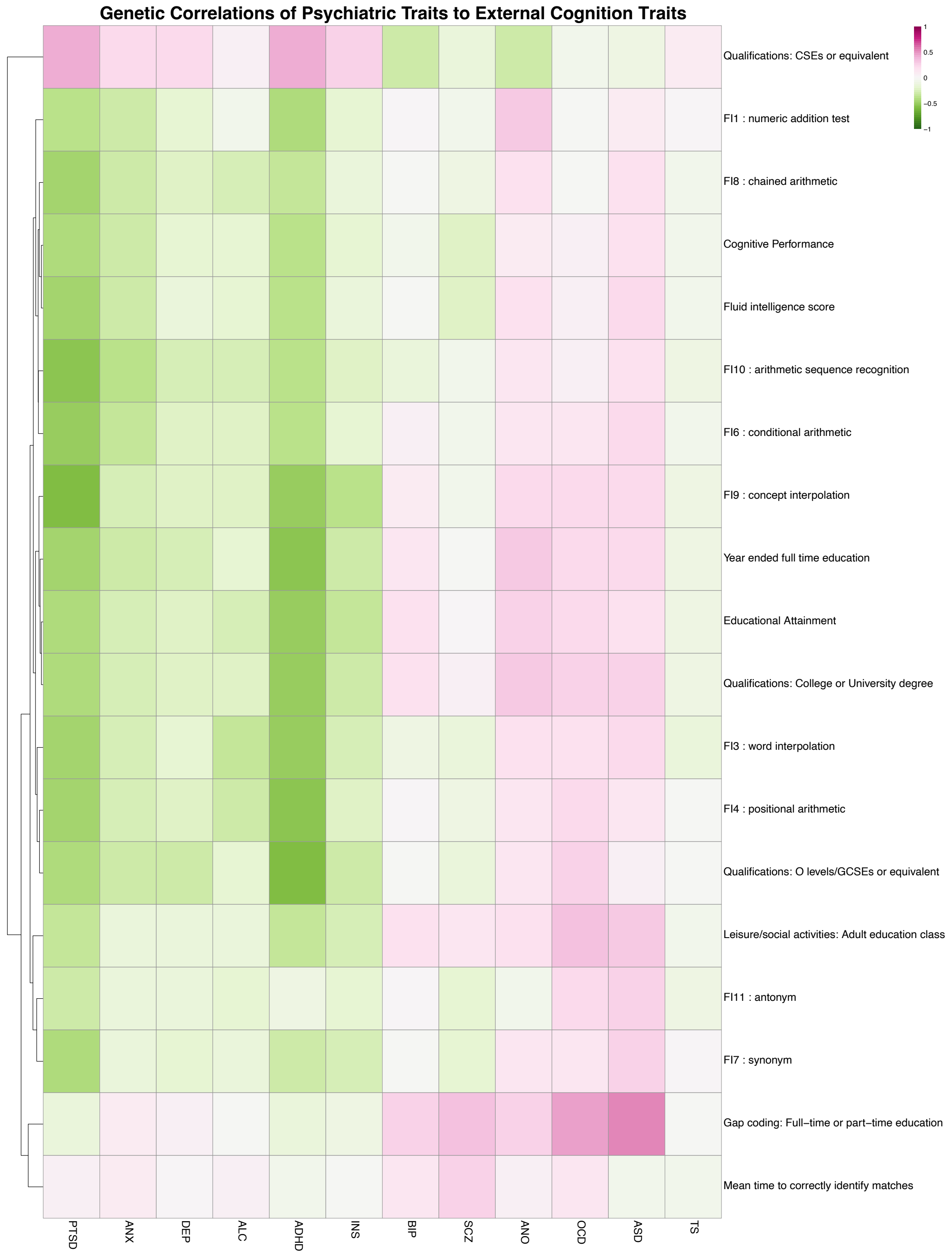

**Supplementary Fig. 6. Genetic correlations of the 12 PDs with cognition-related traits**

Genetic correlations computed across 1,376 external traits through CTG-VL show large degree of concordance in direction of correlation coefficient across multiple PDs. The 1,376 external traits were manually inspected and clustered to predominant categories of traits, resulting in cognitive-, drug-, medically-, medicine-, personal-, physical-, and psychologically-related trait categories, with respective correlation heatmaps to each. Heatmaps display individual traits in the y-axis and all 12 PDs on the x-axis. Positive and negative genetic correlations are denoted by purple and green color, respectively.

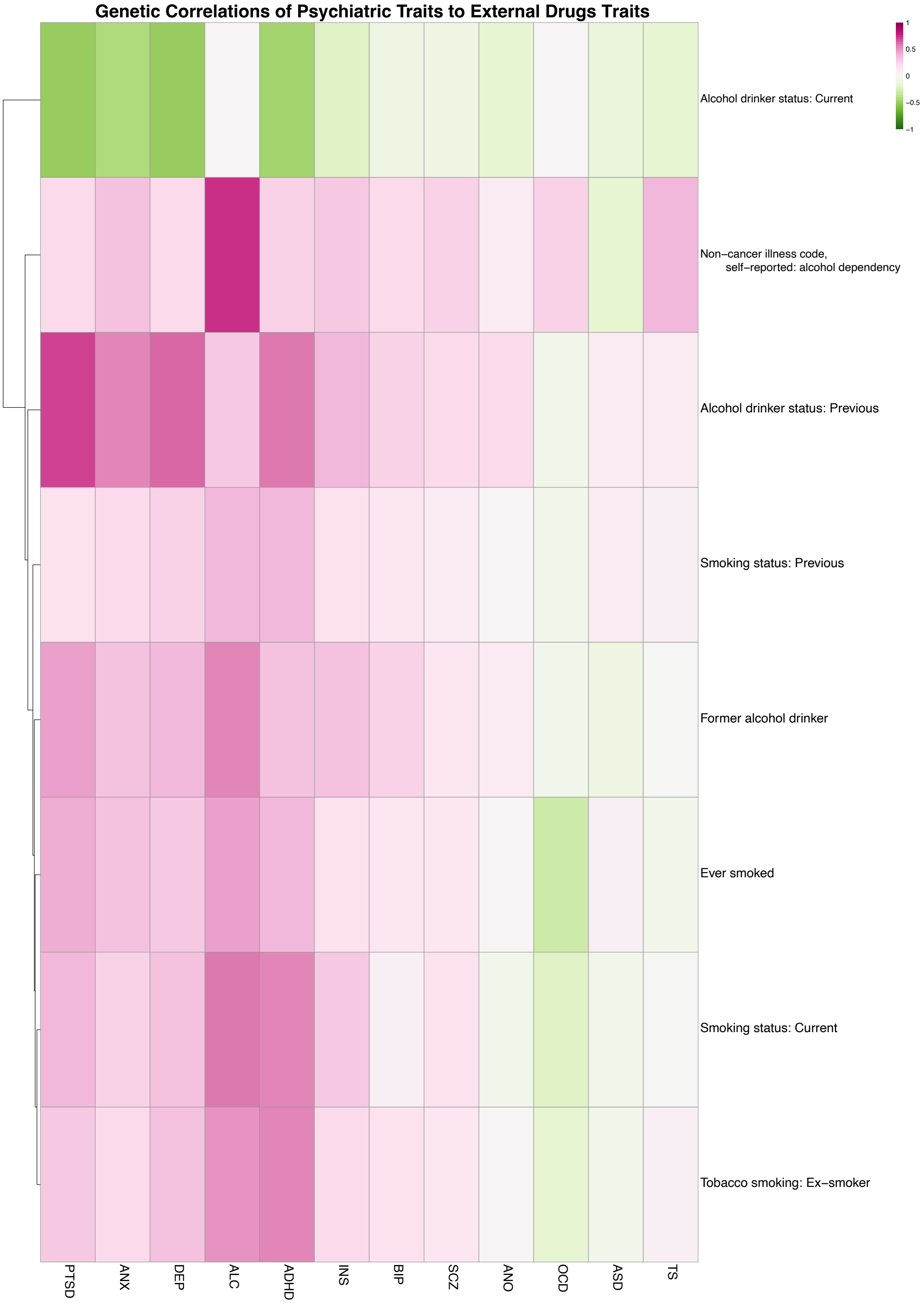

**Supplementary Fig. 7. Genetic correlations of the 12 PDs with drug-related traits**

Genetic correlations computed across 1,376 external traits through CTG-VL show large degree of concordance in direction of correlation coefficient across multiple PDs. The 1,376 external traits were manually inspected and clustered to predominant categories of traits, resulting in cognitive-, drug-, medically-, medicine-, personal-, physical-, and psychologically-related trait categories, with respective correlation heatmaps to each. Heatmaps display individual traits in the y-axis and all 12 PDs on the x-axis. Positive and negative genetic correlations are denoted by purple and green color, respectively.

### Genetic Correlations of Psychiatric Traits to External Medical Traits

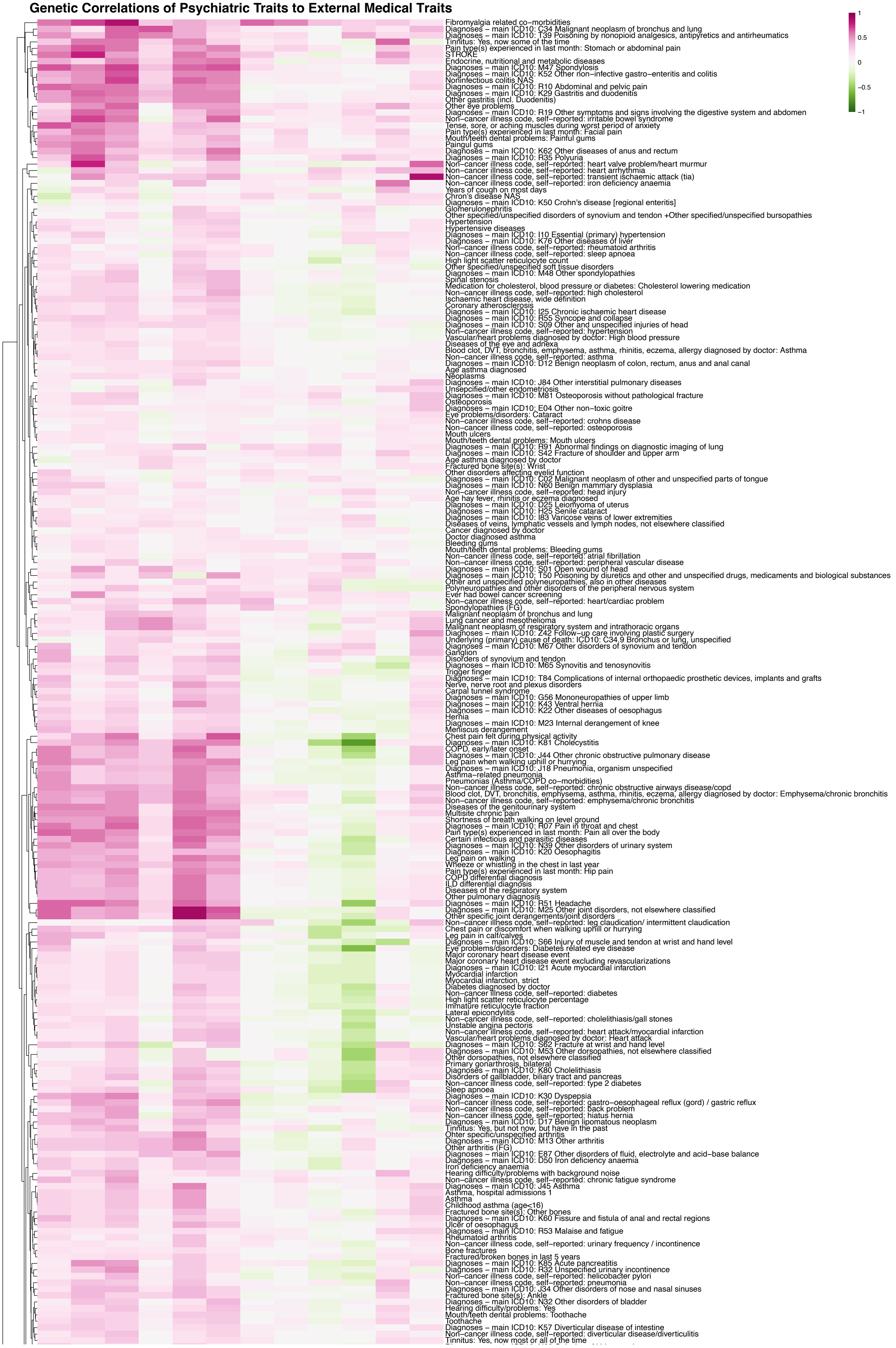

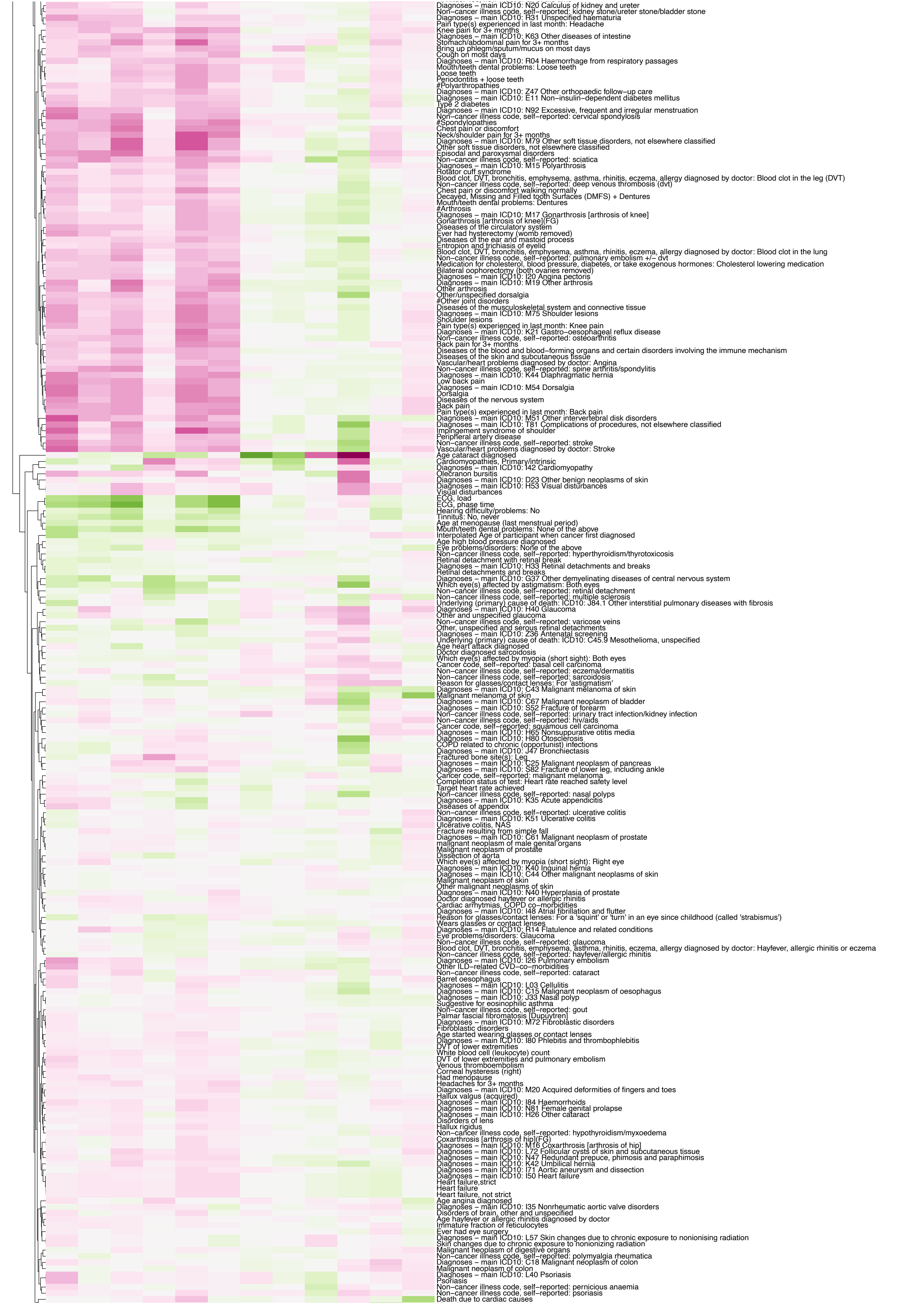

Diagnoses – main ICD10: N20 Calculus of kidney and ureter
Non–cancer illness code, self–reported: kidney stone/ureter stone/bladder stone
Diagnoses – main ICD10: R31 Unspecified haematuria
Pain type(s) experienced in last month: Headache
Knee pain for 3+ months
Diagnoses – main ICD10: K63 Other diseases of intestine
Stomach/abdominal pain for 3+ months
Bring up phlegm/sputum/mucus on most days
Cough on most days
Diagnoses – main ICD10: R04 Haemorrhage from respiratory passages
Mouth/teeth dental problems: Loose teeth
Loose teeth
Periodontitis + loose teeth
#Polyarthropathies
Diagnoses – main ICD10: Z47 Other orthopaedic follow–up care
Diagnoses – main ICD10: E11 Non–insulin–dependent diabetes mellitus
Type 2 diabetes
Diagnoses – main ICD10: N92 Excessive, frequent and irregular menstruation
Non–cancer illness code, self–reported: cervical spondylosis
#Spondylopathies
Chest pain or discomfort
Neck/shoulder pain for 3+ months
Diagnoses – main ICD10: M79 Other soft tissue disorders, not elsewhere classified
Other soft tissue disorders, not elsewhere classified
Episodal and paroxysmal disorders
Non–cancer illness code, self–reported: sciatica
Diagnoses – main ICD10: M15 Polyarthrosis
Rotator cuff syndrome
Blood clot, DVT, bronchitis, emphysema, asthma, rhinitis, eczema, allergy diagnosed by doctor: Blood clot in the leg (DVT)
Non–cancer illness code, self–reported: deep venous thrombosis (dvt)
Chest pain or discomfort walking normally
Decayed, Missing and Filled tooth Surfaces (DMFS) + Dentures
Mouth/teeth dental problems: Dentures
#Arthrosis
Diagnoses – main ICD10: M17 Gonarthrosis [arthrosis of knee]
Gonarthrosis [arthrosis of knee](FG)
Diseases of the circulatory system
Ever had hysterectomy (womb removed)
Diseases of the ear and mastoid process
Entropion and trichiasis of eyelid
Blood clot, DVT, bronchitis, emphysema, asthma, rhinitis, eczema, allergy diagnosed by doctor: Blood clot in the lung
Non–cancer illness code, self–reported: pulmonary embolism +/– dvt
Medication for cholesterol, blood pressure, diabetes, or take exogenous hormones: Cholesterol lowering medication
Bilateral oophorectomy (both ovaries removed)
Diagnoses – main ICD10: I20 Angina pectoris
Diagnoses – main ICD10: M19 Other arthrosis
Other arthrosis
Other/unspecified dorsalgia
#Other joint disorders
Diseases of the musculoskeletal system and connective tissue
Diagnoses – main ICD10: M75 Shoulder lesions
Shoulder lesions
Pain type(s) experienced in last month: Knee pain
Diagnoses – main ICD10: K21 Gastro–oesophageal reflux disease
Non–cancer illness code, self–reported: osteoarthritis
Back pain for 3+ months
Diseases of the blood and blood–forming organs and certain disorders involving the immune mechanism
Diseases of the skin and subcutaneous tissue
Vascular/heart problems diagnosed by doctor: Angina
Non–cancer illness code, self–reported: spine arthritis/spondylitis
Diagnoses – main ICD10: K44 Diaphragmatic hernia
Low back pain
Diagnoses – main ICD10: M54 Dorsalgia
Dorsalgia
Diseases of the nervous system
Back pain
Pain type(s) experienced in last month: Back pain
Diagnoses – main ICD10: M51 Other intervertebral disk disorders
Diagnoses – main ICD10: T81 Complications of procedures, not elsewhere classified
Impingement syndrome of shoulder
Peripheral artery disease
Non–cancer illness code, self–reported: stroke
Vascular/heart problems diagnosed by doctor: Stroke
Age cataract diagnosed
Cardiomyopathies, Primary/intrinsic
Diagnoses – main ICD10: I42 Cardiomyopathy
Olecranon bursitis
Diagnoses – main ICD10: D23 Other benign neoplasms of skin
Diagnoses – main ICD10: H53 Visual disturbances
Visual disturbances
ECG, load
ECG, phase time
Hearing difficulty/problems: No
Tinnitus: No, never
Age at menopause (last menstrual period)
Mouth/teeth dental problems: None of the above
Interpolated Age of participant when cancer first diagnosed
Age high blood pressure diagnosed
Eye problems/disorders: None of the above
Non–cancer illness code, self–reported: hyperthyroidism/thyrotoxicosis
Retinal detachment with retinal break
Diagnoses – main ICD10: H33 Retinal detachments and breaks
Retinal detachments and breaks
Diagnoses – main ICD10: G37 Other demyelinating diseases of central nervous system
Which eye(s) affected by astigmatism: Both eyes
Non–cancer illness code, self–reported: retinal detachment
Non–cancer illness code, self–reported: multiple sclerosis
Underlying (primary) cause of death: ICD10: J84.1 Other interstitial pulmonary diseases with fibrosis
Diagnoses – main ICD10: H40 Glaucoma
Other and unspecified glaucoma
Non–cancer illness code, self–reported: varicose veins
Other, unspecified and serous retinal detachments
Diagnoses – main ICD10: Z36 Antenatal screening
Underlying (primary) cause of death: ICD10: C45.9 Mesothelioma, unspecified
Age heart attack diagnosed
Doctor diagnosed sarcoidosis
Which eye(s) affected by myopia (short sight): Both eyes
Cancer code, self–reported: basal cell carcinoma
Non–cancer illness code, self–reported: eczema/dermatitis
Non–cancer illness code, self–reported: sarcoidosis
Reason for glasses/contact lenses: For 'astigmatism'
Diagnoses – main ICD10: C43 Malignant melanoma of skin
Malignant melanoma of skin
Diagnoses – main ICD10: C67 Malignant neoplasm of bladder
Diagnoses – main ICD10: S52 Fracture of forearm
Non–cancer illness code, self–reported: urinary tract infection/kidney infection
Non–cancer illness code, self–reported: hiv/aids
Cancer code, self–reported: squamous cell carcinoma
Diagnoses – main ICD10: H65 Nonsuppurative otitis media
Diagnoses – main ICD10: H80 Otosclerosis
COPD related to chronic (opportunist) infections
Diagnoses – main ICD10: J47 Bronchiectasis
Fractured bone site(s): Leg
Diagnoses – main ICD10: C25 Malignant neoplasm of pancreas
Diagnoses – main ICD10: S82 Fracture of lower leg, including ankle
Cancer code, self–reported: malignant melanoma
Completion status of test: Heart rate reached safety level
Target heart rate achieved
Non–cancer illness code, self–reported: nasal polyps
Diagnoses – main ICD10: K35 Acute appendicitis
Diseases of appendix
Non–cancer illness code, self–reported: ulcerative colitis
Diagnoses – main ICD10: K51 Ulcerative colitis
Ulcerative colitis, NAS
Fracture resulting from simple fall
Diagnoses – main ICD10: C61 Malignant neoplasm of prostate
malignant neoplasm of male genital organs
Malignant neoplasm of prostate
Dissection of aorta
Which eye(s) affected by myopia (short sight): Right eye
Diagnoses – main ICD10: K40 Inguinal hernia
Diagnoses – main ICD10: C44 Other malignant neoplasms of skin
Malignant neoplasm of skin
Other malignant neoplasms of skin
Diagnoses – main ICD10: N40 Hyperplasia of prostate
Doctor diagnosed hayfever or allergic rhinitis
Cardiac arrhytmias, COPD co–morbidities
Diagnoses – main ICD10: I48 Atrial fibrillation and flutter
Reason for glasses/contact lenses: For a 'squint' or 'turn' in an eye since childhood (called 'strabismus')
Wears glasses or contact lenses
Diagnoses – main ICD10: R14 Flatulence and related conditions
Eye problems/disorders: Glaucoma
Non–cancer illness code, self–reported: glaucoma
Blood clot, DVT, bronchitis, emphysema, asthma, rhinitis, eczema, allergy diagnosed by doctor: Hayfever, allergic rhinitis or eczema
Non–cancer illness code, self–reported: hayfever/allergic rhinitis
Diagnoses – main ICD10: I26 Pulmonary embolism
Other ILD–related CVD–co–morbidities
Non–cancer illness code, self–reported: cataract
Barret oesophagus
Diagnoses – main ICD10: L03 Cellulitis
Diagnoses – main ICD10: C15 Malignant neoplasm of oesophagus
Diagnoses – main ICD10: J33 Nasal polyp
Suggestive for eosinophilic asthma
Non–cancer illness code, self–reported: gout
Palmar fascial fibromatosis [Dupuytren]
Diagnoses – main ICD10: M72 Fibroblastic disorders
Fibroblastic disorders
Age started wearing glasses or contact lenses
Diagnoses – main ICD10: I80 Phlebitis and thrombophlebitis
DVT of lower extremities
White blood cell (leukocyte) count
DVT of lower extremities and pulmonary embolism
Venous thromboembolism
Corneal hysteresis (right)
Had menopause
Headaches for 3+ months
Diagnoses – main ICD10: M20 Acquired deformities of fingers and toes
Hallux valgus (acquired)
Diagnoses – main ICD10: I84 Haemorrhoids
Diagnoses – main ICD10: N81 Female genital prolapse
Diagnoses – main ICD10: H26 Other cataract
Disorders of lens
Hallux rigidus
Non–cancer illness code, self–reported: hypothyroidism/myxoedema
Coxarthrosis [arthrosis of hip](FG)
Diagnoses – main ICD10: M16 Coxarthrosis [arthrosis of hip]
Diagnoses – main ICD10: L72 Follicular cysts of skin and subcutaneous tissue
Diagnoses – main ICD10: N47 Redundant prepuce, phimosis and paraphimosis
Diagnoses – main ICD10: K42 Umbilical hernia
Diagnoses – main ICD10: I71 Aortic aneurysm and dissection
Diagnoses – main ICD10: I50 Heart failure
Heart failure,strict
Heart failure
Heart failure, not strict
Age angina diagnosed
Diagnoses – main ICD10: I35 Nonrheumatic aortic valve disorders
Disorders of brain, other and unspecified
Age hayfever or allergic rhinitis diagnosed by doctor
Immature fraction of reticulocytes
Ever had eye surgery
Diagnoses – main ICD10: L57 Skin changes due to chronic exposure to nonionising radiation
Skin changes due to chronic exposure to nonionizing radiation
Malignant neoplasm of digestive organs
Non–cancer illness code, self–reported: polymyalgia rheumatica
Diagnoses – main ICD10: C18 Malignant neoplasm of colon
Malignant neoplasm of colon
Diagnoses – main ICD10: L40 Psoriasis
Psoriasis
Non–cancer illness code, self–reported: pernicious anaemia
Non–cancer illness code, self–reported: psoriasis
Death due to cardiac causes

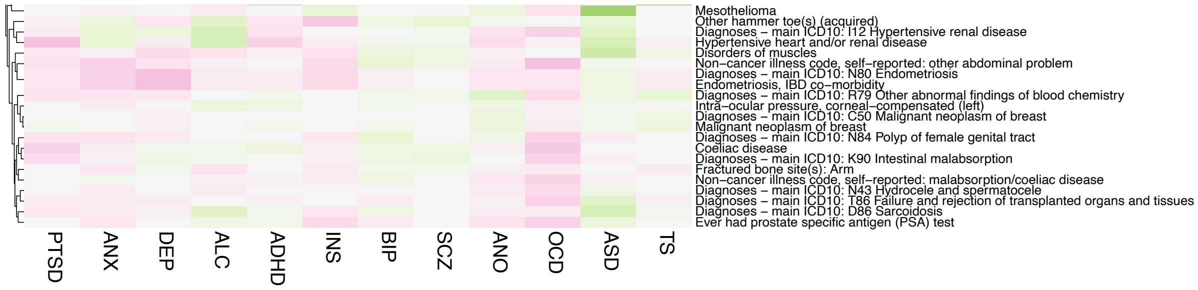

**Supplementary Fig. 8. Genetic correlations of the 12 PDs with medical traits**

Genetic correlations computed across 1,376 external traits through CTG-VL show large degree of concordance in direction of correlation coefficient across multiple PDs. The 1,376 external traits were manually inspected and clustered to predominant categories of traits, resulting in cognitive-, drug-, medically-, medicine-, personal-, physical-, and psychologically-related trait categories, with respective correlation heatmaps to each. Heatmaps display individual traits in the y-axis and all 12 PDs on the x-axis. Positive and negative genetic correlations are denoted by purple and green color, respectively.

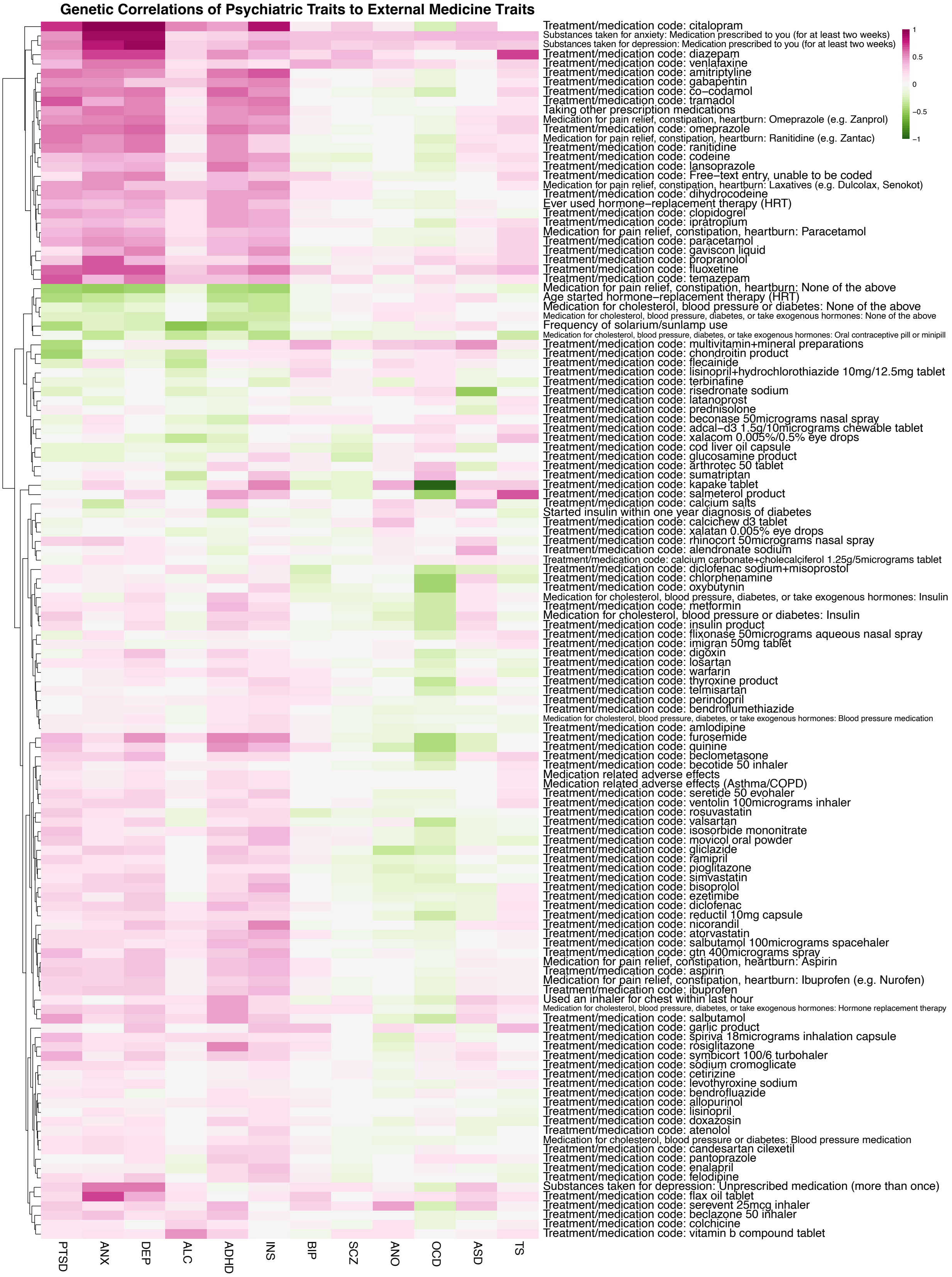

**Supplementary Fig. 9. Genetic correlations of the 12 PDs with medicine-use traits**

Genetic correlations computed across 1,376 external traits through CTG-VL show large degree of concordance in direction of correlation coefficient across multiple PDs. The 1,376 external traits were manually inspected and clustered to predominant categories of traits, resulting in cognitive-, drug-, medically-, medicine-, personal-, physical-, and psychologically-related trait categories, with respective correlation heatmaps to each. Heatmaps display individual traits in the y-axis and all 12 PDs on the x-axis. Positive and negative genetic correlations are denoted by purple and green color, respectively.

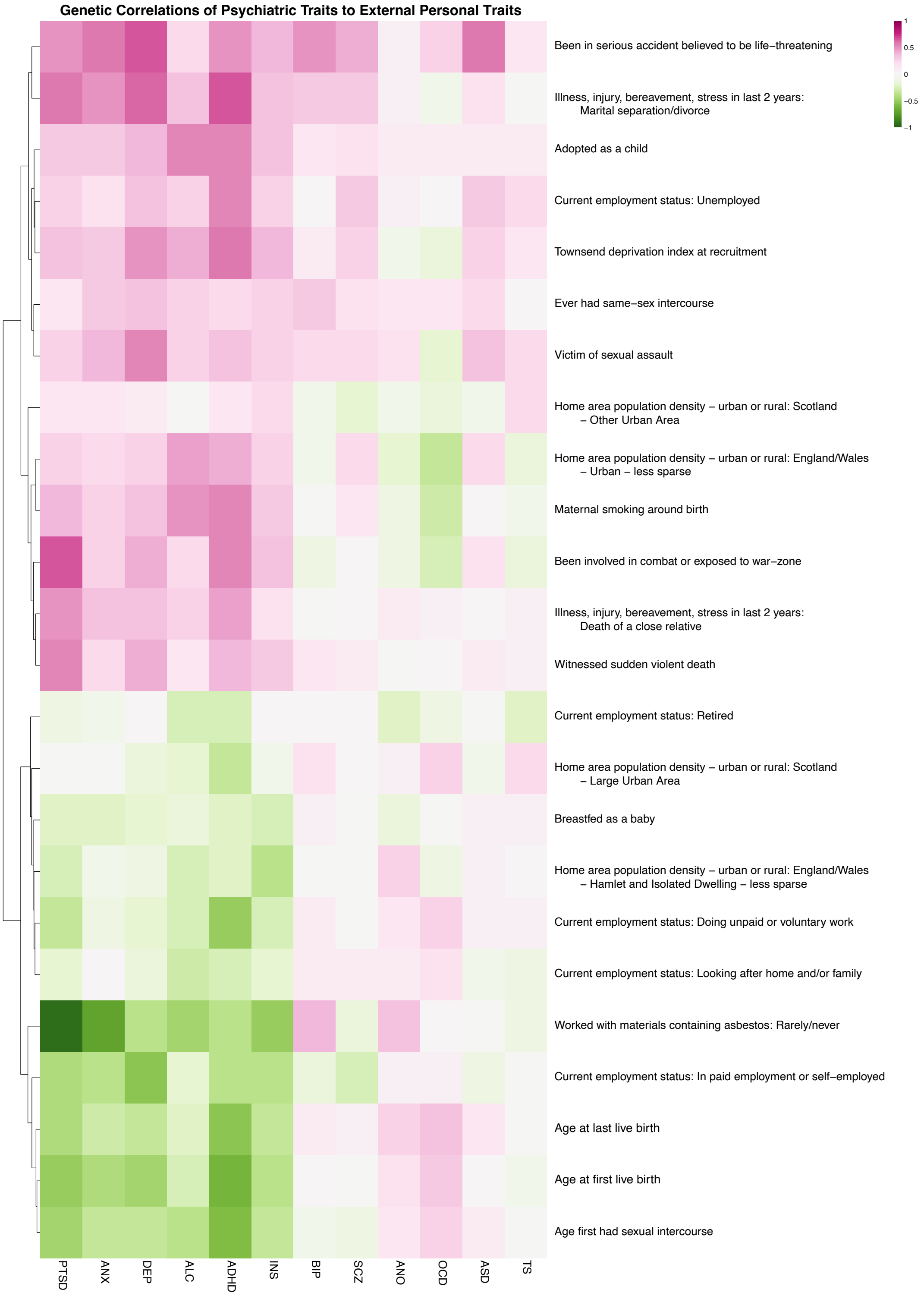

**Supplementary Fig. 10. Genetic correlations of the 12 PDs with personal traits**

Genetic correlations computed across 1,376 external traits through CTG-VL show large degree of concordance in direction of correlation coefficient across multiple PDs. The 1,376 external traits were manually inspected and clustered to predominant categories of traits, resulting in cognitive-, drug-, medically-, medicine-, personal-, physical-, and psychologically-related trait categories, with respective correlation heatmaps to each. Heatmaps display individual traits in the y-axis and all 12 PDs on the x-axis. Positive and negative genetic correlations are denoted by purple and green color, respectively.

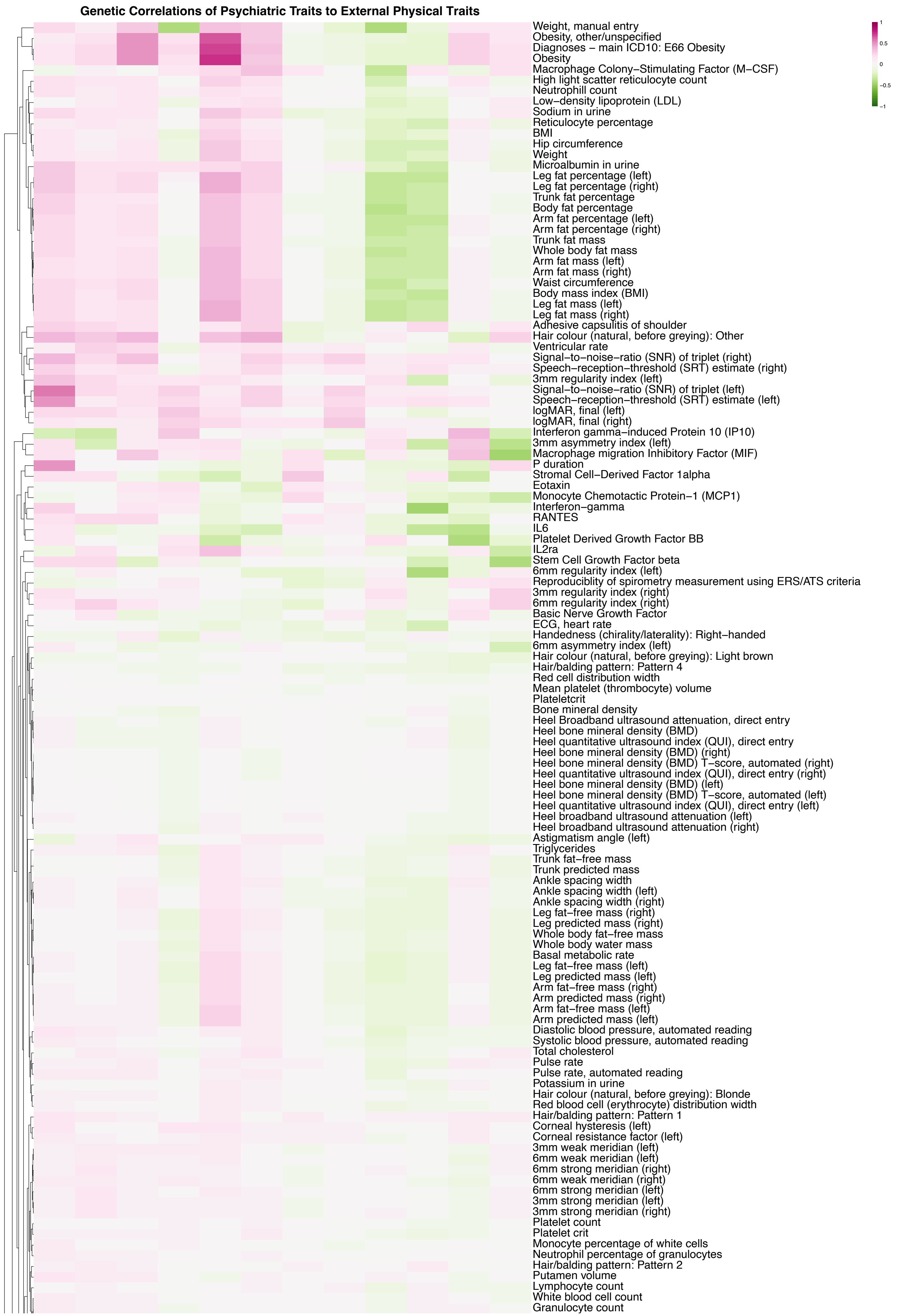
Genetic Correlations of Psychiatric Traits to External Physical Traits
Weight, manual entry
Obesity, other/unspecified
Diagnoses – main ICD10: E66 Obesity
Obesity
Macrophage Colony–Stimulating Factor (M–CSF)
High light scatter reticulocyte count
Neutrophill count
Low–density lipoprotein (LDL)
Sodium in urine
Reticulocyte percentage
BMI
Hip circumference
Weight
Microalbumin in urine
Leg fat percentage (left)
Leg fat percentage (right)
Trunk fat percentage
Body fat percentage
Arm fat percentage (left)
Arm fat percentage (right)
Trunk fat mass
Whole body fat mass
Arm fat mass (left)
Arm fat mass (right)
Waist circumference
Body mass index (BMI)
Leg fat mass (left)
Leg fat mass (right)
Adhesive capsulitis of shoulder
Hair colour (natural, before greying): Other
Ventricular rate
Signal–to–noise–ratio (SNR) of triplet (right)
Speech–reception–threshold (SRT) estimate (right)
3mm regularity index (left)
Signal–to–noise–ratio (SNR) of triplet (left)
Speech–reception–threshold (SRT) estimate (left)
logMAR, final (left)
logMAR, final (right)
Interferon gamma–induced Protein 10 (IP10)
3mm asymmetry index (left)
Macrophage migration Inhibitory Factor (MIF)
P duration
Stromal Cell–Derived Factor 1alpha
Eotaxin
Monocyte Chemotactic Protein–1 (MCP1)
Interferon–gamma
RANTES
IL6
Platelet Derived Growth Factor BB
IL2ra
Stem Cell Growth Factor beta
6mm regularity index (left)
Reproduciblity of spirometry measurement using ERS/ATS criteria
3mm regularity index (right)
6mm regularity index (right)
Basic Nerve Growth Factor
ECG, heart rate
Handedness (chirality/laterality): Right–handed
6mm asymmetry index (left)
Hair colour (natural, before greying): Light brown
Hair/balding pattern: Pattern 4
Red cell distribution width
Mean platelet (thrombocyte) volume
Plateletcrit
Bone mineral density
Heel Broadband ultrasound attenuation, direct entry
Heel bone mineral density (BMD)
Heel quantitative ultrasound index (QUI), direct entry
Heel bone mineral density (BMD) (right)
Heel bone mineral density (BMD) T–score, automated (right)
Heel quantitative ultrasound index (QUI), direct entry (right)
Heel bone mineral density (BMD) (left)
Heel bone mineral density (BMD) T–score, automated (left)
Heel quantitative ultrasound index (QUI), direct entry (left)
Heel broadband ultrasound attenuation (left)
Heel broadband ultrasound attenuation (right)
Astigmatism angle (left)
Triglycerides
Trunk fat–free mass
Trunk predicted mass
Ankle spacing width
Ankle spacing width (left)
Ankle spacing width (right)
Leg fat–free mass (right)
Leg predicted mass (right)
Whole body fat–free mass
Whole body water mass
Basal metabolic rate
Leg fat–free mass (left)
Leg predicted mass (left)
Arm fat–free mass (right)
Arm predicted mass (right)
Arm fat–free mass (left)
Arm predicted mass (left)
Diastolic blood pressure, automated reading
Systolic blood pressure, automated reading
Total cholesterol
Pulse rate
Pulse rate, automated reading
Potassium in urine
Hair colour (natural, before greying): Blonde
Red blood cell (erythrocyte) distribution width
Hair/balding pattern: Pattern 1
Corneal hysteresis (left)
Corneal resistance factor (left)
3mm weak meridian (left)
6mm weak meridian (left)
6mm strong meridian (right)
6mm weak meridian (right)
6mm strong meridian (left)
3mm strong meridian (left)
3mm strong meridian (right)
Platelet count
Platelet crit
Monocyte percentage of white cells
Neutrophil percentage of granulocytes
Hair/balding pattern: Pattern 2
Putamen volume
Lymphocyte count
White blood cell count
Granulocyte count
1
0.5
0
−0.5
−1

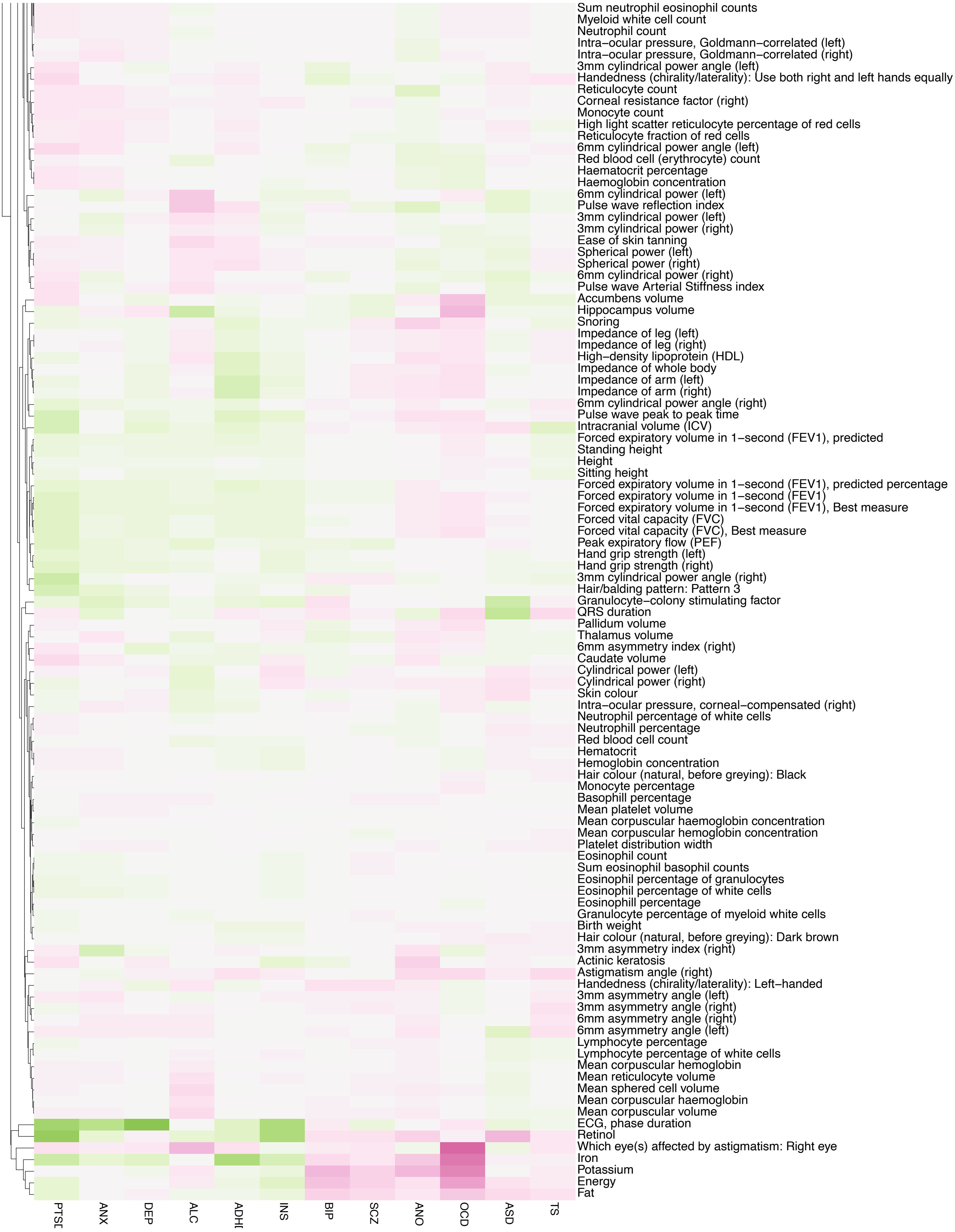

**Supplementary Fig. 11. Genetic correlations of the 12 PDs with physical traits**

Genetic correlations computed across 1,376 external traits through CTG-VL show large degree of concordance in direction of correlation coefficient across multiple PDs. The 1,376 external traits were manually inspected and clustered to predominant categories of traits, resulting in cognitive-, drug-, medically-, medicine-, personal-, physical-, and psychologically-related trait categories, with respective correlation heatmaps to each. Heatmaps display individual traits in the y-axis and all 12 PDs on the x-axis. Positive and negative genetic correlations are denoted by purple and green color, respectively.

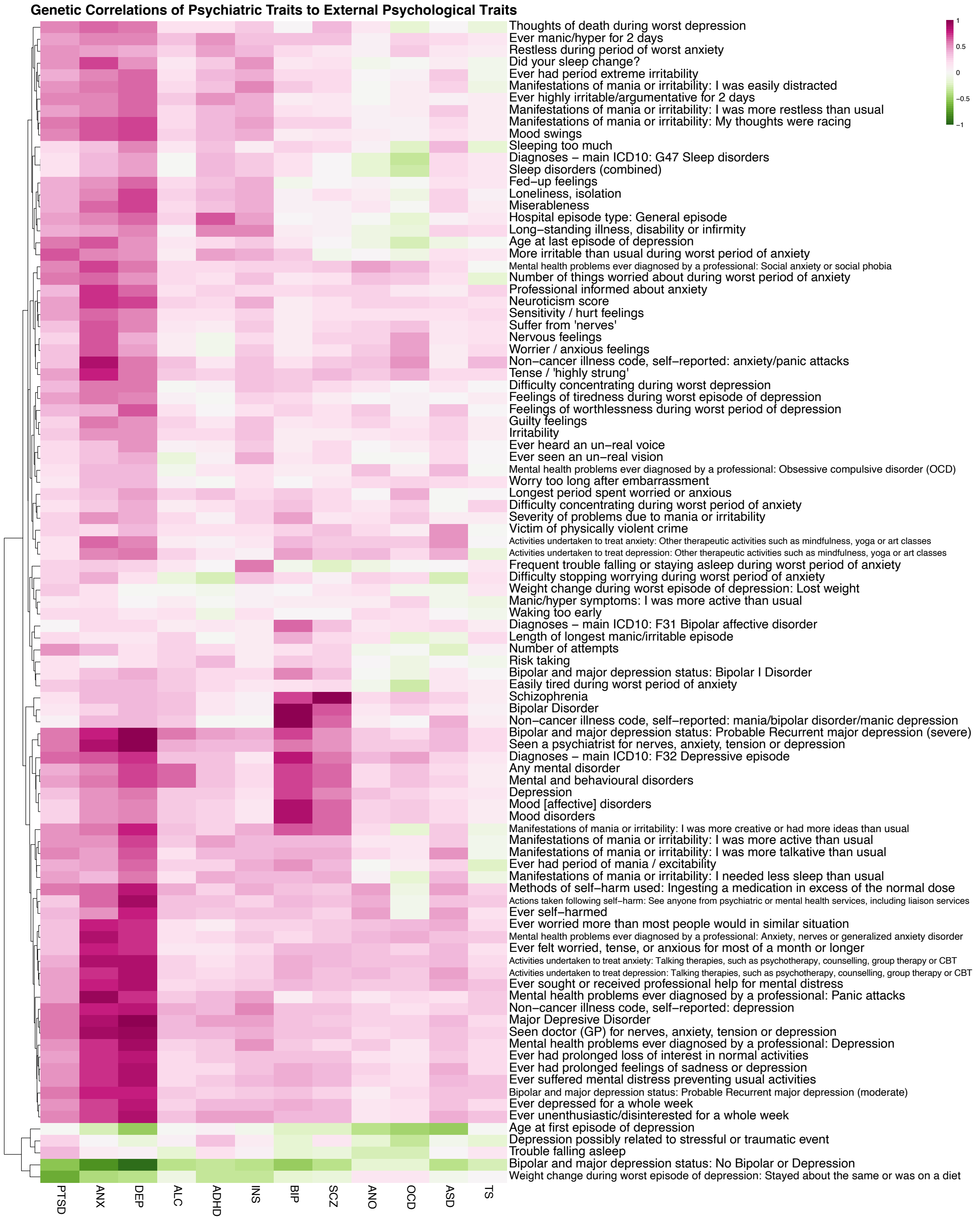

**Supplementary Fig. 12. Genetic correlations of the 12 PDs with psychological traits**

Genetic correlations computed across 1,376 external traits through CTG-VL show large degree of concordance in direction of correlation coefficient across multiple PDs. The 1,376 external traits were manually inspected and clustered to predominant categories of traits, resulting in cognitive-, drug-, medically-, medicine-, personal-, physical-, and psychologically-related trait categories, with respective correlation heatmaps to each. Heatmaps display individual traits in the y-axis and all 12 PDs on the x-axis. Positive and negative genetic correlations are denoted by purple and green color, respectively.

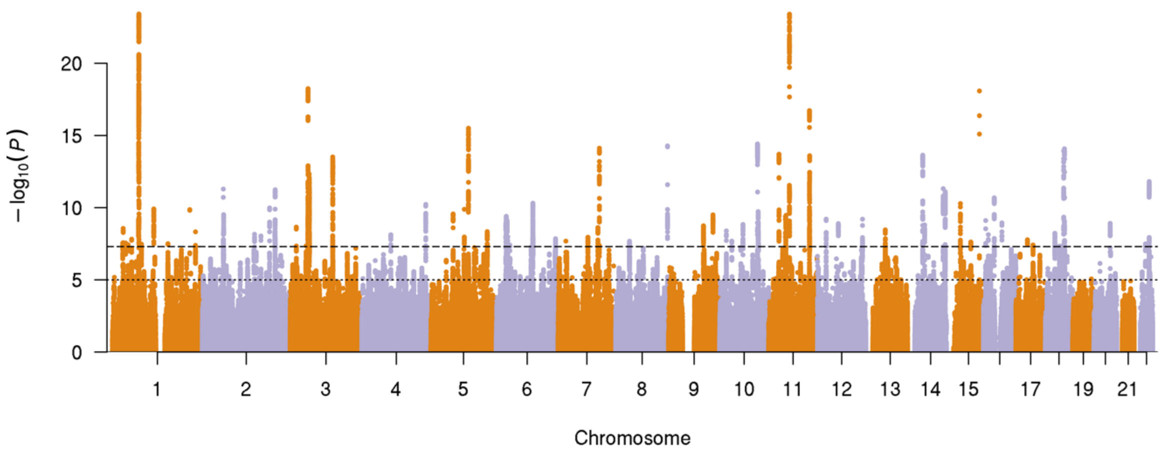

**Supplementary Fig. 13. Manhattan plot for cross-trait meta-analysis**

SNP association results from the cross-trait meta-analysis conducted on all 12 PDs included in the study (only using SNPs shared across all PD summary statistics). Genomic position is shown on the x-axis and -log10 transformed SNP *p*-values are plotted on the y-axis. Dashed lines indicate GWS ($p < 5x10^{-8}$) and dotted lines indicate the 'suggestive' significance threshold ($P < 5x10^{-5}$).

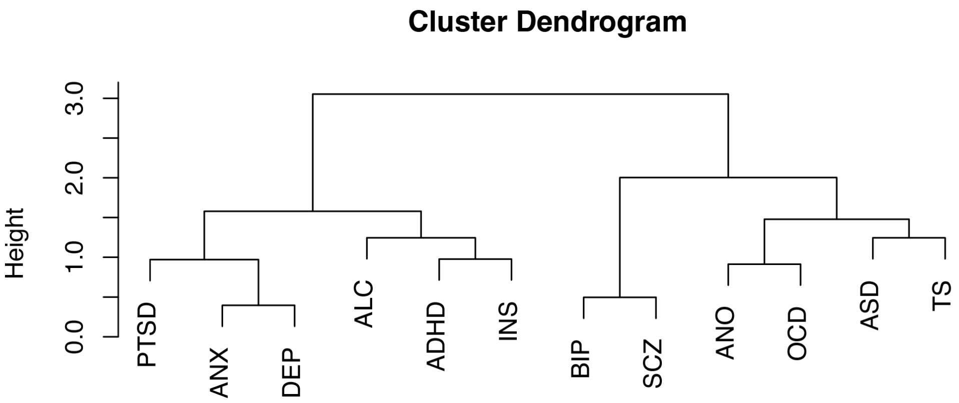

**Supplementary Fig. 14. Dendrogram displaying hierarchical clustering of genome-wide genetic correlations between the 12 PDs**

Cluster dendrogram of PDs displaying the results obtained with Ward's method of hierarchical clustering on the $r_g$'s among all 12 PDs.

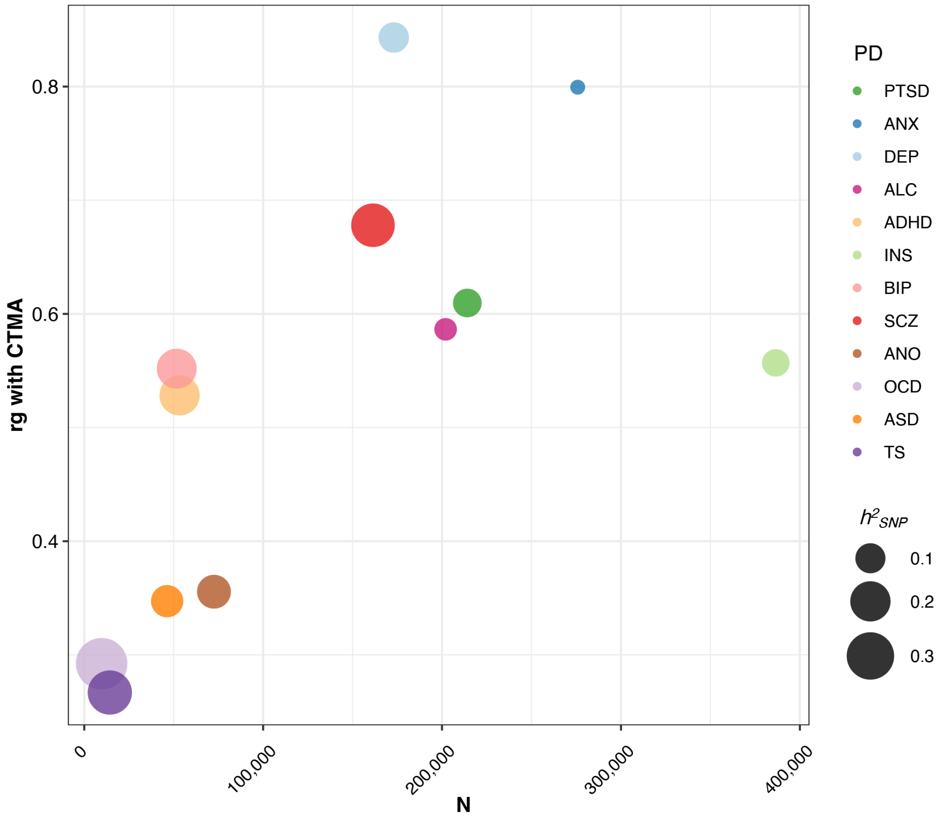

**Supplementary Fig. 15. Relationship between the sample size of individual PD summary statistics with the $r_g$ of the individual PDs with the CTMA.**

Sample size of the summary statistics of the 12 individual PDs (x-axis) plotted against the magnitude of their genetic correlation with the CTMA summary statistics (y-axis). The size of the dots reflects the SNP-based heritability. In general, we see higher genetic correlations with the CTMA for PDs for which the original GWASs were based on larger samples, i.e., for the most highly powered GWASs.

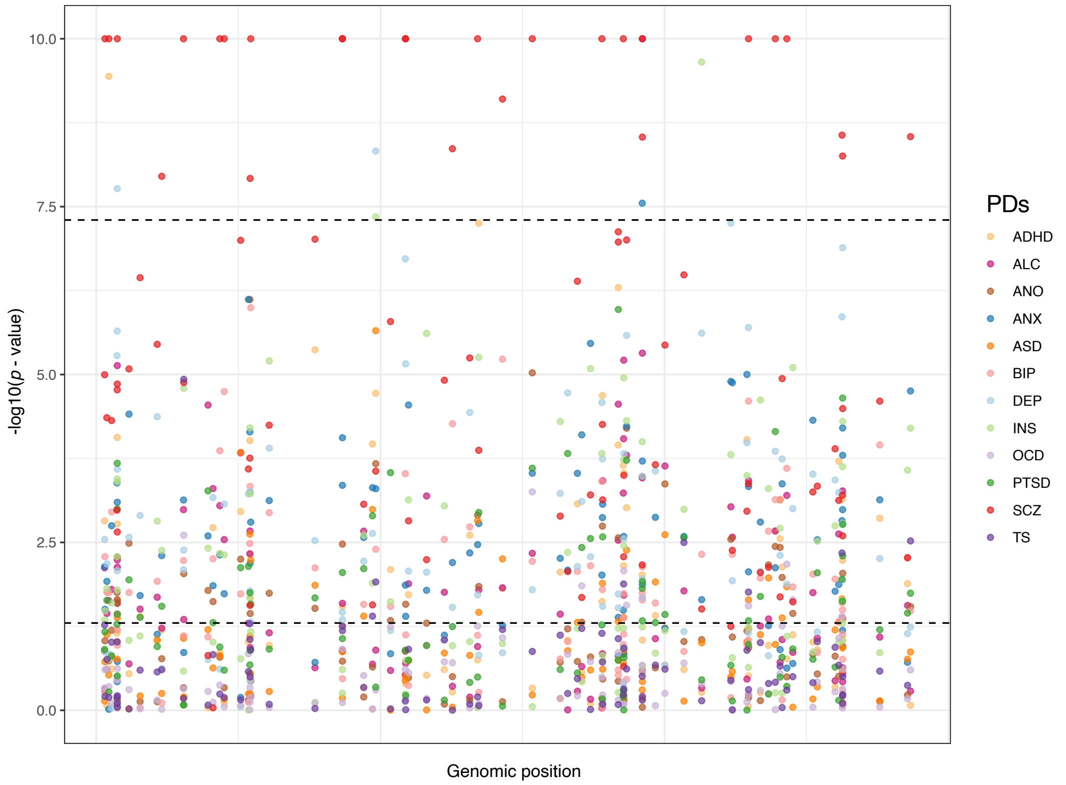

**Supplementary Fig. 16. Individual *p*-values for each PD for the 92 lead SNPs identified in the CTMA.**

*P*-values of the 12 individual PDs for the 92 lead SNPs identified in the cross-trait meta-analysis. Each dot represents a SNP, with colors corresponding to specific PDs (see legends). SNPs are ranked on the x-axis by order in the genome (i.e., distances between SNPs are not meaningful). The y-axis displays -log10 transformed *p*-values. The lower and higher dashed lines denote $p = 0.05$ and $p = 5\times10^{-8}$, respectively. *P*-values have been capped at a y-axis value of 10 for the sake of readability. SCZ shows disproportionately strong signal and seems to be the main driver of the significance for several lead SNPs.

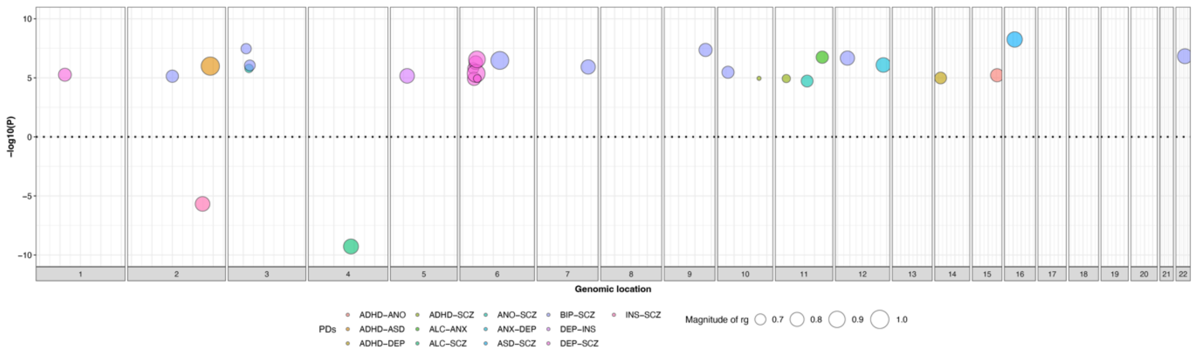

**Supplementary Fig. 17. Genomic position of all 29 significant local genetic correlations using LAVA**

All significant local genetic correlations after correcting for the total number of tests run across all PDs. The -log10 *p*-value multiplied by the sign of the correlation is displayed on the y-axis and dots are ordered along the x-axis according to genomic position. Each dot represents a significant local genetic correlation, with color indicating the combination of PDs. Size of the dots corresponds to the magnitude of the local genetic correlation.

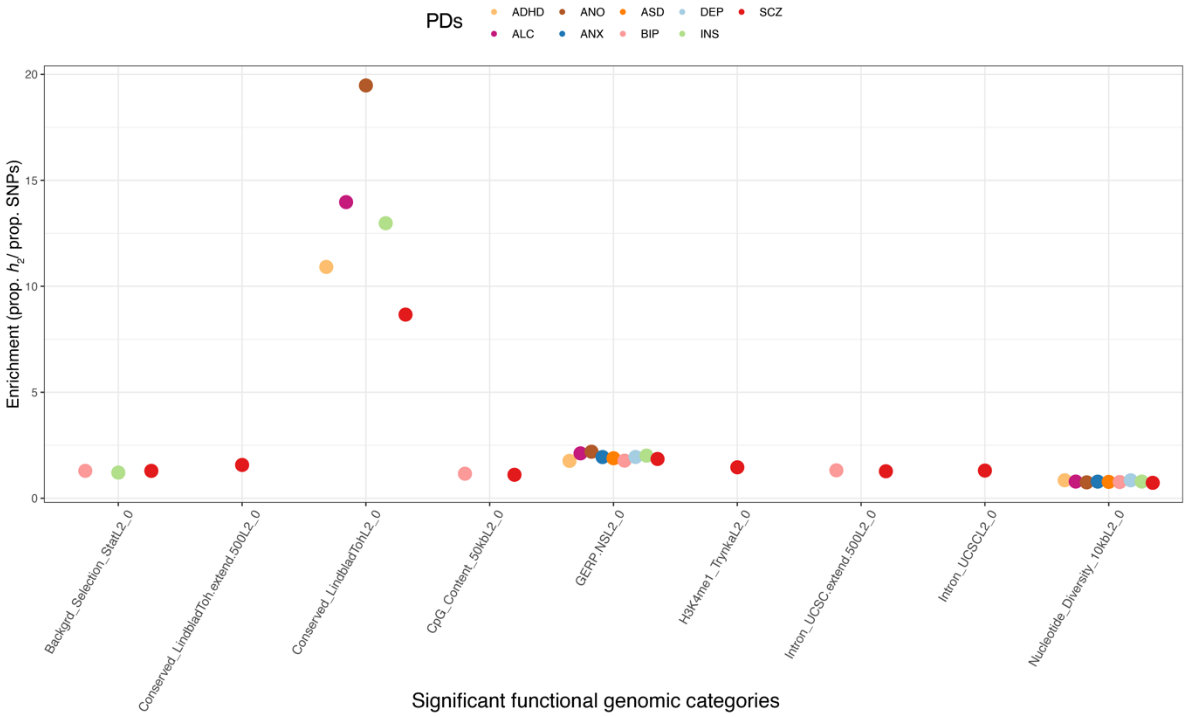

**Supplementary Fig. 18. Significantly enriched functional genetic categories across PDs**

Significant enrichment in multiple PDs after correcting for the number of functional categories is seen in specific (GERP_NSL) and general (Conserved Lindblad) conserved genetic regions as well as for Nucleotide diversity. Background selection showed enrichment for three PDs (BIP, INS, and SCZ). Enrichment on the y-axis is defined as the proportion of SNP heritability relative to the total number of SNPs in the functional category. Only functional genetic categories significant for at least one PD ($p < .05/61 = 8.20 \times 10^{-4}$) are displayed on the x-axis.

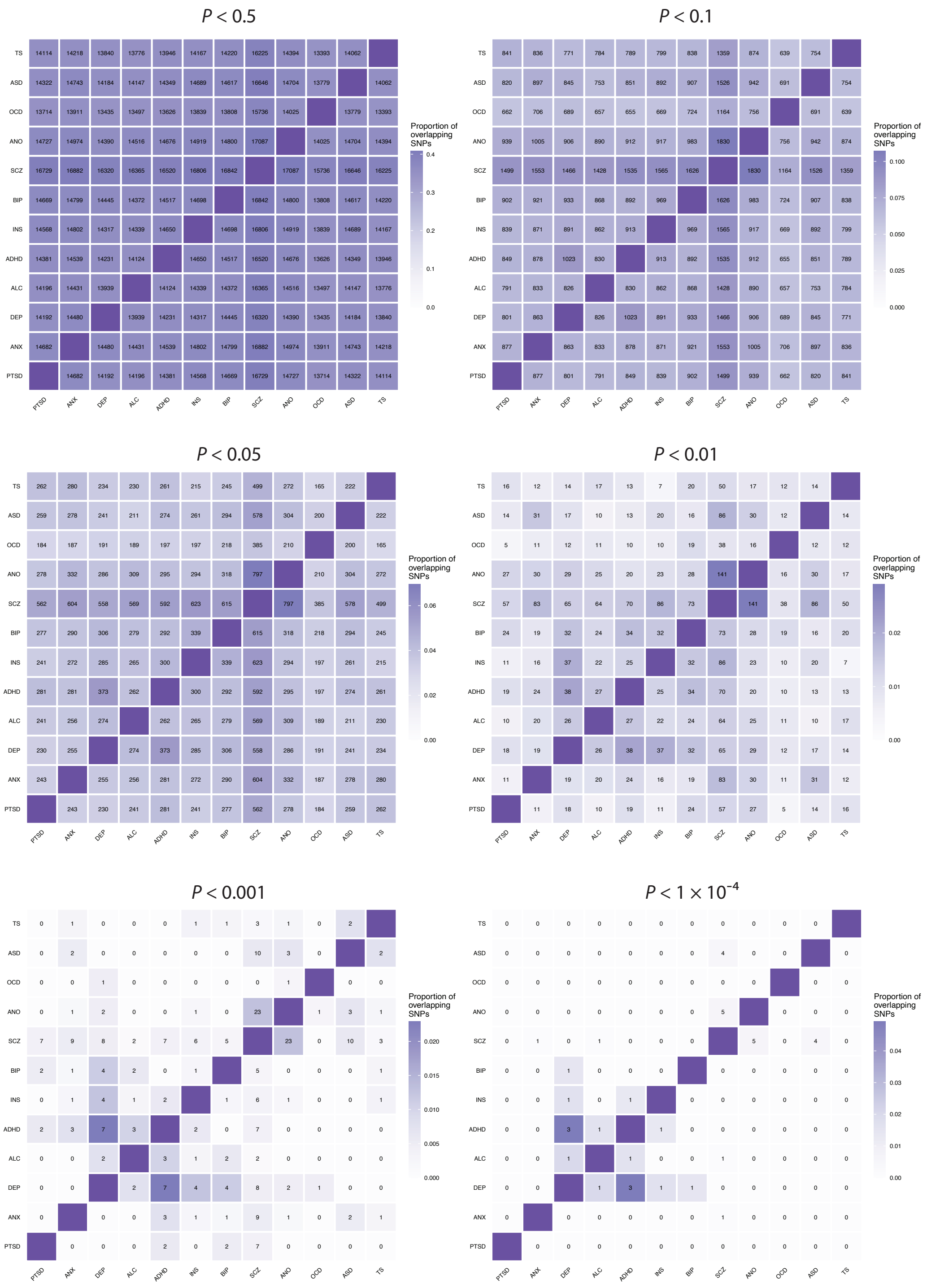

$P < 0.5$
$P < 0.1$
$P < 0.05$
$P < 0.01$
$P < 0.001$
$P < 1 \times 10^{-4}$
PTSD
ANX
DEP
ALC
ADHD
INS
BIP
SCZ
ANO
OCD
ASD
TS
Proportion of overlapping SNPs

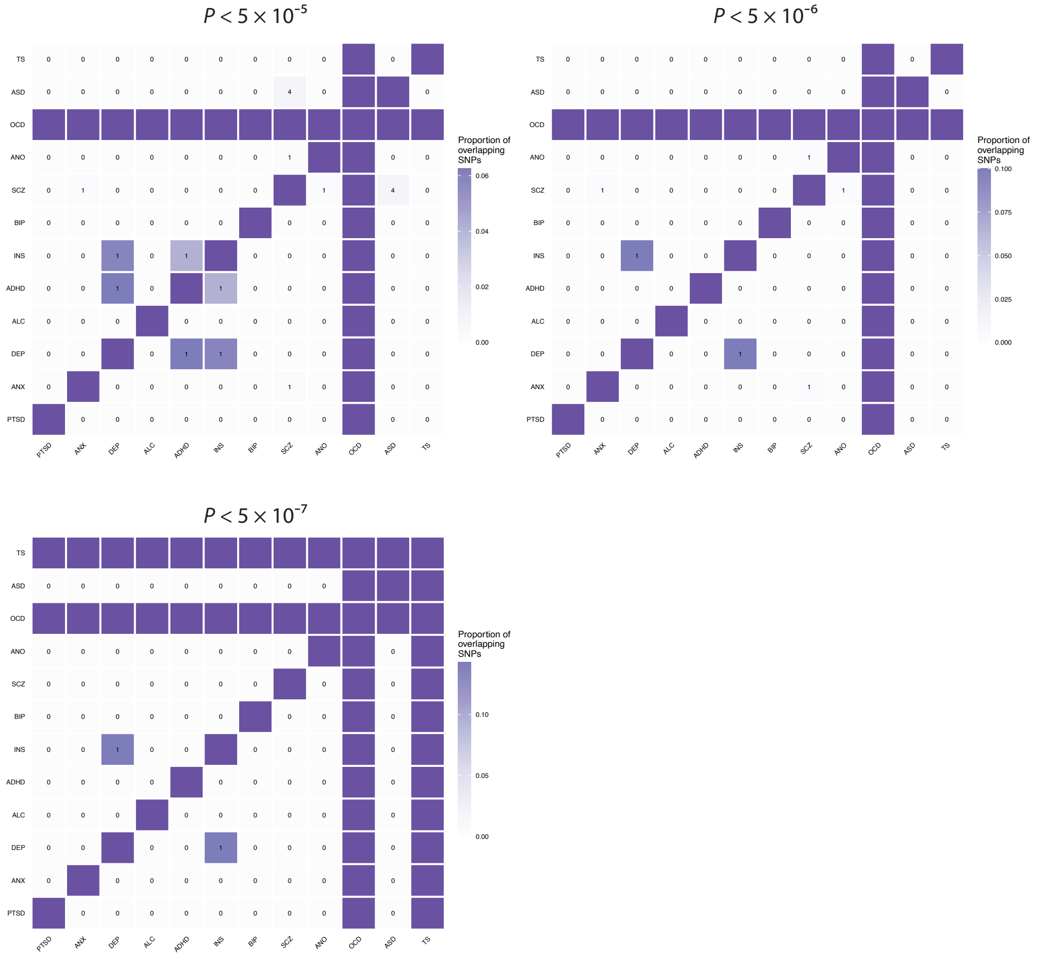

**Supplementary Fig. 19. Heatmaps of Fisher's exact test results for overlap in top SNPs (for different *p*-values thresholds) between the 12 PDs**

Digits in the cells indicate the number of shared SNPs for that specific pair of PDs that exceeded the *p*-values threshold. No value in off-diagonal cell indicates the absence of shared SNPs with a *p*-value lower than the given threshold for that PD and a '0' value indicates that although there were SNPs with *p*-values lower than the threshold for both PDs, these did not overlap. All heatmaps are symmetric in these figures.

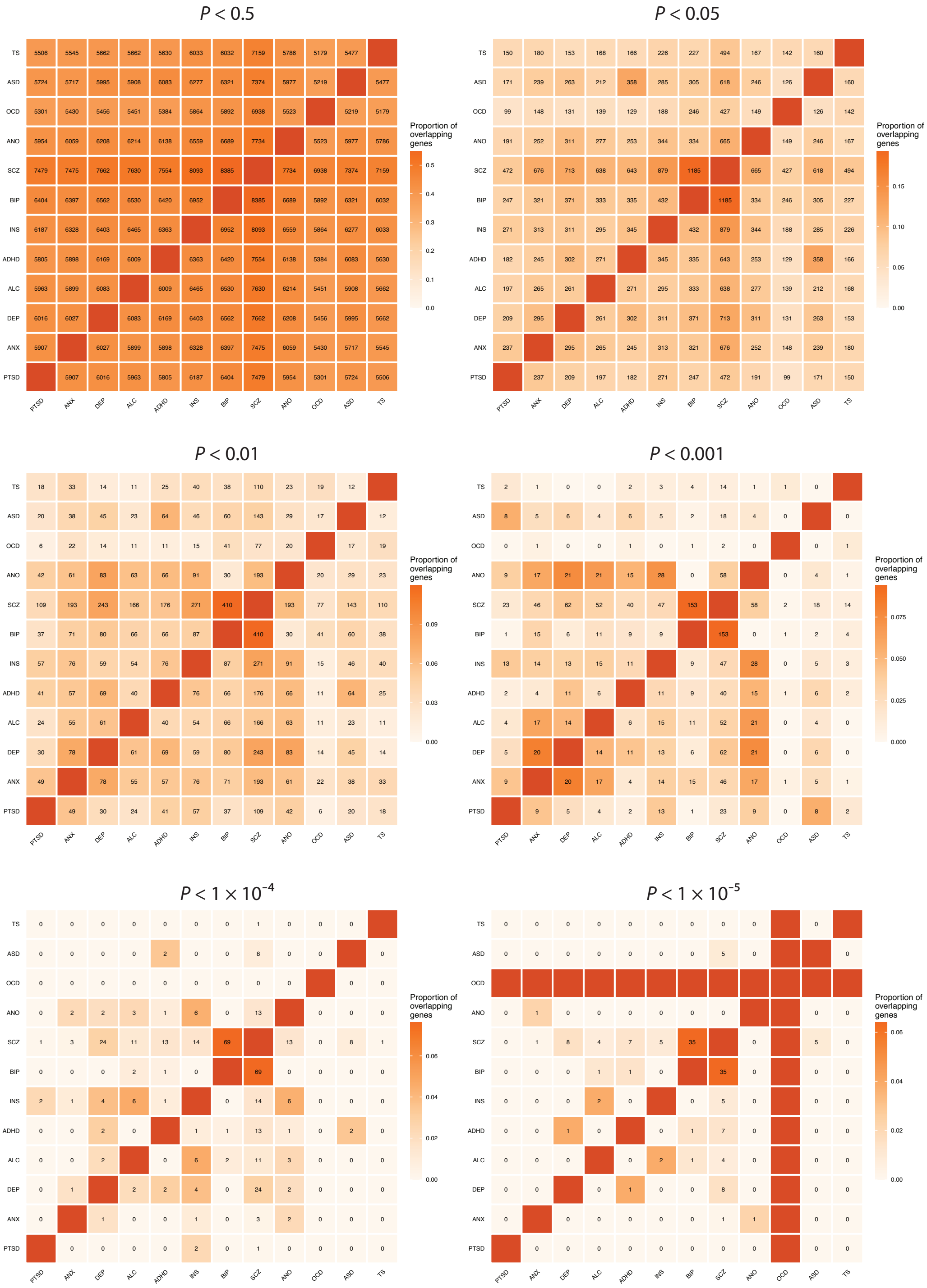

P < 0.5
P < 0.05
P < 0.01
P < 0.001
P < 1 × 10-4
P < 1 × 10-5
TS
ASD
OCD
ANO
SCZ
BIP
INS
ADHD
ALC
DEP
ANX
PTSD
Proportion of overlapping genes

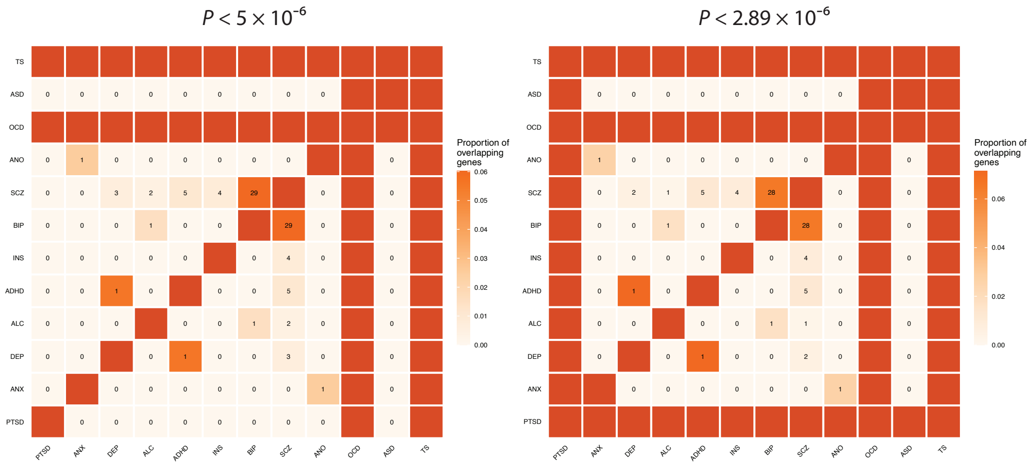

**Supplementary Fig. 20. Heatmaps of Fisher's exact test results for overlap in top gene (for different *p*-values thresholds) between the 12 PDs**

Digits in the cells indicate the number of shared genes for that specific pair of PDs that exceeded the *p*-values threshold. No value in off-diagonal cell indicates the absence of shared genes with a *p*-value lower than the given threshold for that PD and a '0' value indicates that although there were genes with *p*-values lower than the threshold for both PDs, these did not overlap. All heatmaps are symmetric in these figures.
