## Supplementary notes for "Exploring the genetic overlap between 12 psychiatric disorders"

Romero, C. et al.

### Supplementary Note 1. Additional information on summary statistics

*Attention-deficit/hyperactivity disorder (ADHD)*

The ADHD GWAS summary statistics by Demontis et al. (2019) are based on analysis of 12 cohorts: The iPSYCH-ADHD cohort and 11 cohorts aggregated by the PGC. The iPSYCH-ADHD cohort recruited cases through the Danish Psychiatric Central Research Registrar, and diagnostic assessment by psychiatrists were done following ICD-10. The PGC cohorts (seven case-control and four parent-offspring trio samples) consisted of European, Chinese, and North American ancestry. In these cohorts, clinicians defined cases according to research-based assessment criteria and recruitment was done through clinics and medical registries. The combined European sample size consisted of 19,099 cases and 34,194 controls (**Supplementary Table 1**).

*Alcohol use disorder (ALC)*

The ALC GWAS summary statistics by Kranzler et al. (2019) are based on data from the Million Veteran Program (MVP). MVP is a large-scale observational study and biobank, and phenotypes were collected though questionnaires as well as through electronic health records (EHR). GWAS was conducted in European Americans (EA), African Americans (AA), Latino Americans (LA), East Asian Americans (EAA) and South Asian Americans (SAA), stratified by ethnicity followed by a trans-population meta-analysis (N = 274,391). Here we use the case-control GWAS summary statistics based on analysis of EA individuals only (44,864 cases and 195,826 controls).

*Anorexia Nervosa (ANO)*

The ANO GWAS summary statistics by Watson et al. (2019) are based on four cohorts of European ancestry. All cohorts were case-control studies, and cases met criteria for ANO as defined in the DSM-III-r or DSM-IV, or ICD-8, ICD-9 or ICD-10 (apart from the UK Biobank cohort, in which case/control status was defined through self-reported diagnosis history). The combined sample size included 16,992 cases and 55,525 controls. Data from these participants were included in a case-control meta-analysis. The summary statistics used in the current study are based on this meta-analysis.

*Anxiety disorder (ANX)*

Here, we used ANX GWAS summary statistics from two recent studies. First, Purves et al. (2020) conducted a GWAS of lifetime anxiety disorder (25,453 cases and 58,113 controls) in UK Biobank participants of Western European ancestry. Cases either self-reported a lifetime professional diagnosis of one of the core five anxiety disorders (generalized anxiety disorder, social phobia, panic disorder, agoraphobia or specific phobia), or met criteria for a likely lifetime diagnosis of DSM-IV generalized anxiety disorder based on anxiety questions from the Composite International Diagnostic Interview (CIDI) Short-form questionnaire.

Second, Levey et al. analyzed 192,256 European American men and women (28,525 cases and 163,731 controls) who participated in the Million Veteran Program (MVP). We used the secondary phenotype analyzed by Levey et al. ("self-reported physician diagnosis of anxiety disorder"), since that matched the phenotype definition used by Purves et al. best. Individuals responding to the following question with 'yes' were assigned case status: "Please tell us if you have been diagnosed with the following conditions: anxiety reaction/panic disorder."

Summary statistics from Purves et al. and Levey et al. were case-control meta-analyzed using mvGWAMA (see **Methods**), resulting in a pooled N = 275,822 (53,978 cases and 221,844 controls).

*Autism spectrum disorder (ASD)*

The GWAS summary statistics on ASD by Grove et al. (2018) are based on six cohorts of European ancestry. Five of the cohorts were family-based, and cases were defined according to research-based assessment criteria and clinical consensus diagnosis. The case-control iPSYCH-ADHD cohort recruited cases through the Danish Psychiatric Central Research Registrar, and a psychiatrist assessed diagnosis following ICD-10 criteria. The combined sample size consisted of 18,381 cases and 27,969 controls. The summary statistics used in the current study are based on this case-control meta-analysis.

*Bipolar disorder (BIP)*

The BIP GWAS summary statistics by Stahl et al. (2019) are based on analysis of 32 cohorts from Europe, North America, and Australia. All participants were of European ancestry. A clinical diagnosis of lifetime BIP was established for all cases using structured assessment or checklists by a clinician, or through a review of medical history. All diagnoses followed DSM-IV, ICD-9 or ICD-10 criteria. The combined sample size consisted of 20,352 cases and 31,358 controls. Data from these participants were included in a case-control meta-analysis. The summary statistics used in the current study are based on this meta-analysis.

*Insomnia disorder (INS)*

The INS GWAS summary statistics by Jansen et al. (2019) are based on analysis of UK Biobank and 23andMe cohorts. All participants were of European ancestry. Participants that reported that they, during the last four weeks, usually had trouble falling asleep at night or usually woke up in the middle of the night were defined as cases (i.e. suffering from INS), while those responding never/rarely or sometimes were assigned control status. The total sample size consisted of 1,331,010 participants. Data from these participants were included in a case-control meta-analysis. Due to data use restrictions, the summary statistics used in the current study are based on a subset of the individuals analyzed by Jansen et al., i.e., the UKB cohort only; N = 386,533 (109,402 cases and 277,131 controls).

*Depression (DEP)*

The depression GWAS summary statistics by Wray et al. (2018) result from a case-control meta-analysis of seven cohorts with individuals of European ancestry. In six cohorts, case status was determined by a range of methods (self-reported medical history or symptoms, interviews, and national treatment registers), while in the remaining cohort a review of medical history or structured assessment by a clinician following DSM-V, ICD-9, or ICD-10 criteria was applied. The total sample size of all cohorts was 135,458 cases and 344,901 controls. Data from these participants were included in a case-control meta-analysis. Due to data use restrictions, the summary statistics used in the current study are based on the results excluding the 23andMe cohort (i.e., 59,851 cases and 113,154 controls).

*Obsessive-compulsive disorder (OCD)*

The OCD GWAS summary statistics by Arnold et al. (2018) are based on the IOCDF-GC and OCGAS cohorts. All participants were of European ancestry. A clinical diagnosis of OCD was established for all cases following DSM-IV criteria. The combined sample size consisted of 2,688 cases and 7,037 controls. The summary statistics resulting from the case-control meta-analysis of these two cohorts were used in the current study.

*Post-traumatic stress disorder (PTSD)*

The PTSD GWAS summary statistics by Stein et al. (2021) are based on analysis of subjects enrolled in the Million Veteran Program (MVP). Using information from electronic health records, an algorithm-derived approach assigned participants to either case or control status. The summary statistics of the case-control GWAS analyzing 214,408 individuals of European American ancestry (36,301 cases and 178,107 controls) were used in the current study.

*Schizophrenia (SCZ)*

The SCZ GWAS summary statistics presented by The Schizophrenia Working Group of the Psychiatric Genomics Consortium (preprint) are based on analysis of 91 cohorts. The GWAS was conducted in European and East Asian ancestry. Cases met clinical DSM-IV or ICD-10 criteria for either schizophrenia or schizoaffective disorder. In the current study, summary statistics based only on subjects from European descent were included; N = 161,405 (67,390 cases and 94,015 controls)

*Tourette syndrome (TS)*

The TS GWAS summary statistics by Yu et al. (2019) are based on analysis of four cohorts of individuals from European descent. Three of the cohorts consisted of case-control studies, while the latter was a family-based cohort. Cases met criteria for TS as defined by DSM-IV-TR or DSM-V, and were recruited by specialty clinics or through correspondence and clinical assessment online. The combined sample size consisted of 4,819 cases and 9,488 controls. The summary statistics obtained in the case-control meta-analysis of these four cohorts are used in the current study.

### Supplementary Note 2. Comparison of summary statistics before and after filtering to shared SNPs

#### Rationale for filtering

Prior to any analyses, we filtered the summary statistics of all 12 PDs, retaining only those SNPs that were represented in all 12 summary statistics. We emphasize here that we worked with summary statstics from previous GWASs, and while imputation of SNP genotypes is common before the SNP-trait association is being established in GWAS, imputation of SNP genotypes as well as their accompanying trait-association statistics (e.g., beta, Z-score, *p*-value) after GWAS has taken place is neither common nor straightforward. Even though this filtering step implied an often substantial decrease in the number of SNPs (**Supplementary Table 1**), there are several reasons why we believe that the discussed analyses should be based on the same SNPs, such that any observed difference in the observed genetic signal, or lack of genetic overlap between PDs, cannot be attributed to differences in the SNPs representing each individual PD:

First, regarding the cross-trait meta-analysis: inputting only the SNPs that are represented in all PDs assures that the cross-trait meta-analytic signal can actually be representative of all traits, i.e., one avoids situations in which part of the meta-analytic signal is driven by SNPs that were only measured in 1 or a few traits. After all, the genetic similarity between PDs cannot be established for those SNPs that have not been measured in all included PDs.

Second, in comparing the genetic results of individual PDs (i.e., number of associated SNPs, genes, and gene-sets), we wanted to exclude the possibility that results were not comparable because PDs were represented by different (numbers of) SNPs. Specifically, we don't want PDs to seem incomparable with regard to associated SNPs, genes, or gene-sets because of differences in the representations of the genome between individual PDs. Regarding associated SNPs, a conclusion like "traits A and B share 50% of GWS SNPs" is only interpretable if the same SNPs were actually represented in both summary statistics. Similarly, as gene-based analyses evaluate the joint signal of all SNPs residing within a gene, the possibility exists that differences between PDs in associated genes is prompted by differential representation of SNPs between PDs: fair comparison of associated genes between PDs can only be achieved if the gene-based analyses are based on the same set of SNPs. A similar argument holds for gene-set analyses and the establishment of the significance of genomic regions.

Third, various analyses, like local genetic correlation analysis in LAVA and the Fisher exact test, are by default restricted to only those SNPs that are shared between traits. It would be impossible to relate the results of these analyses to those obtained using SNP/region/gene/gene-set analyses if the latter were based on different SNP sets.

Overall, we believe that the genetic signal of multiple PDs, as well as their overlap, can only be evaluated fairly if these are based on the same set of SNPs, and similarly, that the results of different analyses can only be compared if these are based on the same SNPs.

**Impact of filtering**

In order to examine how the filtering may have affected the results, we conducted a number of analyses. First of all, we estimated SNP-based heritability ($h^2_{SNP}$) for both the filtered and the unfiltered versions of the summary statistics, and no significant differences were observed (**Supplementary Fig. 3**).

Subsequently, all within-PD genetic correlations between the unfiltered and filtered versions of the summary statistics were all the same (see **Fig. SN1** below). In addition, patterns of global cross-PD genetic correlations were highly congruent (Pearson's correlation $r$ = 0.99) when using unfiltered vs. filtered summary statistics (**Supplementary Figure 4**).

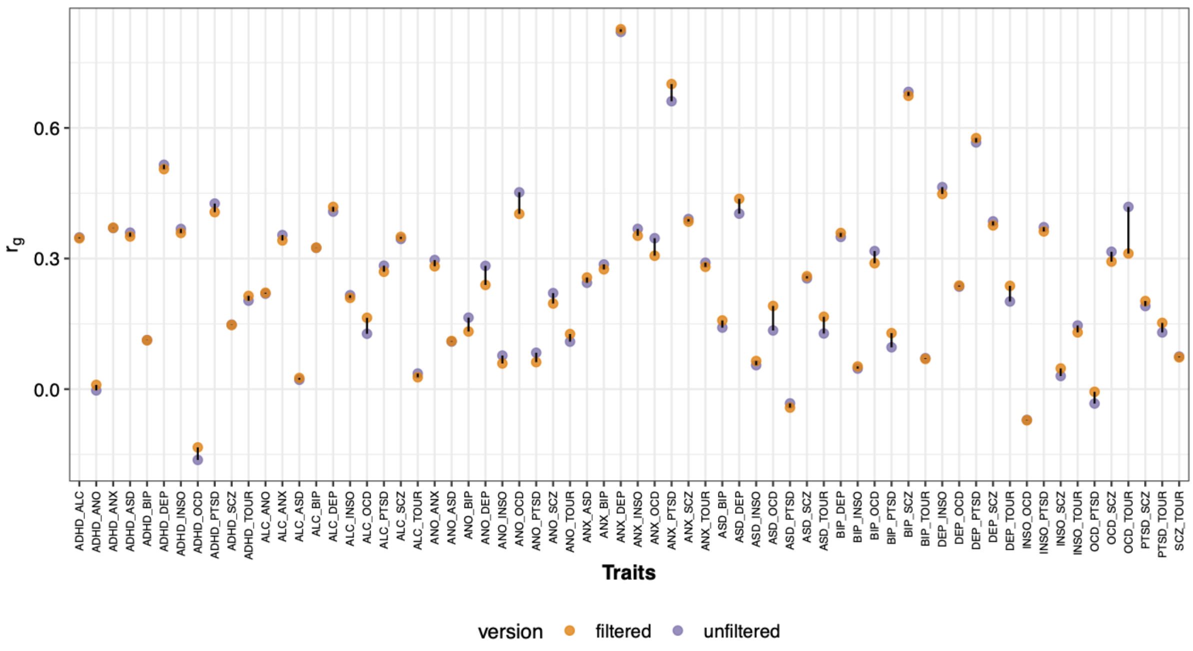

***Fig. SN1. Difference in genetic correlations across PD pairs between filtered and unfiltered summary statistics***

*Genetic correlations based on filtered (yellow) summary statistics show no indication of change in strength between PDs compared to genetic correlations based on unfiltered (purple) summary statistics. Strongest difference is observed between OCD-TS which after filtered is no longer significant after correcting for multiple testing.*

When comparing significant annotations between the filtered and unfiltered versions of the summary statistics, we observed a substantial drop in the number of lead SNPs, risk loci, FUMA mapped genes, and MAGMA genes (see **Table SN1** on page 9) for most of the PDs as a result of filtering while only a few PDs differed in terms of gene-sets, tissues, and cell-types (see **Table SN2** on page 10). This indicates that filtering leads to a reduction in statistical power to pick up significant genetic annotations than unfiltered summary statistics.

Crucially, however, the difference in observed *overlap* of genetic annotations between PDs did not change much as a result of filtering (see **Table SN3** on page 11). For FUMA and MAGMA genes there was only an increase in genes overlapping with more than one PD when not filtering, though not for genes overlapping with more than two PDs. No difference was observed for gene sets both when correcting only within the number of gene-sets and for all 10,126 items tested, while cell-types and tissues both showed an increase in overlap for unfiltered summary statistics. After correcting for all items and number of PDs, 3 cell types and 5 tissues remained significant between SCZ and BIP. Substantial overlap between these PDs was also observed consistently in the result of other analyses using the filtered summary statistics and is thus not contradicting the main conclusions of this study.

**Table SN1. Comparison of no. of associated lead SNPs, genomic risk loci and genes between filtered and unfiltered summary statistics for each of the 12 PDs.**

| | No. of lead SNPs | | | | No. of genomic loci | | | | No. of MAGMA genes | | | | No. of FUMA mapped genes | | | |
|---|---|---|---|---|---|---|---|---|---|---|---|---|---|---|---|---|
| PD | Fil | Unfil | Diff | % decrease | Fil | Unfil | Diff | % decrease | Fil | Unfil | Diff | % decrease | Fil | Unfil | Diff | % decrease |
| ADHD | 8 | 13 | 5 | 38% | 7 | 12 | 5 | 42% | 9 | 15 | 6 | 40% | 29 | 49 | 20 | 41% |
| ALC | 10 | 12 | 2 | 17% | 8 | 8 | 0 | 0% | 17 | 14 | -3 | -21% | 61 | 64 | 3 | 5% |
| ANO | 4 | 9 | 5 | 56% | 4 | 8 | 4 | 50% | 31 | 34 | 3 | 9% | 72 | 99 | 27 | 27% |
| ANX | 2 | 4 | 2 | 50% | 2 | 4 | 2 | 50% | 8 | 8 | 0 | 0% | 4 | 11 | 7 | 64% |
| ASD | 1 | 2 | 1 | 50% | 1 | 2 | 1 | 50% | 3 | 3 | 0 | 0% | 10 | 8 | -2 | -25% |
| BIP | 12 | 15 | 3 | 20% | 12 | 15 | 3 | 20% | 43 | 48 | 5 | 10% | 97 | 116 | 19 | 16% |
| DEP | 4 | 6 | 2 | 33% | 4 | 5 | 1 | 20% | 6 | 21 | 15 | 71% | 6 | 103 | 97 | 94% |
| INS | 12 | 15 | 3 | 20% | 12 | 14 | 2 | 14% | 15 | 21 | 6 | 29% | 126 | 128 | 2 | 2% |
| OCD | 0 | 0 | 0 | NA | 0 | 0 | 0 | NA | 0 | 0 | 0 | 0% | 0 | 0 | 0 | 0% |
| PTSD | 2 | 3 | 1 | 33% | 2 | 3 | 1 | 33% | 0 | 1 | 1 | 100% | 2 | 2 | 0 | 0%% |
| SCZ | 236 | 291 | 55 | 19% | 195 | 237 | 42 | 18% | 416 | 633 | 217 | 34% | 1582 | 1840 | 258 | 14% |
| TS | 0 | 1 | 1 | 100% | 0 | 1 | 1 | 100% | 0 | 1 | 1 | 100% | 0 | 0 | 0 | 0%% |

*Note: For the current comparison we considered genes to be significant in MAGMA's gene-based analysis when $p < 0.05$ / no. of genes tested ($p < 0.05$ / 17,287). PD = psychiatric disorder; Fil = filtered summary statistics; Unfil = unfiltered summary statistics; Diff = difference between unfiltered and filtered; % decrease = percentage decrease when using filtered instead of unfiltered summary statistics.*

**Table SN2. Comparison of no. of associated gene sets, tissue types and cell types between filtered and unfiltered summary statistics for each of the 12 PDs.**

| | No. of gene sets | | | | No. of tissues | | | | No. of cell types | | | |
|---|---|---|---|---|---|---|---|---|---|---|---|---|
| **PD** | **Fil** | **Unfil** | **Diff** | **% decrease** | **Fil** | **Unfil** | **Diff** | **% decrease** | **Fil** | **Unfil** | **Diff** | **% decrease** |
| ADHD | 0 | 0 | 0 | NA | 0 | 0 | 0 | NA | 0 | 0 | 0 | NA |
| ALC | 3 | 0 | -3 | -100% | 0 | 0 | 0 | NA | 0 | 0 | 0 | NA |
| ANO | 0 | 0 | 0 | NA | 0 | 0 | 0 | NA | 0 | 0 | 0 | NA |
| ANX | 0 | 0 | 0 | NA | 0 | 0 | 0 | NA | 0 | 0 | 0 | NA |
| ASD | 0 | 0 | 0 | NA | 0 | 0 | 0 | NA | 0 | 0 | 0 | NA |
| BIP | 0 | 0 | 0 | NA | 0 | 0 | 0 | NA | 0 | 11 | 11 | NA |
| DEP | 0 | 0 | 0 | NA | 0 | 0 | 0 | NA | 0 | 0 | 0 | NA |
| INS | 0 | 2 | 2 | 100% | 0 | 1 | 1 | 100% | 1 | 1 | 0 | 0% |
| OCD | 0 | 0 | 0 | NA | 0 | 0 | 0 | NA | 0 | 0 | 0 | NA |
| PTSD | 0 | 0 | 0 | NA | 0 | 0 | 0 | NA | 0 | 0 | 0 | NA |
| SCZ | 1 | 4 | 3 | 75% | 13 | 13 | 0 | 0% | 114 | 109 | -5 | -5% |
| TS | 0 | 1 | 1 | 100% | 0 | 0 | 0 | NA | 0 | 0 | 0 | NA |

*Note: For the current comparison we considered gene sets, tissue types and cell types significant when p < 0.05 / total no. of gene sets + no. tissues types + no. of cell types tested (p < 0.05 / (9508 + 53 + 565)). PD = psychiatric disorder; Fil = filtered summary statistics; Unfil = unfiltered summary statistics; Diff = difference between unfiltered and filtered; % decrease = percentage decrease when using filtered instead of unfiltered summary statistics.*

**Table SN3. Comparison of no. of associated genes, tissues, cell types that overlap between filtered and unfiltered summary statistics for each of the 12 PDs**

| | | | | | Gene sets | | | | Cell types | | | | Tissues | | | |
|---|---|---|---|---|---|---|---|---|---|---|---|---|---|---|---|---|
| **Overlap** | **Mapped genes** | | **MAGMA genes** | | **Within Bonf** | | **Between Bonf** | | **Within Bonf** | | **Between Bonf** | | **Within Bonf** | | **Between Bonf** | |
| **More than** | **Fil** | **Unfil** | **Fil** | **Unfil** | **Fil** | **Unfil** | **Fil** | **Unfil** | **Fil** | **Unfil** | **Fil** | **Unfil** | **Fil** | **Unfil** | **Fil** | **Unfil** |
| 6 PDs | 0 | 0 | 0 | 0 | 0 | 0 | 0 | 0 | 0 | 0 | 0 | 0 | 0 | 1 | 0 | 0 |
| 5 PDs | 0 | 0 | 0 | 0 | 0 | 0 | 0 | 0 | 0 | 0 | 0 | 0 | 2 | 2 | 0 | 0 |
| 4 PDs | 0 | 0 | 0 | 0 | 0 | 0 | 0 | 0 | 0 | 0 | 0 | 0 | 2 | 2 | 0 | 0 |
| 3 PDs | 0 | 0 | 0 | 0 | 0 | 0 | 0 | 0 | 0 | 0 | 0 | 0 | 3 | 5 | 0 | 0 |
| 2 PDs | 3 | 2 | 2 | 0 | 0 | 0 | 0 | 0 | 0 | 1 | 0 | 0 | 5 | 7 | 0 | 1 |
| 1 PD | 221 | 378 | 39 | 57 | 0 | 0 | 0 | 0 | 6 | 33 | 0 | 11 | 10 | 8 | 0 | 8 |

*Note: For the current comparison we considered genes to be significant in MAGMA's gene-based analysis when p < 0.05 / no. of genes tested (p < 0.05 / 17,287 For pathway and property enrichment analyses, we compared the filtered and unfiltered summary statisitics based on two different p-value thresholds: 1) multiple testing correction was applied within each category separately (column: within Bonf), resulting in a p < 0.05/ 9508, p < 0.05/53, and p < 0.05/565 threshold for gene-sets, tissues and cell types, respectively, and 2) multiple testing correction was applied on the combined number of gene-sets, tissue types and cell types (column between Bonf), resulting in a p < 0.05 / (9508 + 53 + 565). Numbers in tables are cumulative. PD = psychiatric disorder; Fil = filtered summary statstics; Unfil = unfiltered summary statistics.*

### Supplementary Note 3. Regional examples with positive and negative correlations among PDs.

We used Local Analysis of [co]Variant Association (LAVA[1]) to estimate bivariate local genetic correlations in those regions that showed univariate signal for both PDs at $p$ < 1e-4. This resulted in running a total of 2,532 bivariate tests. Of the 29 regions that showed significant local genetic correlation after Bonferroni correction ($p_{BON}$ = .05/(number of bivariate test conducted) = 0.05 / 2,532 = $1.97\times10^{-5}$), we here discuss five in more detail, as only these regions include genes that are mapped to both PDs in the PD pair.

Locus 692, chromosome 4 (bp 102,544,804-104,384,534) contains 1,683 SNPs (139 PCs) showing a strongly negative correlation between ALC and SCZ (rho = –0.84, $p = 5.22 \times 10^{-10}$, R2 = 0.70; **Fig. SN2**). This region contains 11 genes, out of which 3 genes (*NFKB1*, *SLC9B1*, and *BDH2*) were mapped to both traits by chromatin interaction mapping and 1 gene (*SLC39A8*) was mapped to both PDs by positional mapping. *NFKB1*, nuclear factor-kappaB1, is a central transcription factor found in all cell types. It is fast acting in response to cytokines and stress, and is essential in regulating immunological response to infections[2]. *NFKB1* has been previously linked to common variable immunodeficiency and rheumatoid arthritis.

*SLC9B1*, solute carrier family 9 member b1, is a sodium/hydrogen exchanger mainly expressed in the testis and involved in sperm motility and fertility. *SLC9B1* has been associated with Wolfram syndrome an autosomal recessive disorder, which, in 60 percent of cases, result in the development of a neurological or psychiatric disorder with psychiatric symptoms including psychosis, severe depression, and impulsive and aggressive behavior[3].

*BDH2*, 3-hydroxybutyrate dehydrogenase 2, is a dehydrogenase that plays a key role in iron assimilation and homeostasis. It is related to differential utilization of ketone bodies by neuron cell lines[2].

*SLC39A8*, solute carrier family 39 member 8, is found in the plasma membrane and mitochondria and is related to cellular import of zinc at the start of inflammation. Rare variants in *SLC39A8* may result in Leigh-like mitochondrial syndrome with symptoms of developmental delay, dystonia and seizures[2]. All aforementioned genes could confer opposite effects towards the genetic liability of ALC and SCZ, but this needs to be validated.

Locus 62, chromosome 1 (bp 72,513,120-72,992,170) contains 1,773 SNPs (68 PCs) showing a positive correlation between DEP and SCZ (rho = 0.76, $p = 5.47 \times 10^{-6}$, R2 = 0.58; **Fig. SN3**). This region contains only one gene, *NEGR1,* which was positionally mapped to DEP and by chromatin interaction to SCZ. *NEGR1* is a neuronal growth regulator that participate in cell adhesion and may function as a trans-neural growth-promoting factor[2]. *NEGR1* has been previously implicated in Niemann-Pick disease and leptin dysfunction and previously associated to body size and cognition phenotypes (BMI, educational attainment, body height, cognitive function measurement). *NEGR1* could explain part of DEP SCZ comorbidity in this locus, though this needs to be validated.

Locus 457, chromosome 3 (bp 36,840,137-38,729,767) contains 2,138 SNPs (212 PCs) showing a positive correlation between BIP and SCZ (rho = 0.68, $p = 3.4 \times 10^{-8}$, R2 = 0.47; **Fig. SN4**). This region contains 25 genes, out of which only 1 (*TRANK1*) was mapped to both PDs. *TRANK1*, Tetratricopeptide Repeat and Ankyrin Repeat Containing 1, is expressed in numerous tissues and is predicted to enable ATP binding through hydrolase activity[2]. *TRANK1* gene alterations are associated to Kleine-Levin Hibernation syndrome which is a rare disorder characterized by hypersomnia and behavioral and cognitive changes[3]. *TRANK1* could explain part of the BIP SCZ comorbidity in this locus, though this needs to be validated.

Locus 466, chromosome 3 (bp 51,953,969-54,074,844) contains 2,067 SNPs (174 PCs) showing a positive correlation between BIP and SCZ (rho = 0.70, $p = 8.9 \times 10^{-7}$, R2 = 0.49; **Fig. SN5**). This region contains 59 genes out of which 24 genes have been mapped to both PDs. Out of these 24 genes, 10 genes (*ITIH1, ITIH3, ITIH4, GNL3, NEK4, PBRM1, TMEM110, MUSTN1, SMIM4,* and *GLT8D1*) were additionally GWGAS significant for both PDs. *ITIH1, ITIH2* and *ITIH4* code for members of the inter-alpha-trypsin inhibitor, which function as a protease inhibitor and are related to endopeptidase inhibitor activity and cell adhesion-cell matrix glycoconjugates[2].

*GNL3* and *NEK4* interact with different parts of the cell-cycle and are involved with *stem cell proliferation* and *cell cycle arrest in response to double stranded DNA damage*, respectively[2].

*PBRM1* is involved in chromatin organization by coding for a subunit of ATP dependent chromatin remodeling complexes involved in transcriptional activation upon activation of nuclear hormone receptors[2].

*TMEM110* function as a regulator of calcium entry at junctional sites that connect endoplasmic reticulum and the plasma membrane[2]. *MUSTN1* is thought to be involved in the development and regeneration of the musculoskeletal system[2]. The function of *SMIM4* and *GLT8D1* is not clear. All aforementioned genes could explain part of BIP SCZ comorbidity in this locus, though it is not clear which genes are more likely to be involved in relevant biological processes.

Locus 464, chromosome 3 (bp 47,588,462-50,387,742) contains 2,026 SNPs (120 PCs) showing a positive correlation between ANO and SCZ (rho = 0.65, $p = 1.6 \times 10^{-6}$, R2 = 0.42; **Fig. SN6**). This region contains 100 genes, out of which 16 genes (*KLHDC8B, RHOA, CAMKV, AMIGO3, GNAT1,*

*GNAI2, QARS, AMT, TRAIP, TUSC2, WDR6, TCTA, GMPPB, MON1A, CCDC71,* and *HYAL2*) were mapped to both PDs. Common functions between these genes are known for several.

*KLHDC8B, RHOA, CAMKV, AMIGO3, GNAT1,* and *GNAI2* are all genes related to signaling transduction, either by direct or indirect involvement in signaling cascades[2]; *KLHDC8B* forms beta-propeller proteins structures that allow protein-protein interactions; *RHOA* is part of a small family GTPases that perpetrates signaling pathways involved in action cytoskeleton reorganization; *GNAT1* and *GNAI2* are G protein subunit alpha members and are involved in GTPase activity and the phospholipase C pathway, respectively; and *AMIGO3* codes for an adhesion protein that can contribute to signal transduction through its intracellular domain.

*QARS* and *AMT* are both associated to encephalopathy symptoms, through processes involving linkage between amino acids-tRNA nucleotide triplets and glycine cleavage, respectively[2].

*TRAIP, TUSC2,* and *WDR6* acts as suppressors of cell growth and proliferation[2]. The remaining genes are associated to *Cellular fusion during osteoclastogenesis* (*TCTA*), *Production of N-linked oligosaccharides* (*GMPPB*), *Vesicle mediated transport* (*MON1A*), *Cellular lipid metabolism* and *regulator of fat cell differentiation* (*CCDC71,* predicted), and *Glycosaminoglycan metabolism* (*HYAL2*)[2]. All aforementioned genes could explain part of ANO SCZ comorbidity in this locus, though this needs to be validated.

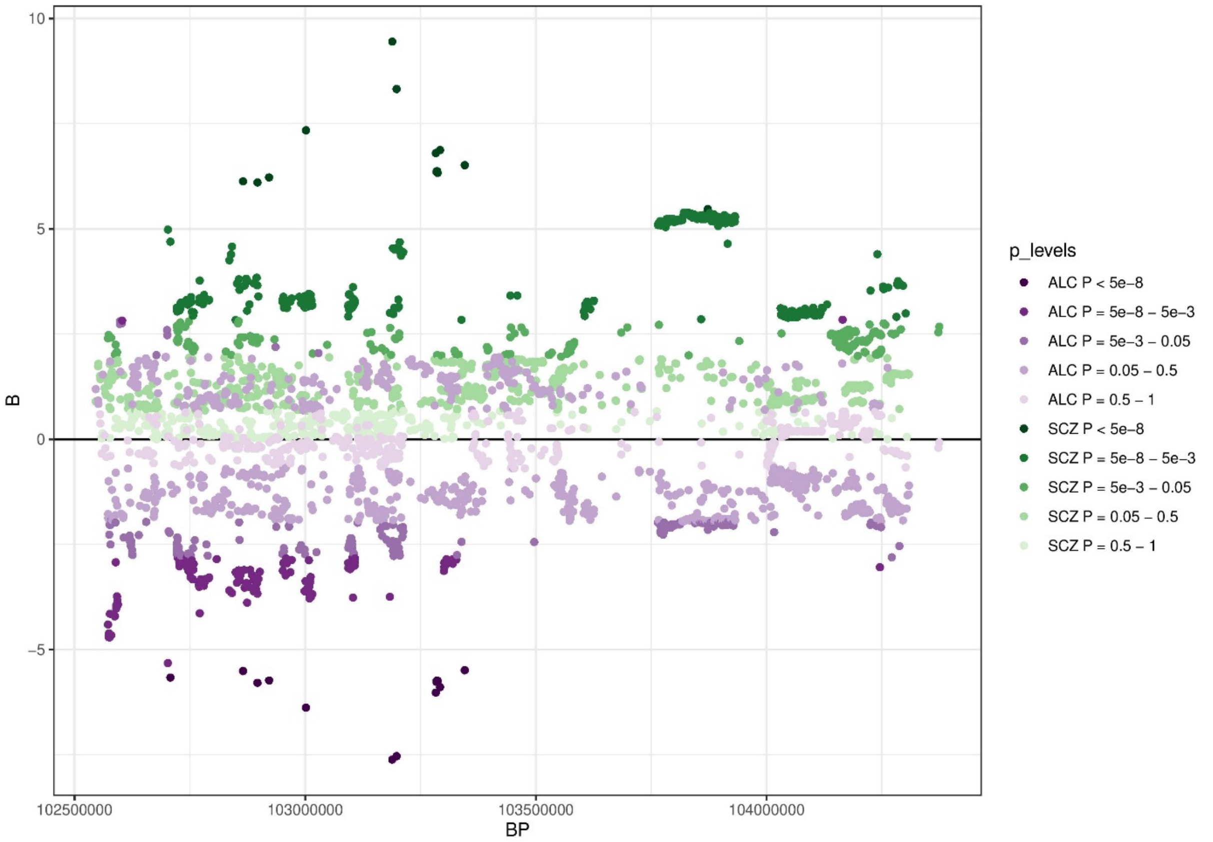

***Fig. SN2. Standardized betas of SNPs within region chr4:102,544,804-104,384,534) where a strong, negative correlation between ALC and SCZ was observed***

*Each dot represents a SNP. On the x-axis is the genomic position in base pairs. The y-axis shows the effect size of the SNP as standardized beta. Color saturation is proportional to significance level of the SNP (with darker colors representing lower p-values). SCZ SNP alleles (in green) were aligned to only show positive betas and used as a reference to align alleles of ALC (in purple).*

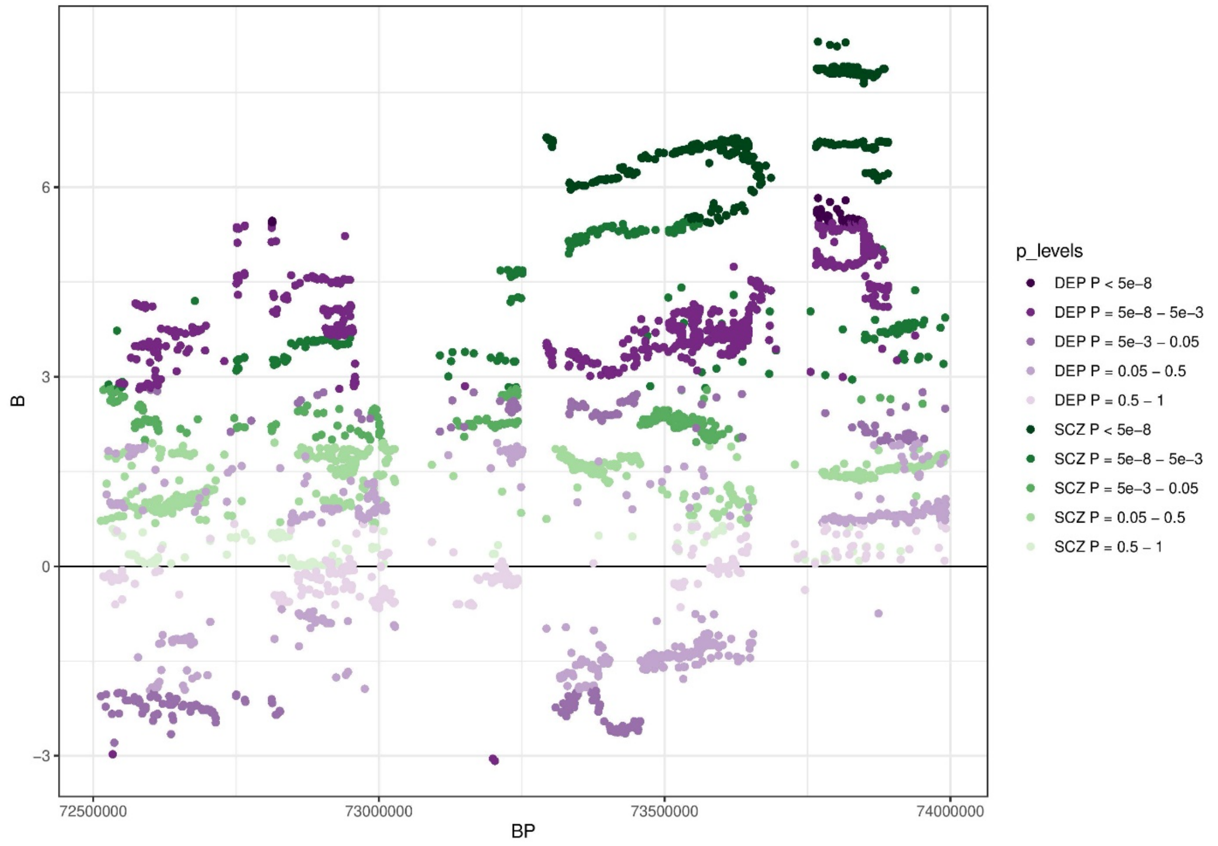

***Fig. SN3. Standardized betas of SNPs within region chr1:72,513,120-73,992,170 where a strong, positive correlation between DEP and SCZ was observed***

*Each dot represents a SNP. On the x-axis is the genomic position in base pairs. The y-axis shows the effect size of the SNP as standardized beta. Color saturation is proportional to significance level of the SNP (with darker colors representing lower p-values). SCZ SNP alleles (in green) were aligned to only show positive betas and used as a reference to align alleles of DEP (in purple).*

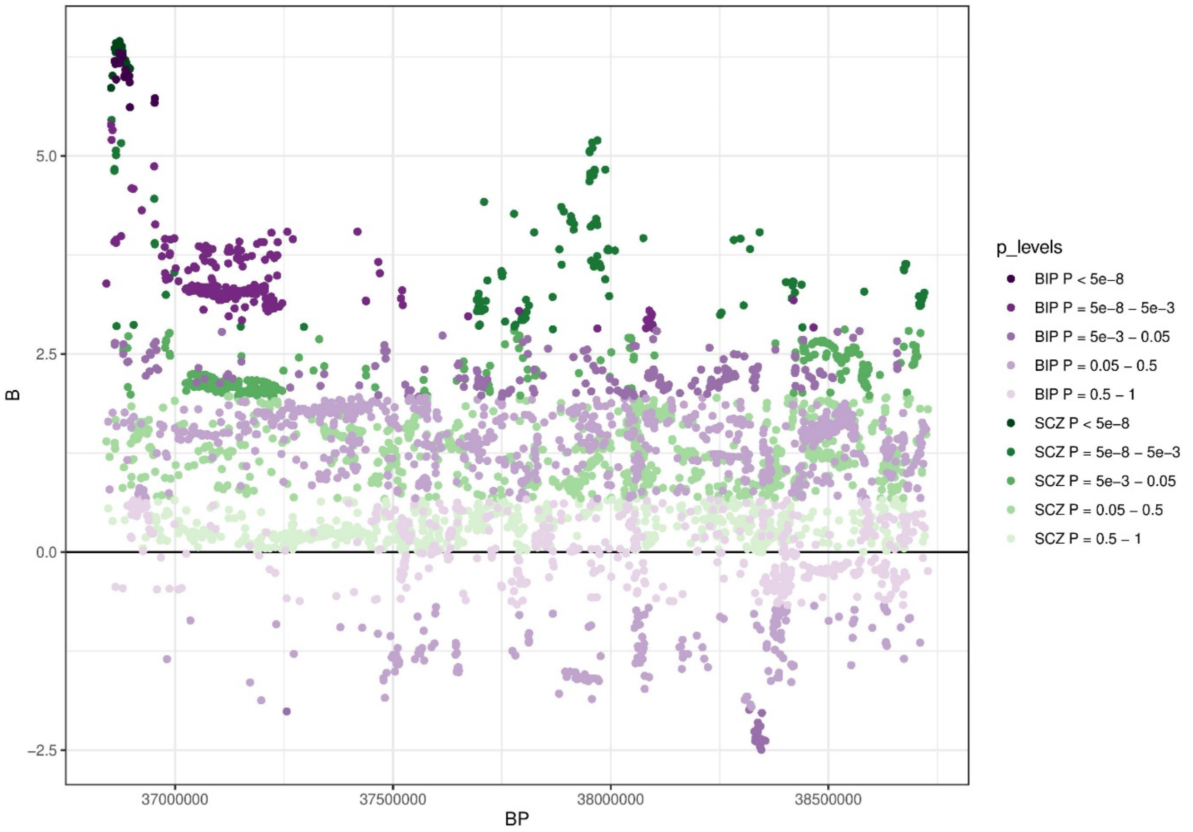

***Fig. SN4. Standardized betas of SNPs within region chr3:36,840,137-38,729,767 where a strong, positive correlation between BIP and SCZ was observed***

*Each dot represents a SNP. On the x-axis is the genomic position in base pairs. The y-axis shows the effect size of the SNP as standardized beta. Color saturation is proportional to significance level of the SNP (with darker colors representing lower p-values). SCZ SNP alleles (in green) were aligned to only show positive betas and used as a reference to align alleles of BIP (in purple).*

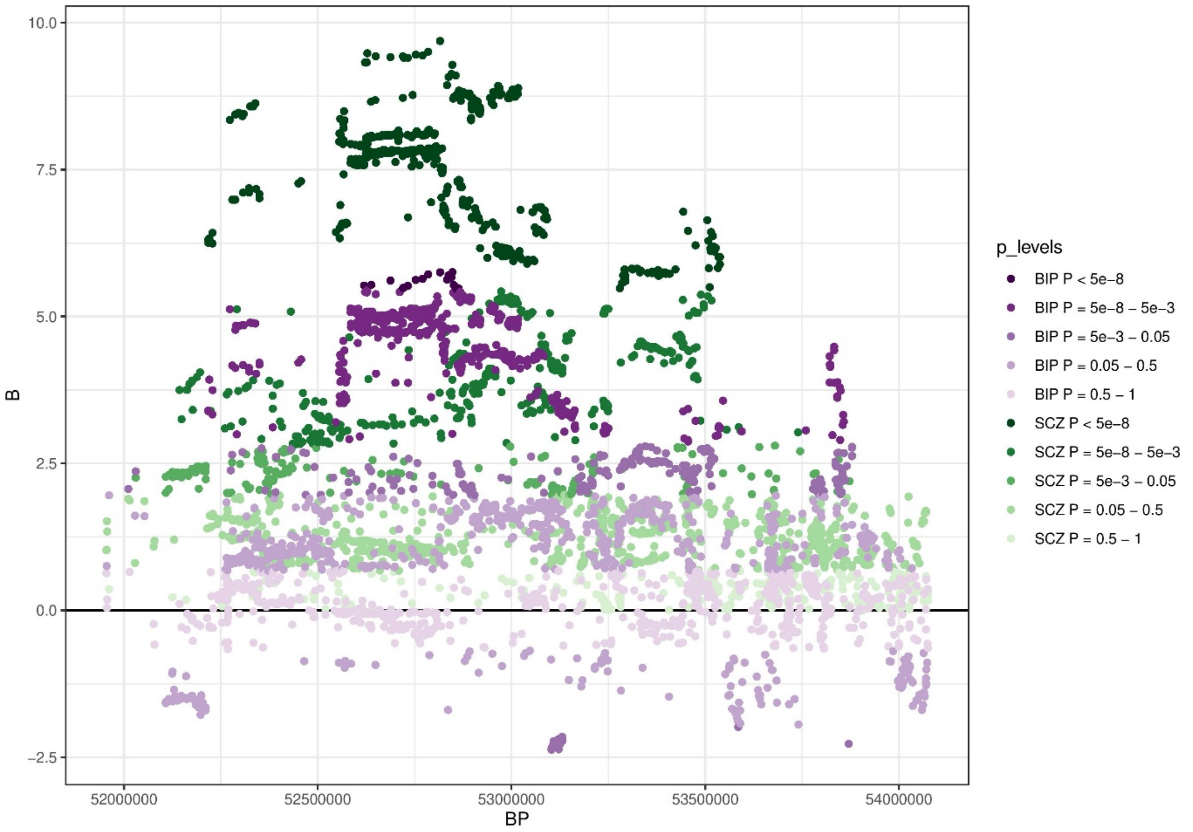

***Fig. SN5. Standardized betas of SNPs within region chr3:51,953,969-54,074,844 where a strong, positive correlation between BIP and SCZ was observed***

*Each dot represents a SNP. On the x-axis is the genomic position in base pairs. The y-axis shows the effect size of the SNP as standardized beta. Color saturation is proportional to significance level of the SNP (with darker colors representing lower p-values). SCZ SNP alleles (in green) were aligned to only show positive betas and used as a reference to align alleles of BIP (in purple).*

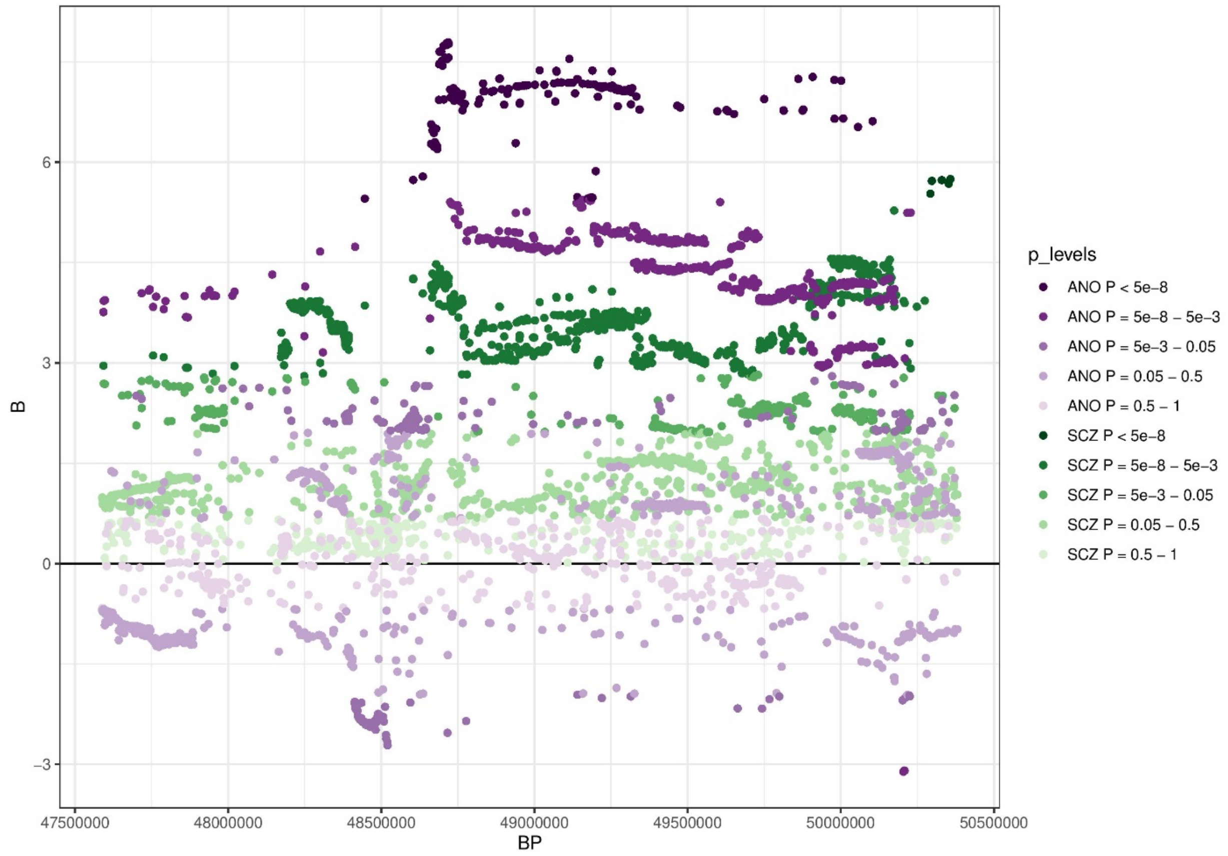

***Fig. SN6. Standardized betas of SNPs within region chr3:47,588,462-50,387,742 where a strong, positive correlation between ANO and SCZ was observed***

*Each dot represents a SNP. On the x-axis is the genomic position in base pairs. The y-axis shows the effect size of the SNP as standardized beta. Color saturation is proportional to significance level of the SNP (with darker colors representing lower p-values). SCZ SNP alleles (in green) were aligned to only show positive betas and used as a reference to align alleles of ANO (in purple).*

### Supplementary Note 4. Assessing gene function convergence across PDs' mapped genes

Convergence in functional pathways based on genes mapped in PDs can illuminate biological origins of comorbidity. Taking a different approach than the MAGMA gene-set analysis, we conducted gene-set enrichment tests within expert curated databases using positionally mapped genes as well as expression quantitative trait locus (eQTL) and chromatin interaction mapped genes as defined in FUMA. For each PD, the number of genes mapped by FUMA was 32 for ADHD, 68 for ALC, 76 for ANO, 11 for ANX, 12 for ASD, 116 for BIP, 12 for DEP, 132 for INS, 0 for OCD, 2 for PTSD, 1,674 for SCZ and 0 for TS.

Previous studies have highlighted the synapse as a hub for shared liability across PDs[4,5]. To assess the degree to which mapped genes converge in functional synaptic processes, we conducted enrichment analyses through SynGO[6], an expert-curated resource for synapse function. We selected PDs with synaptic annotation for more than two genes: ADHD (n of synaptic genes = 3), ANO (9), BIP (6), DEP (4), INS (8), and SCZ (83). All of these contained at least one gene with functional annotations to *chemical synaptic signaling,* and all but BIP contained at least one gene related to *synapse adhesion between pre- and post-synapse*. However, only ANO, BIP, and SCZ showed significant enrichment for *synaptic signaling*, and ANO and SCZ for *synapse organization* (*P*-values between $1 \times 10^{-5}$ and $1 \times 10^{-10}$; **Fig. SN7**).

Another way in which genes can be related to each other is through gene duplication events, which occurs when a region of the genome duplicates and creates two identical copies of the same gene. Over time these related genes can diverge in specific properties due to different mutations. Such genes are termed *paralogs* and constitute gene-sets to identify gene families and consequently genes of similar function that are related across PDs. Mapped genes from all PDs were annotated to their paralog ID[5] and paralog families containing gene members from three traits or more were retained (Methods). Following this procedure, 12 paralog families were retained, none containing genes from more than four traits (**Table SN4**). SCZ, INS, BIP, and ADHD all contained genes from the *MADS box transcription enhancer factor 2 family* which regulate specific gene expression changes in several developmental and adaptive responses[2]. ANO, ANX, INS, and SCZ all contained genes from the *immunoglobulin superfamily* which consists of cell surface and soluble proteins that play a wide role in cell recognition, binding and adhesion processes. This overlap suggests that certain gene families are overrepresented among PDs and, along with the SynGO enrichments, suggests clues to processes which may be common to multiple PDs.

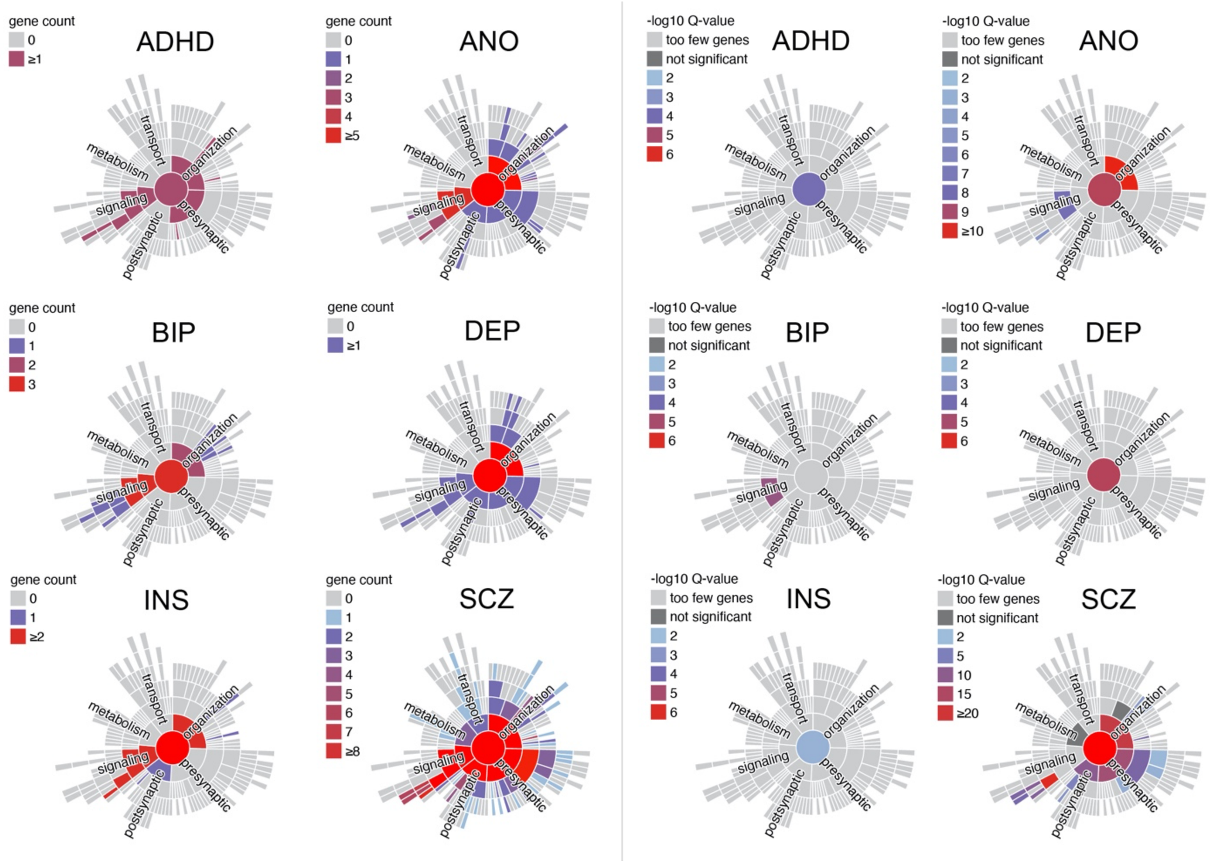

**Fig. SN7.** ***SynGO result of gene count and enrichment test for ADHD, ANO, DEP, BIP, INS, and SCZ using mapped genes.***

*Gene count in synaptic processes to the left and q-value of enrichment test to the right. All figure panels were downloaded from the SynGO platform.*

Table SN4. Paralog groups overlapping across PDs

| **Paralog_set_id** | **PD** | **GENE** | **Gene method** | **gene_id** | **gene_biotype** |
|---|---|---|---|---|---|
| **87** | ALC | RAB10 | FUMA | ENSMUSG00000020671 | protein_coding |
| **87** | INS | RAB1A | FUMA | ENSMUSG00000020149 | protein_coding |
| **87** | INS | RAB1B | FUMA | ENSMUSG00000024870 | protein_coding |
| **87** | SCZ | RAB1B | FUMA | ENSMUSG00000024870 | protein_coding |
| **87** | SCZ | RAB13 | FUMA | ENSMUSG00000027935 | protein_coding |
| **169** | ADHD | MEF2C | MAGMA | ENSMUSG00000005583 | protein_coding |
| **169** | BIP | MEF2B | FUMA | ENSMUSG00000079033 | protein_coding |
| **169** | INS | SRF | FUMA | ENSMUSG00000015605 | protein_coding |
| **169** | SCZ | SRF | MAGMA | ENSMUSG00000015605 | protein_coding |
| **169** | SCZ | MEF2B | FUMA | ENSMUSG00000079033 | protein_coding |
| **257** | ALC | PPM1G | FUMA | ENSMUSG00000029147 | protein_coding |
| **257** | BIP | PPM1M | FUMA | ENSMUSG00000020253 | protein_coding |
| **257** | SCZ | PPM1M | FUMA | ENSMUSG00000020253 | protein_coding |
| **257** | SCZ | PDP2 | FUMA | ENSMUSG00000048371 | protein_coding |
| **257** | SCZ | TAB1 | MAGMA | ENSMUSG00000022414 | protein_coding |
| **428** | ADHD | PTPRF | MAGMA | ENSMUSG00000033295 | protein_coding |
| **428** | DEP | PTPN1 | MAGMA | ENSMUSG00000027540 | protein_coding |
| **428** | SCZ | PTPRF | MAGMA | ENSMUSG00000033295 | protein_coding |
| **758** | DEP | NEGR1 | FUMA | ENSMUSG00000040037 | protein_coding |
| **758** | INS | LSAMP | MAGMA | ENSMUSG00000061080 | protein_coding |
| **758** | SCZ | NEGR1 | MAGMA | ENSMUSG00000040037 | protein_coding |
| **758** | SCZ | OPCML | MAGMA | ENSMUSG00000062257 | protein_coding |
| **758** | SCZ | LSAMP | FUMA | ENSMUSG00000061080 | protein_coding |
| **883** | BIP | NEK4 | MAGMA | ENSMUSG00000021918 | protein_coding |
| **883** | INS | MAP3K14 | FUMA | ENSMUSG00000020941 | protein_coding |
| **883** | SCZ | MAP3K14 | FUMA | ENSMUSG00000020941 | protein_coding |
| **883** | SCZ | NEK4 | MAGMA | ENSMUSG00000021918 | protein_coding |
| **883** | SCZ | TAOK2 | MAGMA | ENSMUSG00000059981 | protein_coding |
| **1174** | ANO | NCAM1 | MAGMA | ENSMUSG00000039542 | protein_coding |
| **1174** | ANX | NCAM1 | MAGMA | ENSMUSG00000039542 | protein_coding |
| **1174** | INS | SDK1 | MAGMA | ENSMUSG00000039683 | protein_coding |
| **1174** | SCZ | SDK1 | MAGMA | ENSMUSG00000039683 | protein_coding |
| **1174** | SCZ | CNTN4 | MAGMA | ENSMUSG00000064293 | protein_coding |
| **1174** | SCZ | IGSF9B | MAGMA | ENSMUSG00000034275 | protein_coding |
| **1174** | SCZ | DCC | MAGMA | ENSMUSG00000060534 | protein_coding |
| **1633** | ADHD | KDM4A | MAGMA | ENSMUSG00000033326 | protein_coding |

| Paralog_set_id | PD | GENE | Gene method | gene_id | gene_biotype |
|---|---|---|---|---|---|
| **1633** | INS | KDM4B | MAGMA | ENSMUSG00000024201 | protein_coding |
| **1633** | SCZ | KDM4A | MAGMA | ENSMUSG00000033326 | protein_coding |
| **1728** | BIP | TRANK1 | FUMA | ENSMUSG00000062296 | protein_coding |
| **1728** | INS | FAM89B | FUMA | ENSMUSG00000024939 | protein_coding |
| **1728** | SCZ | FAM89B | FUMA | ENSMUSG00000024939 | protein_coding |
| **1728** | SCZ | TRANK1 | FUMA | ENSMUSG00000062296 | protein_coding |
| **1736** | BIP | TLR9 | FUMA | ENSMUSG00000045322 | protein_coding |
| **1736** | DEP | LRFN5 | FUMA | ENSMUSG00000035653 | protein_coding |
| **1736** | SCZ | TLR9 | FUMA | ENSMUSG00000045322 | protein_coding |
| **1736** | SCZ | LRRN3 | FUMA | ENSMUSG00000036295 | protein_coding |
| **2085** | ADHD | SORCS3 | MAGMA | ENSMUSG00000063434 | protein_coding |
| **2085** | DEP | SORCS3 | MAGMA | ENSMUSG00000063434 | protein_coding |
| **2085** | SCZ | SORCS3 | MAGMA | ENSMUSG00000063434 | protein_coding |
| **2087** | ALC | KHK | FUMA | ENSMUSG00000029162 | protein_coding |
| **2087** | ANO | ADK | MAGMA | ENSMUSG00000039197 | protein_coding |
| **2087** | SCZ | RBKS | MAGMA | ENSMUSG00000029136 | protein_coding |

*Note: Retained paralog families across traits with mapped genes from three traits or more.*
